# Hydroxychloroquine for SARS-CoV-2 positive patients quarantined at home: The first interim analysis of a remotely conducted randomized clinical trial

**DOI:** 10.1101/2021.02.22.21252228

**Authors:** Ravi K. Amaravadi, Lydia Giles, Mary Carberry, Matthew C. Hyman, Ian Frank, Sunita D. Nasta, Jennifer Walsh, E. Paul Wileyto, Phyllis Gimotty, Michael Milone, Edith M. Teng, Niraj J. Vyas, Steve Balian, Jonathan A. Kolansky, Nabil M. Abdulhay, Shaun K. McGovern, Sarah Gamblin, Olivia Doran, Paul L. Callahan, Benjamin S. Abella

**Affiliations:** Abramson Cancer Center and Division of Hematology-Oncology, Department of Medicine, University of Pennsylvania; Division of Cardiology, Department of Medicine University of Pennsylvania; Division of Infectious Disease, Department of Medicine University of Pennsylvania; Department of Microbiology, University of Pennsylvania; Department of Biostatistics, Epidemiology and Informatics, University of Pennsylvania; Department of Pathology, University of Pennsylvania; Department of Emergency Medicine, University of Pennsylvania

**Keywords:** COVID-19, hydroxychloroquine

## Abstract

**Background:** Older patients are at risk of increased morbidity and mortality from COVID-19 disease due to Severe Acute Respiratory Syndrome Coronavirus 2 (SARS-CoV-2). There are few effective treatments for outpatients with COVID-19.

**Objective:** To evaluate the efficacy of hydroxychloroquine to reduce time in quarantine for symptomatic ≥40 years-old COVID-19 patients.

**Design:** A randomized, double-blind, placebo-controlled clinical trial.

**Setting:** Outpatients with polymerase chain reaction confirmed COVID-19 at a University of Pennsylvania affiliated testing center between April 15, 2020 and, July 14, 2020.

**Participants:** Out of 5511 SARS-CoV-2 positive patients, 1072 met initial eligibility criteria for telephone-based recruitment, but only 34 subjects were able to be randomized.

**Interventions:** Hydroxychloroquine 400 mg per twice daily (n=17) or matching placebo (n=17), taken orally for up to 14 days.

**Measurements:** The primary outcome was the time to release from quarantine. Secondary outcomes included the participant-reported secondary infection of co-inhabitants, hospitalization, treatment-related adverse events, time to symptom improvement, and incidence of cardiac arrhythmia.

**Results:** The median time to release from quarantine for HCQ-treated vs. placebo-treated participants was 8 days (range 4-19 days) vs. 11 days (4-18 days); z-score +0.58, p=n.s. This did not meet the pre-specified criteria for early termination, however, this study was terminated early due to lack of feasibility. There was no mortality in either study arm.

**Limitation:** Since this study was terminated early due to a lack of feasibility, no conclusion can be made about the efficacy of hydroxychloroquine as a treatment for COVID-19 patients 40 years of age or older quarantined at home.

**Conclusion:** The design of this remotely conducted study could guide testing of other more promising agents during the COVID-19 pandemic.

**Trial registration:** Clinicaltrials.gov identifier: NCT04329923

---

Coronavirus Disease 2019 (COVID-19) caused by Severe Acute Respiratory Syndrome Coronavirus 2 (SARS-CoV-2) has afflicted millions of people worldwide. Pharmacologic treatment of COVID-19 has proven challenging despite intensive research efforts. Older COVID-19 patients with pre-existing conditions and symptoms are at greatest risk of morbidity and mortality, but not every such patient meets criteria for hospitalization (1). Many such patients, especially early in the course of disease, are quarantined at home, but worsen and end up in the hospital. African-American patients are more likely to be hospitalized and die compared to matched Caucasian populations (2). Time in quarantine leads to reduced wages, loss of work, and infection of co-inhabitants, magnifying the impact of any one infection (3). Hydroxychloroquine (HCQ) has been shown to clear SARS-CoV-2 cellular infection in vitro (4). Based on these preclinical findings, dozens of COVID-19 treatment studies with HCQ have been launched throughout the world, mostly focused on hospitalized patients. To test the hypothesis that HCQ treatment earlier in the course of disease may be more effective, we designed a double-blind placebo -controlled randomized study in symptomatic COVID-19 patients quarantined at home who developed symptoms within 4 days of testing. This trial was called sub-study 1 (SS1) of the Prevention and Treatment of COVID-19 with Hydroxychloroquine (PATCH) trial. In order to assure the safety of research staff, and the community, this study was designed to be conducted completely in a remote fashion with no in-person visits with the patients after the time of testing.

## METHODS

Subject enrollment and participation spanned from April 15, 2020 to July 14, 2020 within the Penn Medicine health system located in Philadelphia, Pennsylvania. An independent Medical Monitor, Data Safety Monitoring Board (DSMB), and COVID-19 Trials Steering Committee provided oversight of safety and efficacy endpoints. Written informed consent was obtained from all subjects via direct communication with study physician via an internet document signature program (DocuSign). Approval for the study was granted by the University of Pennsylvania Institutional Review Board, and the trial was registered on clinicaltrials.gov (NCT04329923).

### Study Participants

Participants were eligible for inclusion if they met the following inclusion criteria: 1) Age ≥40 years, 2) PCR-positive for the SARS-CoV-2 virus, 3) fever, or cough, or shortness of breath at the time of testing, 4) ≤4 days had elapsed since the first COVID-19 symptom and testing, 5) not taking azithromycin at the time of enrollment, 6) symptomatic at the time of enrollment, 7) did not require hospitalization, 8) lived within 30 miles of Hospital of the University of Pennsylvania, 9) had access to a working computer, or smartphone and have internet access, 10) willing to fill out a daily electronic symptom score, 11) available for a daily phone call, 12) willing to take their own temperature twice a day, and 13) willing to report the observed symptoms and development of COVID-19 in co-habitants. Exclusion criteria included: 1) known history of allergy or sensitivity to HCQ, 2) history of glucose-6-phosphate dehydrogenase deficiency, 3) history of retinal disease, 4) history of significant cardiac disease, 5) current use of tricyclic antidepressants, and other medications known to prolong the QT interval, 6) history of psoriasis, 7) history of significant lung disease, 8) participation in any other pharmacologic research study for COVID-19, and 9) pregnancy or intention to become pregnant within the study period.

### Group assignments and study medications

**Supplemental Figure 1A** shows the study design. Subjects were randomized to either HCQ or placebo in blocks of 8, using established randomization software (SealedEnvelope.com, Clerkenwell Workshops). Subjects assigned to the HCQ arm received hydroxychloroquine 200 mg tablets (provided by Sandoz, a division of Novartis Pharmaceuticals), with instructions to take two tablets twice a day with food. Subjects assigned to the placebo arm received custom-molded identical size/shape microcrystalline cellulose tablets (prepared for this trial by Investigational Drug Service, Temple University, Philadelphia) and given identical instructions.

### Remote recruitment and enrollment

Figure 1. shows the series of procedures employed to recruit and enroll subjects. All subject interactions were conducted remotely without in-person visits. Recruitment of participants was enabled by a SARS-CoV-2 PCR positive alert in the EMR, which the clinical research coordinators (CRC) on the study team screened daily. PCR-positive individuals who met the initial screening criteria of age > 40, fever or cough or shortness of breath, an address within 30 miles, and who had been already contacted by clinical providers to inform them of the COVID-19 diagnosis were eligible for an initial telephone call. If the patient was interested, the CRC would email a PDF of the informed consent form for the study candidate to review. All study communication between CRC and study clinical investigators was performed on a HIPAA – compliant platform (Teams, Microsoft Corporation) administered by the University of Pennsylvania, enabling rapid communication of Protected Health information. Interested study candidates were then contacted by telephone by a physician investigator to obtain consent. Written informed consent was obtained through Docusign, followed by a repeat phone call by the physician investigator to assess eligibility with full access to the patient’s EMR.

**Figure 1.**
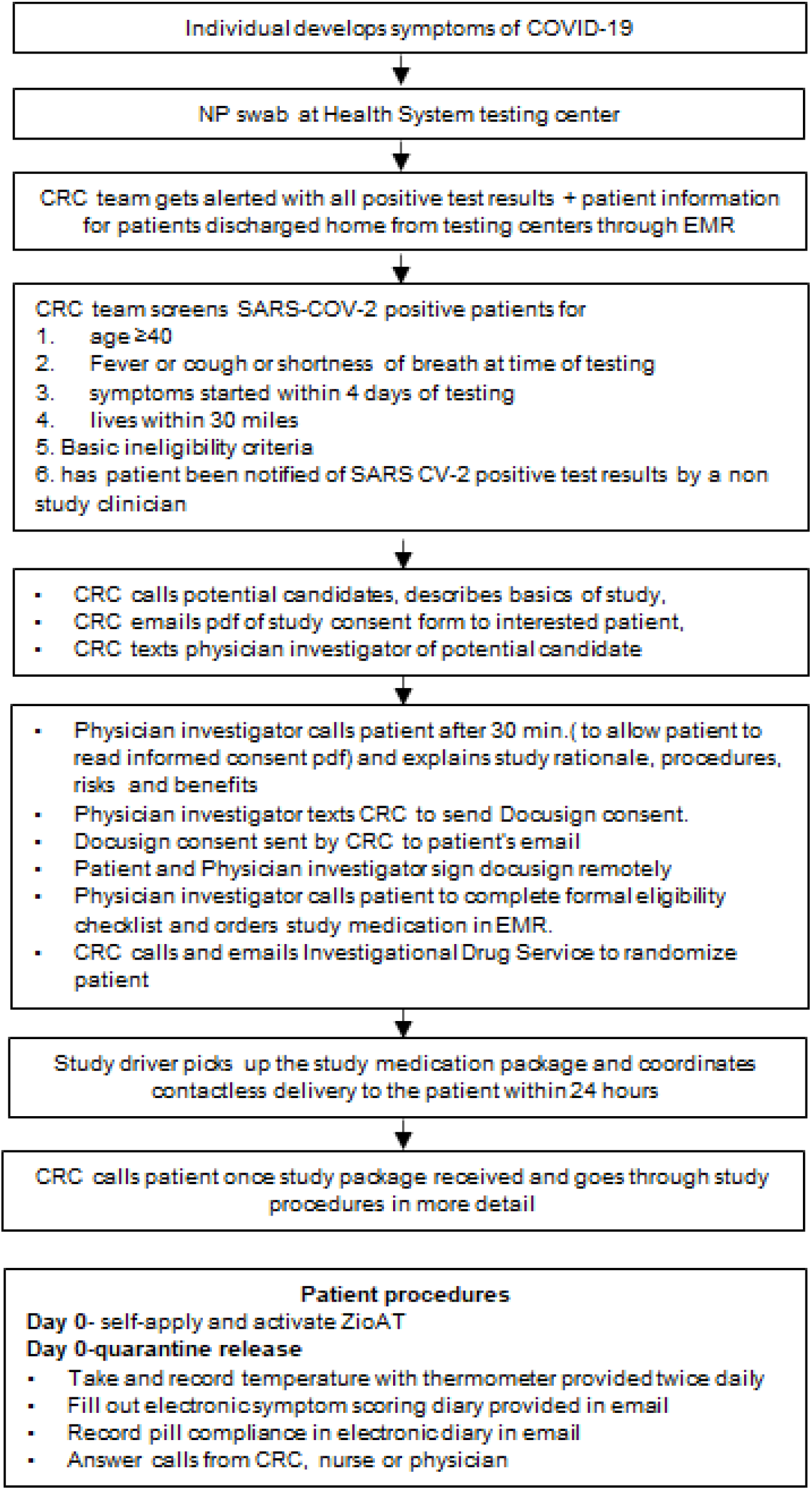
Procedure for remote enrollment of COVID-19 patients. Amaravadi et al. Hydroxychloroquine for outpatients with COVID-19

### Patient conducted study procedures

Once randomized, the study team would perform contactless delivery of a study package by van to the participants with study medication followed by further telephone instructions. Day 0 was defined as the day the patient started study medication. For safety, a protocol amendment was made after initial study enrollment had commenced to include cardiac arrhythmia monitoring. A mobile telemetry patch (ZioAT™, iRhythm Technologies) was included with the study package and trial participants were asked to self-apply and activate the device (see Supplemental Study Methods). Subjects completed a daily electronic survey via email (REDCap) to track symptoms, fever, and pill compliance. Daily telephone calls were conducted by the study team to assess adverse events (AE) according to Common Terminology Criteria for Adverse Events (CTCAE) v.5.0 grading scale.

#### Clinical symptom monitoring

Patients filled out electronic symptom diary that tracked COVID-19 symptoms (Cough, shortness of breath, fatigue, muscle aches, diarrhea, abdominal pain, loss of smell, other). In addition to track potential HCQ AEs, nausea and vomiting, loss of appetite, rash and visual disturbance was scored daily but did not contribute to the COVID-19 symptom score. For each symptom patients would report a score of 1 (mild), 2 (moderate), or 3 (severe). A computer algorithm would then generate the daily COVID-19 symptom score for the patient. An example of this scoring system is provided in eTable1. Each day participants were to provide their daily patient-reported outcomes, and if this information was not provided, the study team would collect these data directly from the patient on the telephone.

#### Adverse Event (AE) assessment

HCQ treatment can acutely cause nausea, diarrhea and rash, and visual disturbance after months of treatment. Diarrhea and nausea overlap with the COVID-19 symptoms. If an overlapping symptom was worsening while other symptoms were resolving it was reclassified as a treatment-related AE. Cardiac AEs were assessed by a cardiologist.

#### Triage

Every participant’s symptoms, fever pattern and pill compliance was discussed daily on virtual rounds. Symptoms were assessed as likely related to COVID-19 or possibly related to study treatment. If the latter, a research nurse would call and assess the symptom directly and discuss with the physician investigator. If a dose reduction was required, the physician investigator would call the patient and explain the reduction. If COVID-19 symptoms were worsening, the physician investigator would call the patient to determine if there was a need to send the patient to the hospital.

#### Temperature measurements

Participants were asked to record their temperature twice daily with a provided thermometer.

#### Study completion and release from quarantine (RFQ)

Once the criteria for RFQ were met, the physician investigator informed the participant to stop taking study medication, remove and return the ZioAT device directly to iRhythm through the mail.

#### Outcome measures

The primary outcome measure was the median time to RFQ, which was allowed when the following US Centers for Disease Control criteria were met: 1) no fever for 72 hours without the use of fever-reducing medications, 2) improvement in other symptoms, and 3) 7 or 10 days have elapsed since symptom onset. The first date of quarantine was considered the date of SARS-CoV-2 nasopharyngeal swab test that confirmed positivity. Secondary outcome measures included rate of participant-reported secondary infection of co-habitants, rate of hospitalization, rate of treatment related adverse events, and time to symptom improvement.

#### Statistical power and analysis

The CDC guidelines for RFQ changed during the course of this study. The criteria were: 1) 72 hours without a fever, 2) improvement in symptoms, and 3) Originally 7 days, but changed to 10 days mid-study, since the beginning of symptoms have elapsed. Our original null hypothesis was that in the placebo cohort the median time to RFQ would be 10 days. HCQ would be considered more effective if patients had a median time to RFQ of 5 days on study drug. With 50 subjects each in the placebo and HCQ arms, the one-sided z-test (alpha=.05) had an overall 95% power to detect a significant difference between the two groups with medians of 10 and 5 days. Based on the new CDC guidelines for quarantine release (10 days) the null hypothesis was a median time to RFQ in the placebo cohort of 14 days. HCQ would be considered more effective if the median time to RFQ was 7 days. The one-sided z-test (alpha=.05) had an overall 93% power to detect a significant difference between the two groups with medians of 14 and 7 days. The *hypothesis* was tested using a one-sided test using a z-score corresponding to the log of the hazard-ratio for RFQ between the two groups. Two interim analyses targeting 34% and 68% completion of participant enrollment were planned to test for early efficacy or futility, using z-score boundaries that follow Hwang-Shih-DeCani alpha spending rules (5). The study protocol and statistical analysis plan are included as Supplement 1 and Supplement 2.

## RESULTS

### Accrual

During the study accrual period, the Philadelphia region experienced a peak of infection rates followed by a relatively rapid decline in cases (**Supplemental Figure 1B**). Accrual to the study mirrored the peak and decline of infection rate in the area (Supplemental **Figure 1C**). We screened 5511 COVID-19 positive patients and identified 1072 who were possibly eligible based on age and symptoms recorded at the time of testing (**Figure 2**). Nearly 40% (407/1072) of the potentially eligible subjects met an ineligibility criterion either in the medical chart or after telephone review (**Figure 2**). Roughly 20% were not interested in the study, and the remainder were not called or not reached due to a number of practical reasons, including candidates that indicated initial interest but then did not return calls. Ultimately, only 36 subjects electronically signed informed consent. Two subjects met protocol exclusion criteria after signing informed consent and undergoing full eligibility screening by a physician investigator leaving 34 patients eligible for randomization. Six randomized patients failed to complete the study leaving 28 patients for analysis of the primary outcome.

**Figure 2.**
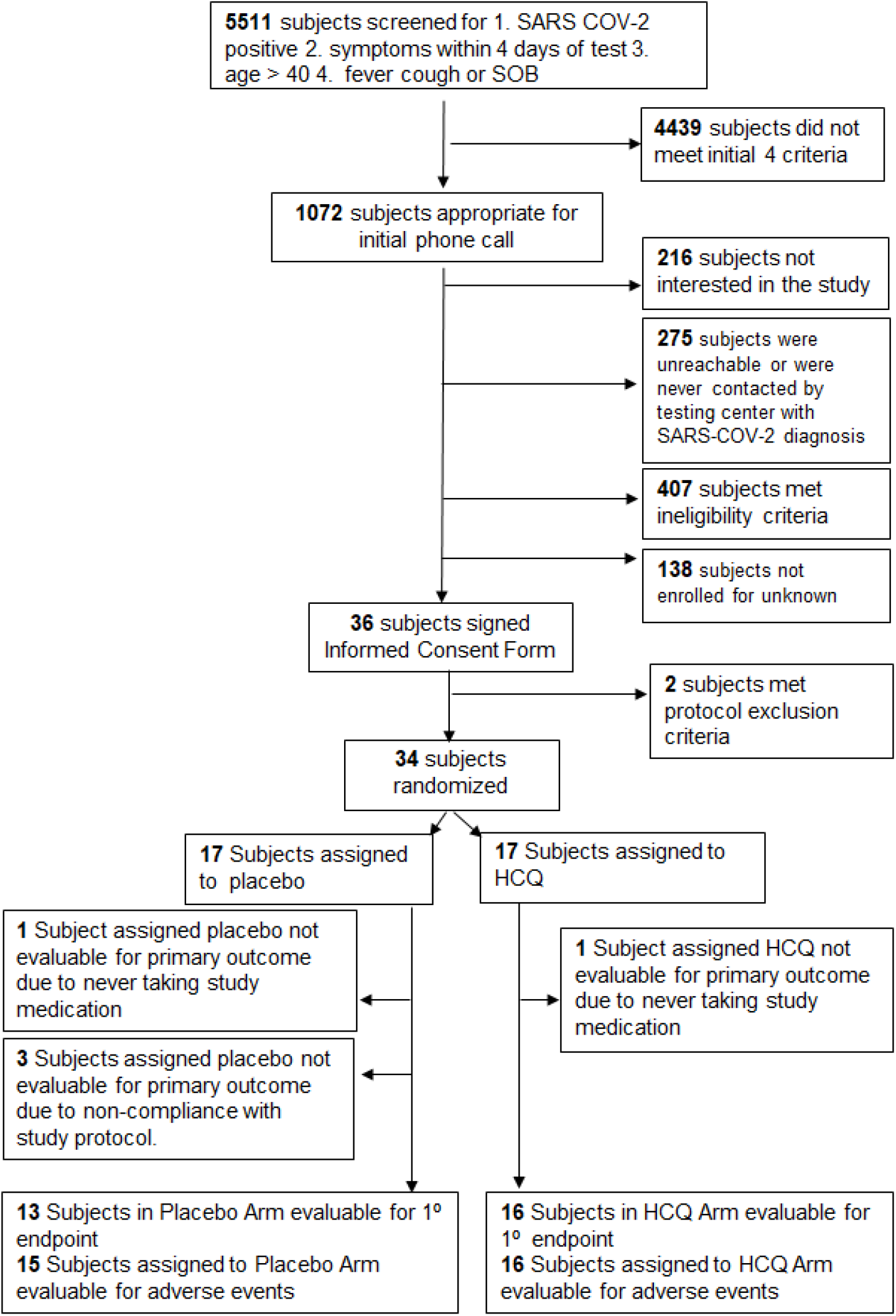
Recruitment and randomization of participants into HCQ and Placebo.

### Demographics and baseline symptoms

Recruited patients had a median age of 53 (range 40-80), and were mostly female (62%), and non-white (77%) with few co-morbidities (**Table 1**). The only symptom that differed significantly between the two groups was cough at the time of testing **(Supplemental Table 2)**. The presence of fever decreased significantly for both groups between day of testing and day 0 (**Supplemental Table 3)**. All baseline symptoms for subjects on Day 0 are presented in **Table 2**. There were no significant differences between the two groups for any baseline symptoms.

**Table 1.**
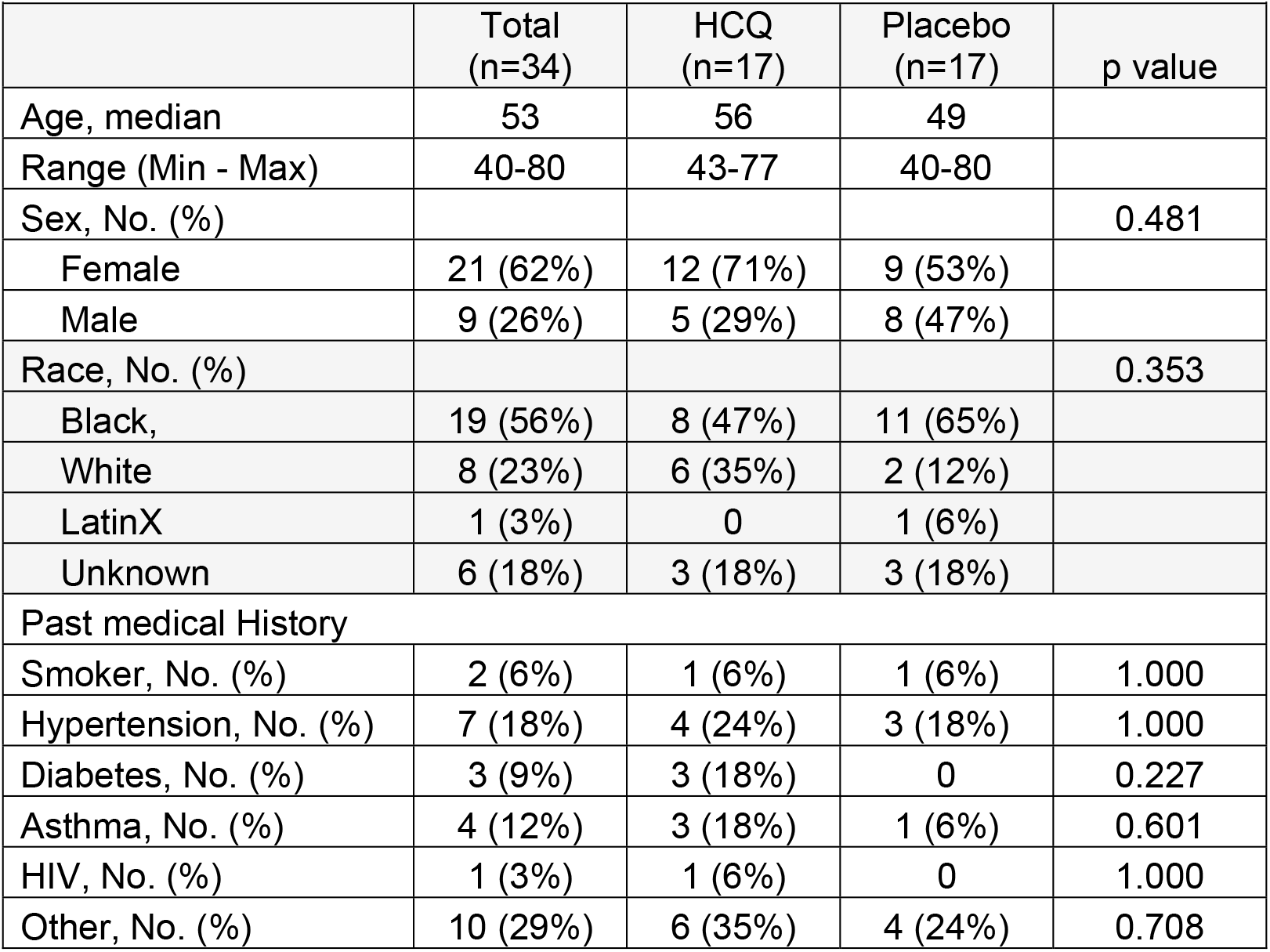
Sub-study 1 Demographics

**Table 2.** All baseline Symptoms on Day 0.

| **Table 2. All baseline Symptoms on Day 0.** |  |  |  |  |  |
| --- | --- | --- | --- | --- | --- |
|  | HCQ |  | Placebo |  | p-value |
|  | n | No. (%) | n | No. (%) |  |
| Fever | 17 | 5 (29) | 17 | 5 (29) | 1.000 |
| Cough | 17 | 10 (59) | 17 | 15 (88) | 0.118 |
| Shortness of breath | 17 | 5 (29) | 17 | 5 (29) | 1.000 |
| Fatigue | 17 | 11 (65) | 17 | 10 (59) | 1.000 |
| Myalgia | 17 | 7 (41) | 17 | 11 (65) | 0.303 |
| Headache | 17 | 5 (29) | 17 | 10 (59) | 0.166 |
| Loss of smell | 17 | 7 (41) | 17 | 7 (41) | 1.000 |
| Loss of taste | 17 | 3 (18) | 17 | 1 (6) | 0.601 |
| Loss of appetite | 17 | 11 (65) | 17 | 14 (82) | 0.438 |
| Nausea or vomiting | 17 | 1 (6) | 17 | 5 (29) | 0.175 |
| Diarrhea | 17 | 4 (24) | 17 | 6 (35) | 0.708 |
| Visual disturbance | 17 | 1 (6) | 17 | 1 (6) | 1.000 |
| Other | 17 | 5 (29) | 17 | 5 (29) | 1.000 |

**Table 3.**
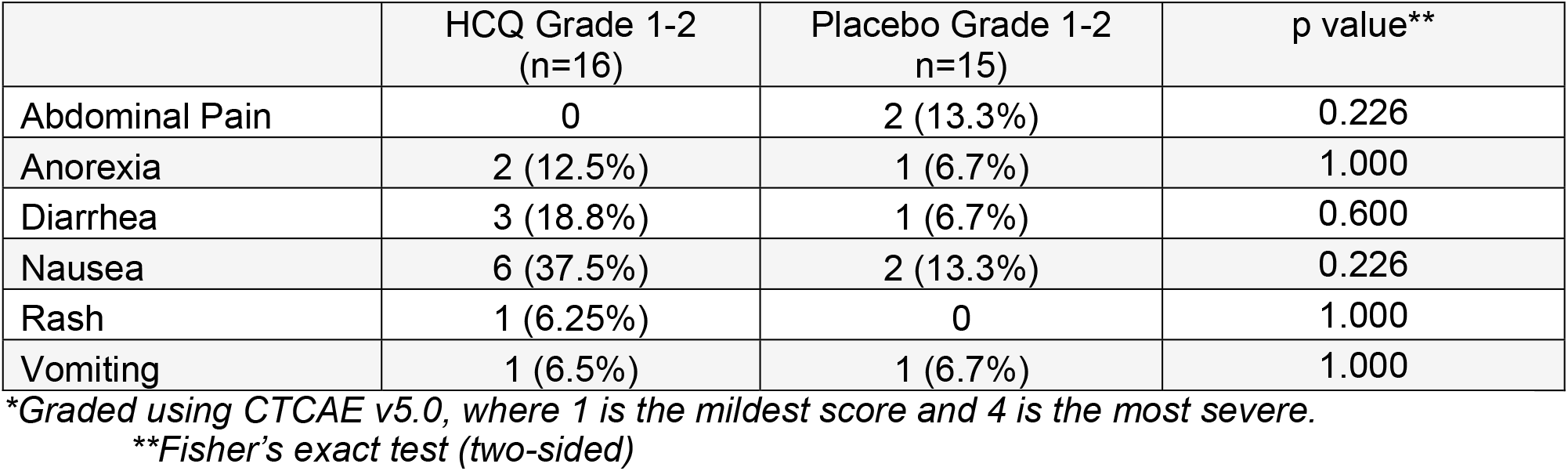
Adverse Events (possibly related to study medication)*

| **Table 3. Adverse Events (possibly related to study medication)\*** |  |  |  |
| --- | --- | --- | --- |
|  | HCQ Grade 1-2<br>(n=16) | Placebo Grade 1-2<br>(n=15) | p value** |
| Abdominal Pain | 0 | 2 (13.3%) | 0.226 |
| Anorexia | 2 (12.5%) | 1 (6.7%) | 1.000 |
| Diarrhea | 3 (18.8%) | 1 (6.7%) | 0.600 |
| Nausea | 6 (37.5%) | 2 (13.3%) | 0.226 |
| Rash | 1 (6.25%) | 0 | 1.000 |
| Vomiting | 1 (6.5%) | 1 (6.7%) | 1.000 |

\*Graded using CTCAE v5.0, where 1 is the mildest score and 4 is the most severe.
\*\*Fisher's exact test (two-sided)

### RFQ

Although the criteria for RFQ changed during the course of this study, for this interim analysis we analyzed all 28 patients (HCQ n=15, Placebo n=13), evaluable for primary outcome in one analysis. The median time to RFQ For HCQ was 8 (range 4-19) days, and for placebo 11 (range 4-18) days. A test of medians resulted in a z-score of +0.58 with a p-value (one-sided) of 0.28 (**Supplemental Figure 2**).

### Hospitalization

One patient assigned to HCQ was hospitalized during the study. This subject was very short of breath at baseline, and was admitted for hypoxia and COVID-19 pneumonia on day 0. The Medical Monitor concluded this hospitalization was not due to an AE from study medication. Per protocol the subject was removed from the study when hospitalized.

### Crossover

5 subjects (2 in the HCQ arm and 3 in the placebo arm) developed progressive symptoms (an increase in symptoms score of 2 points on any day on study) while on study and were unblinded after completing 7 days of their originally assigned study treatment. Of the two subjects who were assigned to HCQ, one was not fully compliant with the study medication until she was unblinded, Once she started taking study medication as prescribed she experienced rapid recovery (**Supplemental Figure 3**, blue line). The other subject who progressed on HCQ while the other was fully compliant. Of the 3 participants who were originally assigned to placebo, all three crossed over to HCQ and had a rapid recovery avoiding hospitalization (**Supplemental Figure 3**).

### Adverse Events and mobile telemetry

The one patient assigned to HCQ treatment hospitalized for shortness of breath was the only SAE. There were no grade 3-4 adverse events in the remaining patients (**Table 3**). However, there was a significant increase in the rate of grade 1 and 2 diarrhea in subjects treated with HCQ compared to placebo. No cardiac events were observed in this study in either arm. Seventeen subjects wore the ZioAT unit during the study; none had sustained ventricular arrhythmias or episodes of atrial fibrillation. QTc measurements at baseline prior to the first dose of study medication and on the day of quarantine release showed that subjects treated with HCQ (n=12) had a median ΔQTc of 9.5 milliseconds (minimum = −22 and maximum = 40) while placebo-treated participants had a median ΔQTc of 2 milliseconds (minimum = −11 and maximum= 11). The two medians were not significantly different (Wilcoxon two-sample, p=0.370) (**Supplemental Figure 4)**.

### Infection of co-inhabitants of study participants

Roughly 40% of participants had co-inhabitants during the study. While more HCQ-treated participants had baseline positive co-inhabitants than placebo-treated patients, in both groups, co-inhabitants of 2 HCQ-treated and 2 placebo-treated participants developed COVID-19 during the course of the study.

### Improvement in COVID-19 Symptoms

Among those subjects who were released from quarantine only 3 people had complete resolution of COVID-19 symptoms. The median time to the first occurrence of the lowest symptom score was 4 days (range: 1 to 14 days) in the HCQ arm and was 7 days (range: 1 to 11 days) for the Placebo arm; the exact test for the equality of the medians was not significant (Log-rank, p=0.212) (**Supplemental Figure 5**).

### Interim analysis, Data Safety and Monitoring Board review and decision to terminate early

Because of the slow accrual, we modified the interim analysis to accommodate the realized 28% information rather than the projected 34%. A test of medians resulted in a z-score of +0.58 with a p-value (one-sided) of 0.28. The analysis plan was based on Cox regression. The z-statistic from the Hazard Ratio (HR) following Cox regression was estimated to be HR=1.10 (95%CI: 0.51-2.34), with a corresponding z-score of 0.24 (**Supplemental Figure 6**). The z-score of 0.24 falls between the boundary value for significance (z = +2.8954) and futility (z= −1.1049). The DSMB concluded there was no scientific reason to terminate the study. However, based on the large numbers of patients screened with few enrolled (**Figure 1**), growing literature about the lack of efficacy of HCQ for COVID-19, and concern that our study was not likely to change the treatment of COVID-19 during a rapidly evolving treatment landscape, the decision was made to terminate the study early.

## DISCUSSION

This remotely conducted randomized controlled study of a long course of high-dose HCQ for patients 40 years of age or older with COVID-19 quarantined at home was not able to be completed due to poor accrual. Consequently, all findings including the trend towards improved symptoms with HCQ should not be considered evidence of benefit with HCQ. The reasons the results of this interim analysis are informative include 1) demonstration of methodology for conducting entirely remote clinical trials and 2) identification of the challenges with recruitment that such an approach faces, in the setting of shifting evidence pertaining to HCQ that may have impacted subject interest in the study.

Out of 1072 potentially eligible candidates for this trial, only 34 were able to be enrolled. There was no significant difference in time to RFQ, adverse events, or cardiac events as measured by mobile cardiac telemetry. There was a nonsignificant trend towards faster improvement in COVID-19 symptoms in patients receiving HCQ compared to placebo. While the results of our comparison of HCQ and placebo in this population did not lead to a conclusion, our preliminary findings are consistent with the only other published randomized placebo-controlled HCQ trial for outpatients with COVID-19 (6). In this University of Minnesota study, 461 patients were randomized to a 5-day course of HCQ given at 800 mg once and 600 mg daily for 4 days or folate (placebo). Recruitment of patients from many US states and Canadian provinces occurred through an internet website. In order to get drug to patients quickly the study required only symptoms compatible with COVID-19 and not PCR confirmation for eligibility. The minimum age for participants was 18, and the average age of this study population was 40. There were only 3% African-Americans, and only 34% of the patients were SARS-CoV-2 positive by PCR. The primary outcome was a change in patient –reported symptom score which was not significantly different between HCQ and placebo, but patients with HCQ had a slightly faster recovery than patients treated with placebo. Our study was different from the study from the University of Minnesota study, as it recruited patients using direct phone calls to patients who had tested positive for SARS CoV-2. Therefore, all the patients on this study had confirmed symptomatic COVID-19 infection. Our study recruited patients 40 years or older, since older patients have increased mortality with COVID-19. The majority of our participants were non-white, which is also a group at higher risk of poor outcomes.

Many factors contributed to the poor accrual in this study. First the prevalence of COVID-19 in the Philadelphia area declined during the study. Many patients on our study felt better between the time of testing and day of initial telephone call to recruit them. Our focus on patients 40 years of age or older significantly reduced the pool of potential study candidates, especially since if a patient 40 years of age or older had moderate symptoms they could be hospitalized rather than quarantined at home. Recruitment of patients to a remote clinical trial using a “cold call” approach (from investigative staff who had never met the subject in person) proved to be very difficult. Without direct in person contact with a physician, patients were often suspicious or simply did not answer the telephone. Because of the highly infective nature of COVID-19 we could not identify a safer means of recruiting patients.

Perhaps the single most important impediment to recruitment for this study was the unusual publicity HCQ received during the course of this study. Numerous uncontrolled studies, which suggested harm with HCQ treatment in hospitalized patients, emerged in non-peer reviewed platforms and became widely known (7). A global observational study (found subsequently to be fraudulent) published in a high impact peer-reviewed journal concluded that HCQ could increase death in COVID-19 patients (8). Other randomized studies showed no benefit for HCQ in the hospitalized setting, and extensive discussion about the rationale for treating earlier in the course of disease did not resonate with some of the potential candidates for our study (9). The Early Use Authorization for HCQ by the Food and Drug Administration followed by retraction of this guidance confused potential subjects. The politicization of HCQ in the press also served as a point of confusion that limited interest. The unique challenges to recruitment we faced conducting this HCQ study may not be encountered if this model were used to study other medications for infectious disease.

Once patients decided to enroll, the conduct of the study went smoothly, with patient safety maintained and high-quality data was able to be captured. Controlled delivery of study materials to patients’ home was executed without any infection of study personnel. Patients, despite being older in age, suffering from COVID-19 symptoms, and often being isolated from family members, were able to navigate the substantial technology requirements (multiple daily phone calls, email reporting of symptoms, twice daily temperature checks, and self-application of a mobile cardiac telemetry monitor) of this study. The daily data stream coming from patients was sufficient for our medical team to make clinical decisions on patient dispositions. Only one patient was hospitalized and no patients died of COVID-19 on this study. In conclusion, this study was terminated due to external factors, but the data collected demonstrates that an entirely remotely conducted clinical trial is feasible for infectious disease trials in future.

## Supporting information

PATCH protocol

CONSORT checklist PATCH trial

## Data Availability

All data from this report will be available to scientists and clinicians upon a reasonable request

## Funding Source

Philanthropic donations from Leonard and Madlyn Abramson and Mark and Cecilia Vonderheide.

## Acknowledgements

Cardiac arrhythmia monitoring was provided as an in-kind gift by iRhythm Technologies, Inc.

## Notes

### Competing Interest Statement

The authors have declared no competing interest.

### Clinical Trial

NCT04329923

### Funding Statement

The funding sources were exlcusively philanthropic donations made to the University of Pennsylvania by Leonard and Madlyn Abramson and Mark and Cecilia Vonderheide.Cardiac arrhythmia monitoring was provided as an in-kind gift by iRhythm Technologies, Inc.

### Author Declarations

University of Pennsylvania Institutional Review Board

