## Supplemental Figures, Tables, Methods for "Hydroxychloroquine for SARS-CoV-2 positive patients quarantined at home: The first interim analysis of a remotely conducted randomized clinical trial"

**Supplemental Figures, Tables and Methods**

*Amaravadi et al. Hydroxychloroquine for outpatient treatment of COVID-19*

**Supplemental Figures 1-6.**

A

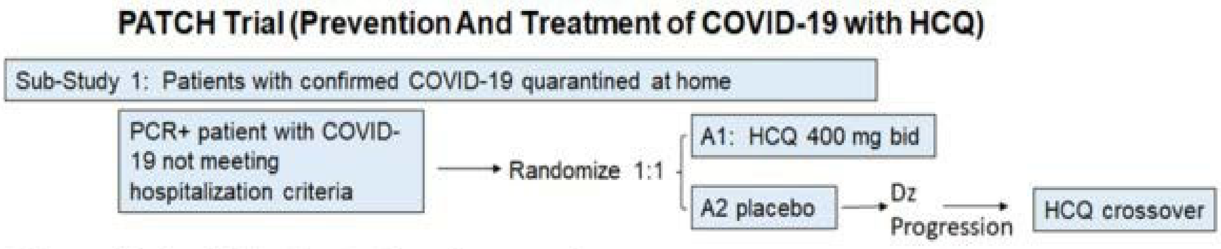


B

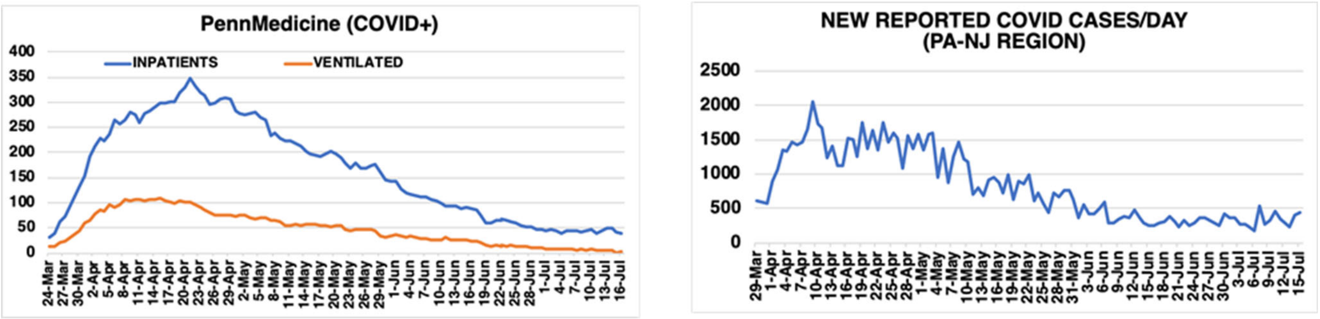


C

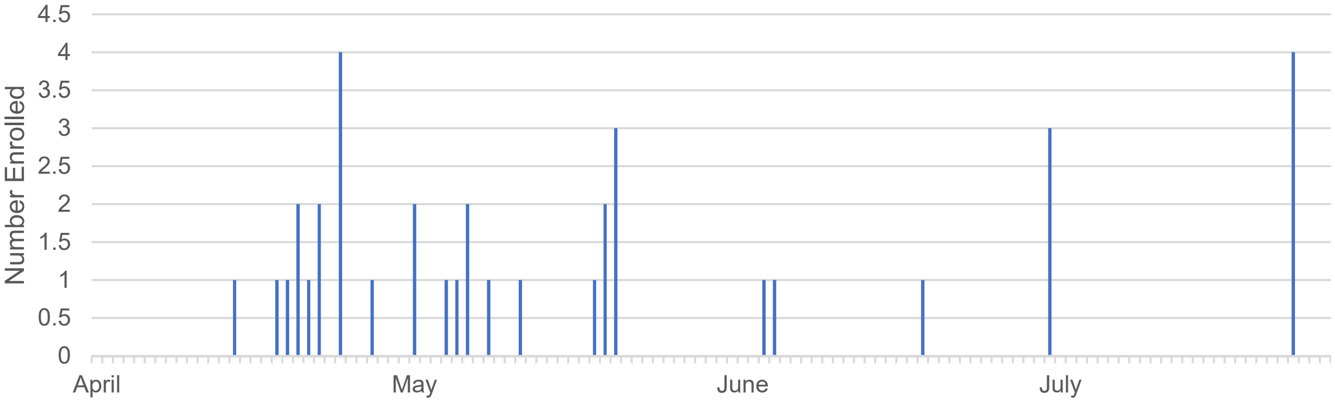

**Supplemental Figure 1. Study design, SARS-CoV-2 infection rate and study accrual**. A. Study design of PATCH sub-study 1 B. Infection Rate in the hospital system and in the region during study accrual. B. Accrual timeline to PACTH sub-study 1

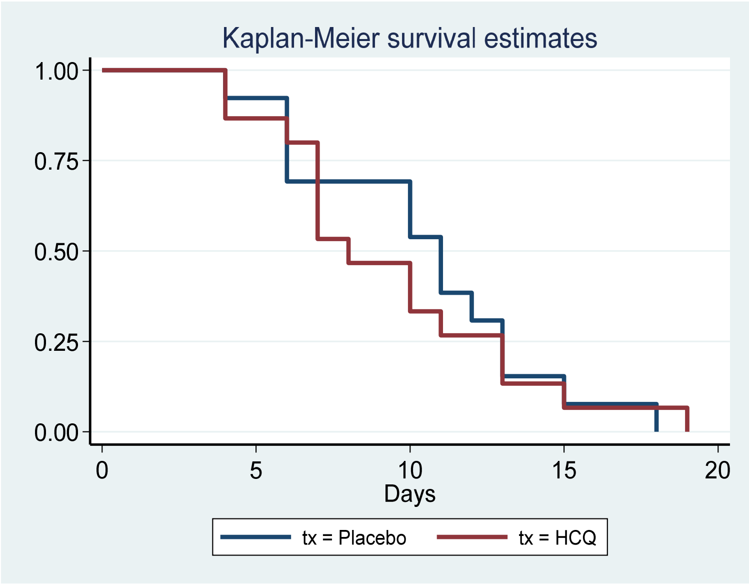


**Supplemental Fig. 2 Kaplan-Meier survival estimates for number days in quarantine for PATCH SS1 patients.**

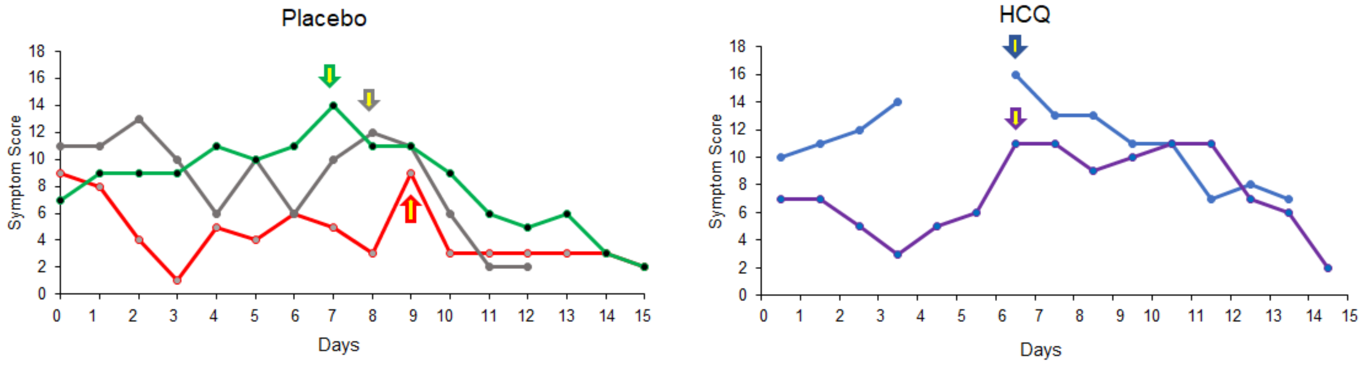


**Supplemental Figure 3. Patients who were unblinded due to progression of symptoms after being on study for 7 days.** The yellow arrows indicate the day that unblinding occurred. For subjects on placebo, all three crossed over to HCQ 400 mg bid on the day of unblinding.

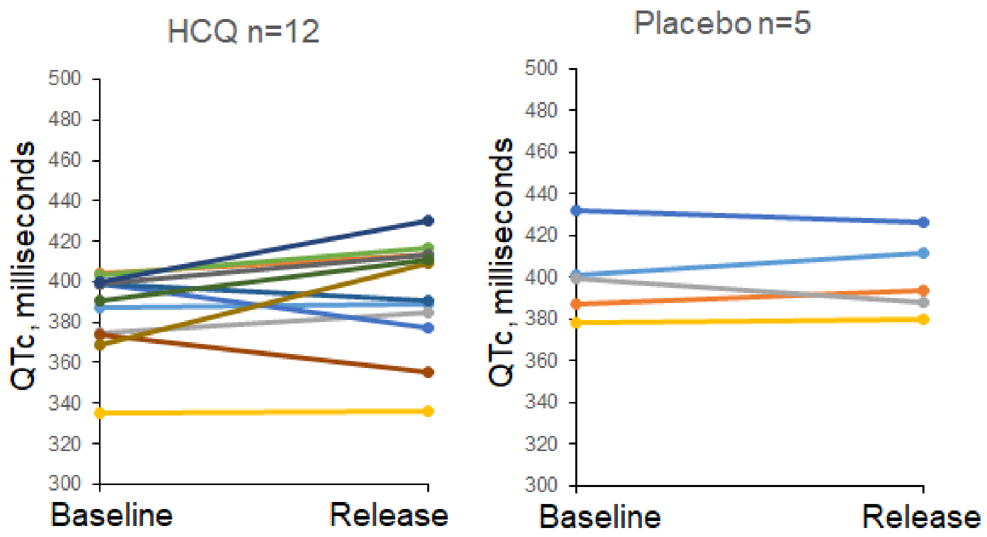


**Supplemental Figure 4. QTc at baseline and on day of release from quarantine for the 17 subjects that wore the ZioAT mobile telemetry device.**

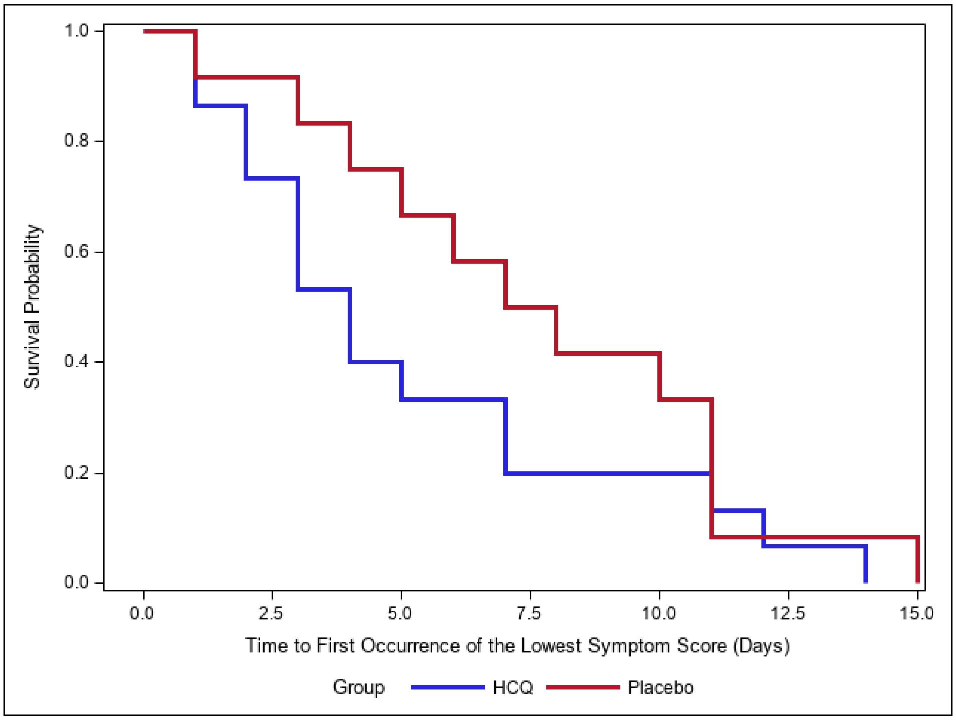


**Supplemental Figure 5. Time to the first occurrence of the lowest symptom score (days).**

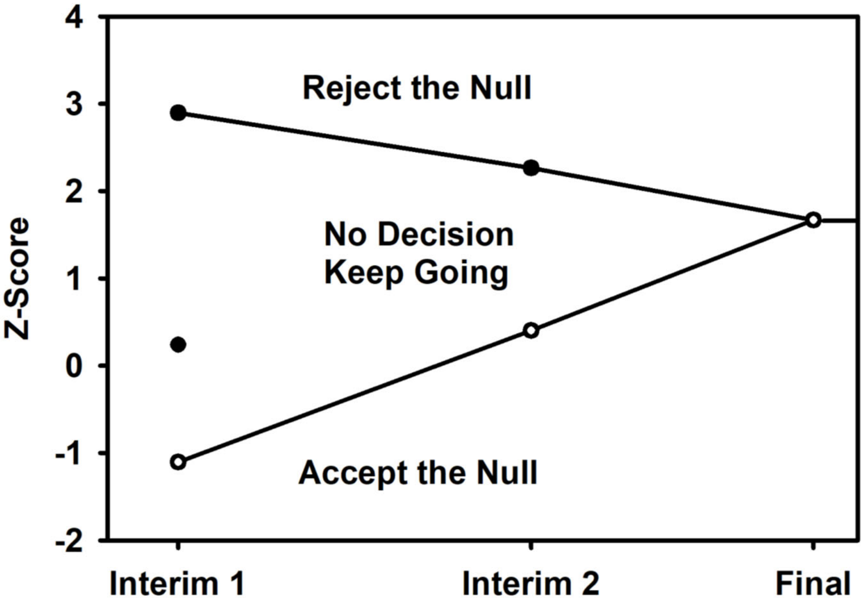


**Supplemental Figure 6. Graph of interim analyses and z scores that guide early stopping of this study.**

**Supplemental Tables 1-3**

*Abella et al. Hydroxychloroquine for treatment of SARS-COV-2 infections at home*

**Supplemental Table 1.** Example of Daily COVID-19 Symptom tracking score table

| Symptom | Mild =1 | Moderate =2 | Severe =3 | Score |
|---|---|---|---|---|
| Abdominal pain | | | | 0 |
| Fatigue | | | | 0 |
| Cough | | 2 | | 2 |
| Shortness of breath | 1 | | | 1 |
| Diarrhea | | | | 0 |
| Myalgias | | 2 | | 2 |
| Headache | | | | 0 |
| Smell disturbance | | | | 0 |
| Other: | | | | 0 |
| Total Score | | | | 5 |

**Supplemental Table 2. Baseline eligibility symptoms at time of testing and on day 0, by timepoint.**

| | HCQ | | Placebo | | |
|---|---|---|---|---|---|
| | n | No. (%) | n | No. (%) | p-value |
| At Time of Testing | | | | | |
| Fever | 17 | 12 (71) | 17 | 13 (76) | 1.000 |
| Cough | 17 | 11 (65) | 17 | 17 (100) | 0.018 |
| SOB | 17 | 7 (41) | 17 | 3 (18) | 0.259 |
| On Day 0 | | | | | |
| Fever | 17 | 5 (29) | 17 | 5 (29) | 1.000 |
| Cough | 17 | 10 (59) | 17 | 15 (88) | 0.118 |
| SOB | 17 | 5 (29) | 17 | 5 (29) | 1.000 |

**Supplemental Table 3. Comparison of eligibility symptoms at time of testing and on day 0, by treatment.**

| | Positive on Test Day | | Positive on Day 0 | | |
|---|---|---|---|---|---|
| | n | Percent | n | Percent | p-value |
| HCQ | | | | | |
| Fever | 17 | 12 (71) | 17 | 5 (29) | 0.016 |
| Cough | 17 | 11 (65) | 17 | 10 (59) | 1.000 |
| SOB | 17 | 7 (41) | 17 | 5 (29) | 0.909 |
| Placebo | | | | | |
| Fever | 17 | 13 (76) | 17 | 5 (29) | 0.008 |
| Cough | 17 | 17 (100) | 17 | 15 (88) | na |
| SOB | 17 | 3 (18) | 17 | 5 (29) | 0.500 |

**Supplemental Methods:**

*Cardiac monitoring with mobile cardiac telemetry:* When this study opened, we utilized the guidance from the American College of Rheumatology that baseline or serial EKg was not required to treat outpatients with HCQ. However, as 6 observational studies of COVID-19 revealed the possibility of QTc prologation and arrhythmia, we amended the protocol to include mobile cardiac telemetry. Participants were provided with a mobile cardiac telemetry device (ZioAT, iRhythm Technologies) to facilitate continuous single lead ECG monitoring. Baseline and daily QTcs and were assessed by the study team's cardiologist (MH) and iRhythm in a blinded fashion. iRhythm notified the study team if QTc prolongation to >500ms or sustained atrial or ventricular arrhythmias were observed. Refusal to wear the mobile cardiac telemetry device did not prevent study participation and/or did not result in removal from the trial.
