## Supplementary material for "Hydroxychloroquine for SARS-CoV-2 positive patients quarantined at home: The first interim analysis of a remotely conducted randomized clinical trial": PATCH protocol

**Supplement: Clinical Trial Protocol for the Prevention and Treatment of COVID-19 with hydroxychloroquine (PATCH trial).**

**Introduction:** The manuscript *Amaravadi et al.* reports the outcomes of Sub-Study 1 (SS1) of the PATCH trial protocol. It does not report on Sub-Study 3(SS3), a randomized trial of hydroxychloroquine versus placebo in health care workers, or Sub-Study 2 (SS2), a comparison of low-dose versus high-dose hydroxychloroquine in hospitalized patients with COVID-19. Sub-study 2 was closed upon recommendation of the University of Pennsylvania COVID-19 Clinical Trials Working Group. Sub-study was completed and the intervention was declared ineffective after the second interim analysis. The PATCH trial is closed to accrual. During the course of the study, there were 4 amendments to the protocol. The final protocol is contained in this supplement. The changes for each protocol amendment are summarized below.

**Original Protocol** Institutional Review Board (IRB) approval date: April 2, 2020

**Amendment 1 IRB approval date: April 22, 2020**

1.1 Cardiology involvement to monitor Qtc.
1.1a) Recruitment of cardiologists as sub-Investigators
1.1b) 4.1.1 Included home continuous telemetry monitoring with ZioAT (irhythm) for SS1
1.1c) 4.1.1 Have included a clause that states if baseline Qtc>500 once the ZIOAT is activated the patient comes off study and is sent to the hospital and replaced in the study.
1.1d) Included SS2 EKG and telemetry analysis by cardiology as available according to hospital guidelines which are rapidly adjusting to surge capacity.
1.1e) included optional mobile telemetry monitoring for patients on SS2.
1.1f) SS3 baseline EKGs and one month EKGs on health workers.
1.1g) We have included QTc prolongation as a criteria for dose modification.

1.2 Sub-study 1 Primary Objective Modification:
Clarification of definition of first date of quarantine.

1.3 Eligibility Modifications
1.3a). SS1: Changed “Fever and cough OR fever and SOB” to “Fever OR cough OR SOB”.
1.3b) Clarification that azithromycin not allowed at the time of enrollment
1.3c) SS1: Clarification patient must be symptomatic at time of enrollment
1.3d) SS1: Clarification: Must have access to working computer, smartphone and have internet access
1.3e) SS2: Patients who received up to 24 hours of standard of care HCQ are allowed to participate.
1.3f) SS3 clarification: HCW with symptoms of COVID-19 who tested negative for coronoavirus in the past 2 weeks are eligible.
1.3g) SS2 lab eligibility modifications: Removal of LFTs and coagulation eligibility requirements. 1.3h) Exclusion criteria: Clarification co-enrollment onto a interventional COVID-19 study is not allowed.
1.3i) Clarification: removal of the method of Qtc measurement. Justification: this requirement is too restrictive to ensure safety according to our new cardiology sib-investigators.

1.4 Participant recruitment and registration
1.4a) All of the details of testing that finds patients SS1 were removed.
1. 4b) SS1 Allows for paper consent if the Docusign process fails.
1.4c) Details of drug ordering and delivery have been clarified based on the most efficient process we have observed.
1.4d) Details of subject recruitment have been simplified to reflect on the ground conditions
1.4e) SS2 Paper consenting in case Docusign fails or subject has no cell phone.
1.4f) Process of recruitment and enrollment have been clarified to meet conditions on the ground.

1.5 Concomitant Meds, drug-drug interactions and Procedures
Website link to credible meds to search for Qtc prolonging drugs was included

1.6 Duration of Protocol treatment and follow-up.
Clarification of when HCQ will be stopped

1.7 Crossover
Clarification of the details of Crossover initiation and management are provided

1.8 Participant safety monitoring and data collection
1.8a) Mobile Cardiac Telemetry added to SS1 and SS2. This will be strongly recommended but not mandated for study participation. These kits with instructions will be provided to the patient and are self-administered.
1.8b) Data from these units will be routed to our cardiology Sub-Is who will be responsible for contacting the patient research and clinical team if arrhythmias are detected.
1.8c) Included patient collected nares swabs to assess viral titer in SS1
1.8d) Clarified on how to handle positive baseline and one month swab results for asymptomatic subjects on SS3.
1.8e) Included Qtc measurements on SS3.

1.9 Hydroxychloroquine Dose reduction.
Added Qtc criteria for dose holiday and reduction with HCQ.

1.10 Schedule of Events
Table 4 SS1
1. Clarified that HCQ dosing should not be delayed to collect the baseline oral fluid collection in the AM for SS1. Justification: for consistency of measurement morning collection is preferred.
2. Added mobile cardiac telemetry
3. Added nares swabs

Table 5 SS2
1. Added telemetry as clinically indicated
2. Added Mobile cardiac telemetry which is optional and self-administered
3. Clarified that COVID-19 specific labs were not study required.
4. Removed serum pregnancy test. Justification: the safety of HCQ in pregnancy is well established and COVID-19 is more deadly than HCQ side effects in early pregnancy.
5. A citrate blue top tube was added to the blood collection with an additional volume of 3 ml. This will be used for coagulation research. COVID-19 coagulopathy is becoming better understood and HCQ may have an impact on this phenomenon.

Table 6 SS3
1. Research EKG added

1.11 Research Specimen Process, banking and analysis
A new chart showing flow provided

1.12 Statistical Considerations
We included a new Bayseian approach to monitoring toxicity on SS2, that will allow us to terminate the high dose arm if there is a high rate of discontinuation of HCQ due to toxicity without concomitant efficacy.

**Amendment 2 IRB approval date: May 6, 2020**

2.1 Discontinuation of Sub-Study 2 (HCQ for the treatment of COVID-19 in hospitalized patients).
2.2 Clarification to Sub-Study 3 eligible population (to include “healthcare workers”).
2.3. FDA Safety Alert for Hydroxychloroquine and associated revised informed consent documents.
2.4. Clarification of language around mobile telemetry on SS1
2.5. New participant facing materials to facilitate study operations and re-consent from Amendment 1

**Amendment 3 IRB approval date May 24, 2020**

3.1 SS1mobile telemetry clarification
3.2 SS1 Change from 7 to 10 days quarantine release per CDC guidelines
3.3 SS1 statistical analysis plan changed to specify how this will be analyzed
3.4 HCQ dose reduction limited to one dose lebel
3.5 Added Sandoz required language for reporting misuse of drug
3.6 Clarification of symptom scoring table for SS1

**Amendment 4 IRB approval date: June 23, 2020**

4.1 establishment of a Data Safety and Monitoring Board (slide 29)
4.2 SS3 Secondary objective # shifts missed removed as this measurement is not useful
4.3 Removed some Sub-Is.
4.4 Clarification of evaluable patients section 10 excludes SS3 PCR+ subjects at baseline from primary outcome.
4.5 clarification that subjects that withdraw from treatment can complete other study procedures.
4.6 clarification that subjects must agree to self report pregnancy during study and 90 days post. This is in accordance with the current consent form.

**The PATCH (Prevention And Treatment of COVID-19 with HCQ) Trial:**
**A 2 Arm Randomized Trial of Hydroxychloroquine in the Prevention and Treatment of COVID-19**

| | |
|---|---|
| Sponsor-Investigator/<br>Principal Investigator: | Ravi Amaravadi, MD (Medicine)<br>University of Pennsylvania Perelman School of Medicine<br>Department of Medicine, Division of Hematology Oncology<br>852 BRB 2/3 421 Curie Blvd,<br>Philadelphia, PA 19104<br> |
| Primary Emergency Medicine Sub-Investigator | Benjamin Abella, MD (Emergency Medicine)<br>University of Pennsylvania Perelman School of Medicine<br>Department of Emergency Medicine<br>Hospital of the University of Pennsylvania<br>Emergency Department, Ground Ravdin<br>3400 Spruce Street<br>Philadelphia, PA 19104<br> |
| Primary Infectious Diseases Sub-Investigator | Ian Frank, MD<br>502 Johnson Pavilion<br>3610 Hamilton Walk 6073<br>Philadelphia, PA 19104-6073<br> |
| Other Significant<br>Sub-Investigators | Anish Agarwal, MD (EM)<br>Julie Uspal, MD (EM)<br>Ari Friedman, MD (EM)<br>John Greenwood, MD (EM)<br>Felipe Teran, MD (EM)<br>Michael Milone, MD PhD (Pathology)<br>William Short, MD (ID)<br>E. Paul Wileyto (Statistics)<br>Phyllis Gimotty, PhD (Statistics)<br>Joseph Carver, MD (Cardiology)<br>Matthew Hyman, MD, PhD<br>(Cardiology)<br>Dasha Babushok, MD (Oncology)<br>Saar Gill, MD (Oncology)<br>IvanMaillard Md PhD (Oncology)<br>Alexander Huang, MD (Oncology)<br>Dan Vogl, MD (Oncology) |
| Medical Monitor: | Sunita Nasta, MD<br>University of Pennsylvania Perelman School of Medicine<br>Department of Medicine, Division of Hematology Oncology<br><br>215-605-2493 |

| | |
|---|---|
| Funding: | Internal Funding |
| Protocol Number: | IRB: 842838 |
| Clinical Trial Management System | Penn-CTMS (Velos) |
| Investigational Agents: | Hydroxychloroquine/Placebo |
| IND #: | IND exempt (Granting entity University of Pennsylvania, Office of Clinical Research (OCR)) |
| Version: | 6.17.2020 |

**PRINCIPAL INVESTIGATOR SIGNATURE**

| | |
|---|---|
| SPONSOR-INVESTIGATOR: | Ravi Amaravadi, MD |
| STUDY TITLE: | The PATCH (Prevention And Treatment of COVID-19 with HCQ) Trial: A 2 Arm Randomized Trial of Hydroxychloroquine in the Prevention and Treatment of COVID-19 |
| STUDY ID | IRB #842838 |
| PROTOCOL VERSION | Amendment 4 (6.17.2020) |

I have read the referenced protocol. I agree to conduct the study in accordance to this protocol, in compliance with the Declaration of Helsinki, Good Clinical Practices (GCP), and all applicable regulatory requirements and guidelines.

| | |
|---|---|
| Ravi Amaravadi, MD | Ravi Amaravadi |
| Principal Investigator Name | Signature |
| University of Pennsylvania | 4/2/20 |
| Affiliation | Date |

**Abbreviations:**

Ab: antibody
AE: adverse event
ALT: alanine aminotransferase
ANC: absolute neutrophil count
AST: aspartate aminotransferases
BID: twice daily
BP: blood pressure
BSA: body surface area
CNS: central nervous system
CT: computed tomography
CTCAE: Common Terminology Criteria for Adverse Events
DSMC: Data Safety and Monitoring Committee
eCRF: electronic case report form
EKG: electrocardiogram
FDA: Food and Drug Administration
FFPE: formalin fixed-paraffin embedded
HBV: hepatitis B virus
HCQ: hydroxychloroquine
HCV: hepatitis C virus
HUP: Hospital of the University of Pennsylvania
IHC: immunohistochemistry
IND: investigational new drug
INR: international normalization ratio
IRB: institutional review board
IV: intravenous
LLN: lower limit of normal
LVEF: left ventricular ejection fraction
PPMC: Penn-Presbyterian MedicalCenter
PPT1: palmitoyl-protein thioesterase 1
PR: partial response
PT: prothrombin time
PTT: partial thromboplastin time
SAE: serious adverse event
SUSAR: Suspected, Unexpected Serious Adverse Reaction
ULN: upper limit of normal
WBC: white blood cell

**PRINCIPAL INVESTIGATOR SIGNATURE** ........................................ 3

**1.0 OBJECTIVES** ........................................ 11
1.1 PRIMARY OBJECTIVE ........................................ ERROR! BOOKMARK NOTDEFINED.
1.2 SECONDARY OBJECTIVES ........................................ ERROR! BOOKMARK NOTDEFINED.
1.3 CORRELATIVE OBJECTIVES ........................................ ERROR! BOOKMARK NOTDEFINED.
11.4 UPENN ABRAMSON CANCER CENTER (ACC)'S DATA AND SAFETY MONITORING COMMITTEE (DSMC) NOTIFICATION BY ALL SITES ........................................ ERROR! BOOKMARK NOT DEFINED.

Study Summary

| | |
|---|---|
| **Title** | The PATCH (Prevention And Treatment of COVID-19 with HCQ) Trial: A 2 Arm Randomized Trial of Hydroxychloroquine in the Prevention and Treatment of COVID-19 |
| **Phase** | Phase II |
| **Methodology** | A prevention and treatment trial with two sub-studies including randomized double-blind placebo controlled sub-study design. |
| **Study Duration** | 1 year |
| **Study Center(s)** | Single Institution: University of Pennsylvania (HUP and PPMC) |
| **Objectives** | **1.1 <u>PRIMARY OBJECTIVES</u>**<br>Sub-Study 1 (home quarantined COVID-19 patients): Median time to release from quarantine by meeting the following criteria: 1) No fever for 72 hours without the use of fever-reducing medications 2) improvement in other symptoms and 3) 7 or 10 days have elapsed since the beginning of symptom onset, as determined by CDC guidelines at the time. The first date of quarantine will be considered the date of SARS-CoV-2 nasopharyngeal swab test that confirmed positivity.<br><br>Sub-Study 3 Healthcare worker prophylaxis: Rate of COVID-19 infection at 2 months<br><br>**1.2 <u>SECONDARY OBJECTIVES</u>**<br>Sub-Study 1: Rate of participant-reported secondary infection of housemates, rate of hospitalization, rate of treatment related adverse events<br>Sub-Study 3:rate of treatment related adverse events<br><br>**1.3 <u>CORRELATIVE OBJECTIVE</u>**<br>To bank serially collected samples to enable correlative science related this trial |
| **Number of Subjects** | Sub-Study 1: 100; interim analysis after 34 and 68<br>Sub-Study 3: 200; interim analysis after 50 and 100<br>**Total: 300** |
| **Diagnosis and Main Inclusion/Exclusion Criteria** | • Age ≥ 18 years old (Sub-study 3)<br>• Competent and capable to provide informed consent |

- Subjects meeting the following criteria by Sub-Study

Sub-Study 1:

- Age ≥40 years since the risk of prolonged disease that progresses to severe COVID-19 disease increases with age.
- PCR-positive for the SARS-CoV2 virus
- (Fever or cough, or shortness of breath,
- ≤4 days since the first symptoms of COVID-19 and date of testing
- Not taking azithromycin at the time of enrollment
- Symptomatic at the time of enrollment
- Not requiring hospitalization and is sent home for quarantine.
- Must live within 30 miles of HUP or Penn Presbyterian Medical Center to facilitate drop-off of medication
- Must own a working computer, or smartphone and have internet access
- Must be willing to fill out a daily symptom diary
- Must be available for a daily phone call,
- Must take their own temperature twice a day
- Must be willing to report the observed symptoms and development of COVID-19 in the co-inhabitants of the residence at which the quarantine will be served.

Sub-Study 3 Healthcare Worker Prevention

- Healthcare worker at HUP or PPMC
- ≥20 hours per week of clinical work scheduled in the coming 2 months during the COVID-19 pandemic
- No fever, cough, or shortness of breath in the past 2 weeks unless a SARS-CoV-2 test was performed for these symptoms and the result was negative prior to enrollment

- Willing to report compliance with HCQ in the form of a diary
- Patients must be able to swallow and retain oral medication and must not have any clinically significant gastrointestinal abnormalities that may alter absorption such as malabsorption syndrome or major resection of the stomach or bowels.

**Exclusion Criteria**

- <18 years of age
- Prisoners or other detained persons
- Allergy to hydroxychloroquine

| | |
|---|---|
|  | • Pregnant or lactating or positive pregnancy test during pre-medication examination<br>• Receiving any treatment drug for SARS-CoV-2 within 14 days prior to screening evaluation (off label, compassionate use or trial related).<br>• Co-enrollment onto another interventional COVID-19 study is not allowed.<br>• Known history of retinal disease including but not limited to age related macular degeneration.<br>• Taking any of the following medications that prolong Qtc: Chlorpromazine, Haloperidol, Droperidol, Quetiapine, Olanzapine, Amisulpride, Thioridazine<br>• History of interstitial lung disease or chronic pneumonitis unrelated COVID-19.<br>• Due to risk of disease exacerbation patients with porphyria or psoriasis are ineligible unless the disease is well controlled and they are under the care of a specialist for the disorder who agrees to monitor the patient for exacerbations.<br>• Patients with serious intercurrent illness that requires active infusional therapy, intense monitoring, or frequent dose adjustments for medication including but not limited to infectious disease, cancer, autoimmune disease, cardiovascular disease.<br>• Patients who have undergone major abdominal, thoracic, spine or CNS surgery in the last 2 months, or plan to undergo surgery during study participation.<br>• Patients receiving cytochrome P450 enzyme-inducing anticonvulsant drugs (i.e. phenytoin, carbamazepine, Phenobarbital, primidone or oxcarbazepine) within 4 weeks of the start of the study treatment<br>• History or evidence of increased cardiovascular risk including any of the following:<br> • Left ventricular ejection fraction (LVEF) < institutional lower limit of normal. Baseline echocardiogram is notrequired.<br> • Current clinically significant uncontrolled arrhythmias. Exception: Subjects with controlled atrial fibrillation<br> • History of acutecoronary syndromes (including myocardial infarction and unstable angina), coronary angioplasty, or stenting within 6 months prior to enrollment<br> • Current ≥ Class II congestive heart failure as defined by New York Heart Association |
| **Study Product, Dose, Route, Regimen** | Hydroxychloroquine, various doses, oral<br>Placebo, oral |

| | |
|---|---|
| **Duration of administration** | Sub-Study 1 at home quarantine<br>Arm 1: HCQ 2 weeks 400 mg bid; Arm 2: Placebo 2 pills bid for 2 weeks; crossover allowed upon disease progression or lack of clearance of virus<br>Sub-Study 3 Healthcare worker prophylaxis. Arm 1: HCQ 600 mg qd for 2 months versus Arm 2: Placebo 3 pills qd for 2 months; crossover from placebo to HCQ is allowed upon disease progression/ virus confirmation |
| **Study design** | 1:1 randomization in each cohort between the arms 1 and 2 |
| **Duration of trial** | Approximately 1 year |

#### 2.0 OBJECTIVES

##### 2.1 PRIMARY OBJECTIVES

Sub-Study 1 (home quarantined COVID-19 patients): Median time to release from quarantine by meeting the following criteria: 1) No fever for 72 hours without the use of fever-reducing medication 2) improvement in other symptoms and 3) 7 or 10 days have elapsed since the beginning of symptomonset, depending on CDC guidelines at the time. (Outcomes based on 7 and 10 days will be constructed for each subject.)
The first date of quarantine will be considered the date of SARS-CoV-2 nasopharyngeal swab test that confirmed positivity.
Sub-Study 3 Healthcare worker prophylaxis: Rate of COVID-19 infection at 2 months

##### 1.2 SECONDARY OBJECTIVES

Sub-Study 1: Rate of participant-reported secondary infection of housemates, rate of hospitalization, rate of treatment related adverse events
Sub-Study 3: rate of treatment related adverse events

##### 1.4 CORRELATIVE OBJECTIVE

To bank serially collected samples to enable correlative science related this trial

#### 2 BACKGROUND AND RATIONALE

**2.1 Chloroquine derivatives show preclinical efficacy against coronavirus but very little clinical data is available.** Emerging viral diseases (EVDs) encompass a growing list of zoonotic viruses that have a major impact on global health and economics. These include Ebola, SARS, MERs, Marburg, and the recently identified virus that causes COVID-19 disease (Wuhan Coronavirus: SARS-CoV-2) (1). In each of these cases there is currently no effective drug treatments or prophylaxis agents that can be quickly applied to large populations at risk for the virus. As of March 2020, The SARS-CoV2 virus has infected more than 180,000 people causing more than 5000 deaths. This has led to a lockdown of entire megacities in China, international travel bans, and disruptions in global supply chains. More alarming is the recent evidence that health care workers are getting infected and dying of the virus, wreaking havoc on the chain of care. In addition, while prior epidemics such as Ebola seemed to be contained in Africa, the current COVID-19 global crisis demonstrates how with the extent of globalization and international travel, no country is safe from these EVDs.

Coronaviruses are a large family of viruses that commonly infect many animals, including camels, cattle, cats, and are often found in bats as their zoonotic reservoir. While it is rare, animal coronaviruses can infect people and then spread between people, as had occurred with Middle East respiratory coronavirus (MERS-CoV) and severe acute respiratory coronavirus (SARS-CoV), and now the newly emerged coronavirus SARS-CoV-2 (also known as COVID-19). The SARS-CoV-2 virus is a betacoronavirus, like MERS-CoV and SARS-CoV. All three of these viruses have their origins in bats. While SARS-CoV and MERS-CoV exhibit significantly higher mortality than SARS-CoV-2, the ability to spread between humans is less than SARS-CoV-2. There is emerging evidence from Asia that patients can be highly contagious with one patient spreading viral particles to 87% of 15 sites within the patient's room. Virus can be shed in urine and feces as well as respiratory secretions increasing the likelihood that sick contacts and hospital staff will become infected (2). An epidemiological survey of 72,000 cases in China found and overall 3.8% infection rate in hospital workers, but a 68% infection rate of hospital workers in Wuhan at the epicenter of disease. This suggests that if low rates of infection are in a community, the likelihood of hospital workers contracting disease is low, but if there are a large number of cases then the likelihood of hospital workers contracting the disease is high (3). Patients above the age of 50 are more likely to die of this disease and the time from diagnosis to death can occur within one month despite access to high- level intensive care units (4). A high rate of intra-family transmission and rapid community transmission has been documented in Shenzen China (5). Meanwhile a screen of compounds found that chloroquine prevented and irradicated established infection of SARS-CoV-2 virus in vitro. The Chinese and Korean consensus guidelines for COVID-19 treatment include chloroquine. However, based on our experience in cancer clinical trials we believe hydroxychloroquine is a safer drug than chloroquine and affords the ability to dose escalate to concentrations that we know are effective at blocking the lysosome in patients. A recent study found that hydroxychloroquine produced rapid clearance of virus in hospitalized patients with COVID-19 (Gautret et al Int. J Antimicrobial Agents 2020 in press).

Our ***hypothesis*** is that high doses of hydroxychloroquine for at least 2 weeks can be effective antiviral medication both as a treatment in hospitalized and ambulatory patients and prophylaxis in health care workers because it impairs lysosomal function and reorganizes lipid raft (cholesterol and sphingolipid rich microdomains in the plasma membrane) content in cells, which are both critical determinants of EVD infection. This hypothesis is based on a growing literature linking chloroquine to antiviral activity and our own work in the field. We believe there is enough information to launch a clinical trial at the Hospital of the University of Pennsylvania of hydroxychloroquine for COVID-19

***2.2 Mechanistic rationale for antiviral properties of chloroquine derivatives.***

**2.2a Coronaviruses.** Coronaviruses (CoV) infect mammals and birds causing respiratory, gastrointestinal, and central nervous system diseases. Coronavirus virions contain an envelope, a helical capsid, and a single-stranded RNA genome, the largest among all RNA viruses. The name "coronaviruses" derives from the spike proteins on their envelope that give the virions a crown-like shape. CoVs include the following: 1) α-genus: human coronavirus NL63 (HCoV-NL63), porcine transmissible gastroenteritis coronavirus (TGEV), and porcine respiratory coronavirus (PRCV) 2) β-genus: SARS-CoV-2/COVID-19, severe acute respiratory syndrome coronavirus (SARS-CoV), Middle East respiratory syndrome coronavirus (MERS-CoV), mouse hepatitis coronavirus (MHV), and bovine coronavirus (BCoV). 3) CoVs of the γ-genus include avian infectious bronchitis virus (IBV) and do not infect humans so are valuable laboratory agents (6). Coronaviruses impose health threats to humans and animals. SARS-CoV caused the SARS epidemic in 2002 to 2003, with over 8,000 infections and a fatality rate of ~10%. MERS-CoV emerged from the Middle East in 2012 causing 877 infections with a fatality rate of ~36%. HCoV-NL63 from the α-genus is a widespread pathogen that produces the common cold in healthy adults and acute respiratory illness in young children. CoVs are also major animal pathogens. TGEV and MHV cause close to 100% fatality in young pigs and young mice, respectively; BCoV and IBV also cause significant healthcare burden for domesticated cattle and chickens, respectively. Therefore, research on coronaviruses has strong health and economic implications (6).

**2.2b CoV infection requires functional lysosomes.** Receptor recognition by viruses is the first and essential step of viral infections of host cells. An envelope-anchored spike protein mediates coronavirus entry into host cells by first binding to a receptor on the host cell surface and then fusing viral and host membranes. CoVs recognize a number of different host receptors. Once the receptor is bound, there is viral and cell membrane fusion, followed by endocytosis and lysosomal processing. CoVs like SARS enter the cell through lipid raft enriched endocytosis (7). The details of the interaction between the virus spike protein, virus membrane and the lipid rafts are not worked out. In the lipid raft literature, it is often the case that lipid rafts (semisolid phase membrane filled with cholesterol and sphingolipids) are only physiologically functional if there is a non-lipid raft membrane region next to it. After membrane fusion, endosomal pH acidification is a fusion trigger for CoVs and other viruses including influenza virus and vesicular stomatitis virus (VSV). For instance the use of lysosomal inhibitor blocked the entry of the model coronavirus IBV (8). The authors directly assessed the pH dependence of IBV fusion and found that fusion only occurs at acidic pH. For some CoVs that harbor a non-cleaved spike protein on their surface, such as MHV-2 and SARS-CoV, it has been shown that they rely on lysosomal proteases for productive entry. In fact one study found that bat cells have more efficient lysosomal proteolysis than human cells providing an explanation for why bats are preferred zoonotic hosts for CoVs (9).

**2.2c Evidence that chloroquine prevents coronavirus infection.** It has been reported that chloroquine has strong antiviral effects on SARS-CoV infection and spread in vitro (10-12). In to increasing endosomal/lysosomal pH, these studies demonstrated that CQ abrogates glycosylation of ACE2, the cellular receptor of SARS-CoV, which may contribute further to infection suppression. The IC50 of chloroquine for inhibition of SARS-CoV in vitro is 8.8 micromolar which would require higher concentrations than typically delivered in malaria. Importantly, the suppressive effects are observed when the cells are treated with chloroquine either before or after exposure to the virus, suggesting both prophylactic and therapeutic treatment paradigms could be employed (10, 11). The work by Kayaerts looked at human coronavirus OC43 which causes neonatal death in mice. Treatment with chloroquine of pregnant mothers provided a 98.6% protection against death in newborn mice. This is in the only in vivo demonstration of chloroquine efficacy against a CoV. DeWilde et al. screened a library of 348 FDA-approved drugs for anti- MERS-CoV activity in cell culture and only four compounds (chloroquine, chlorpromazine, loperamide, and lopinavir) were found to inhibit the viral replication (50% effective concentrations, EC50 3–8 Imol/L) (12). The protective activity of chloroquine against CoVs such as SARS, MERS and COVID-19 has not yet been established in animal models.

**2.2d Chloroquine is active against COVID-19/ SARS-CoV-2:** A recent paper from Wuhan China isolated SARS-CoV2 (the virus that causes COVID-19 disease) and tested 6 modern antiviral drugs and chloroquine and found that the best suppressors of viral infection were remdesivir (IC50 0.77 μmol) and chloroquine (IC50 1 μmol) (13). Only chloroquine given at 10 μM was able to suppress infection either with pretreatment, at the time of infection, or after infection occurred. This has led to clinical trials and off label use of chloroquine in China (Dr. Amaravadi personal communication with scientists in China). There are Chinese guidelines for the use of chloroquine 500 mg bid for the treatment of COVID-19.

**2.2e Evidence for immunopathology in severe CoVs infection**. Although the mechanisms of pathology in severe SARS and MERS CoV infections are not well understood, the severe lung damage observed in SARS patients is associated with high viral loads during the early phase of infection and abundant macrophage-monocyte and neutrophil accumulation in the lungs (14). Dysregulated innate and adaptive immunity have been postulated to contribute to severe CoV disease. The acute lung injury of severe CoV infection is associated with elevated serum pro-inflammatory cytokines. In particular, fatal SARS is associated with a high and persistent expression of interferon (IFN) and IFN-responsive genes with evidence for impaired adaptive immunity, most notably reduced anti-Spike neutralizing antibody responses (15). In a murine model of SARS, disruption of type I IFN signaling or monocyte-macrophage depletion protected mice from lethal infection supporting a role for dysregulated type I IFN expression in CoV pathology (16).

**2.2 f Chloroquine modulates type I interferon expression and T cell immune responses**. The endosomal compartment plays a central role in immunobiology, especially in innate pathogen sensing as well as antigen presentation. Toll-like receptors that recognize virally-associated nucleic acid are localized to the endosome of monocytes and specialized DC populations including the plasmacytoid DC (pDC), which are a major producer of type I interferons (17). HCQ is able to disrupt this endosomal sensing pathway to reduce type I interferons production by these cells, and may attenuate excessive type I interferon expression that has been postulated to contribute to severe CoV infection. HCQ has also been reported to modulate endosomal membrane permeability to enhance cross-presentation of endosomal antigens via MHC class I, which enhances adaptive CD8+ T cell immunity.

**2.3 Clinical experience with high dose hydroxychloroquine in cancer patients.** In contrast to CQ, which can produce blindness at high cumulative doses, our recent body of work has demonstrated that high dose HCQ can be administered safely to humans for months. Our work on autophagy as a resistance mechanism to cancer therapy had identified CQ derivatives as potential anti-cancer agents (18). Since HCQ has had a longer track record of dose escalation and chronic dosing in rheumatoid arthritis and lupus, we chose HCQ as our lysosomal autophagy inhibitor to test in combination regimens in cancer patients. We reported the first 6 phase I dose escalation clinical trials involving HCQ in combination with FDA approved anti-cancer drugs in refractory stage IV cancer patients (19-24). In most patients on these trials we were able to escalate the dose of HCQ to 1200 mg per day and dose patients in some cases safely for more than one year. In over 220 patients treated across multiple studies the rate of grade 3-4 non-hematological toxicities was < 10%. Most of these had more to do with the severity of the cancer and would likely not be seen in a healthy population. Specifically, no retinal, cardiac, neurological, hepatic or renal toxicity was observed. Hematological toxicities could be attributed to the other cancer drug (chemotherapy) that HCQ was paired with, and the most common side effects included manageable gastrointestinal symptoms such as bloating, diarrhea, constipation and mild non-bloodydiarrhea.

**2.4 Low dose versus high dose HCQ.** In HCQ studies in cancer patients we conducted population pharmacokinetic studies and pharmacodynamic studies and determined that 800-1200 mg daily was required to effectively and consistently impair the lysosome in peripheral blood mononuclear cells. Our PK studies determined that it took roughly 2 weeks to achieve steady state in cancer patients. For these reasons there is rationale to propose a high dose (600 mg po bid) prolonged schedule treatment (2 weeks). In contrast an in vitro study recently published which included PK-PD modeling extrapolated from published PK studies indicated that HCQ was very active against COVID-19 but only 400 mg po bid X1 followed by 200 mg po bid for 5-10 days was enough for treatment. A recent non-randomized open label French trial (Gautret et al Int. J Antimicrobial Agents 2020) showed that 600 mg qd of HCQ cleared virus in 70 % of mildly ill hospitalized patients compared to 12.5% at 6 days of control patients. This generates a major question in how to use this agent in different populations that can best be answered in this three-part placebo controlled randomizedtrial.

#### 3.0 ELIGIBILITY

##### 3.1 Inclusion Criteria

3.1.1 Age ≥ 18 years old, except sub-study 1
3.1.2 Competent and capable to sign informed consent
3.1.3 Subjects meeting the following criteria by arm:
3.1.3 Sub-Study 1:

- Age ≥40 years since the risk of prolonged disease that progresses to severe COVID-19 disease increases with age.
- PCR-positive for the SARS-CoV2 virus
- Fever or cough or shortness of breath
- ≤4 days since the first symptoms of COVID-19 and date of testing
- Not requiring hospitalization and is sent home for quarantine.
- Not taking azithromycin at the time of enrollment
- Symptomatic at the time of enrollment
- Must live within 30 miles of HUP or Penn Presbyterian Medical Center to facilitate drop-off of medication
- Must have access to a working computer, or smartphone and have internet access
- Must be willing to fill out a daily symptom diary
- Must be available for a daily phone call,
- Must take their own temperature twice a day
- Must be willing to report the observed symptoms and development of COVID-19 in the co-inhabitants of the residence at which the quarantine will be served.

Sub-Study 3 Health Care Worker Prevention

- Healthcare worker at HUP or PPMC (Healthcare worker is defined as: a UPHS employee that physically interact with COVID-19 patients e.g., doctors, nurses, respiratory techs, certified nursing assistants, emergency room techs, OB ultrasound techs).
- ≥20 hours per week of clinical work scheduled in the coming 2 months during the COVID-19 pandemic
- No fever, cough, or shortness of breath in the past 2 weeks unless a SARS-CoV-2 testwas performed for these symptoms and the result was negative prior toenrollment

3.1.4 Sub-studies 1 and 3: Willing to report compliance with HCQ in the form of a diary
3.1.5 Sub-Studies 1 and 3: Participants are willing to collect oral fluid using a simple applicator that is provided, and freeze these samples.
3.1.6 Not pregnant and/or breastfeeding, as determined below:

Sub-Study 1 and 3: Not known to be pregnant or breast feeding as determined through self-report

[All participants] 3.1.7 Patients must be able to swallow and retain oral medication and must not have any clinically significant gastrointestinal abnormalities that may alter absorption such as, but not limited to malabsorption syndrome, major resection of the stomach or bowels, gastric bypasss, lap banding.

**3.2 Exclusion Criteria**

3.2.1 <18 years of age [Sub-Study 3]; <40 years old [Sub-Study 1]
3.2.2 Prisoner or other detained person
3.2.3 Allergy to hydroxychloroquine, 4 aminoquinolines, or quinine
3.2.2 Patients with known history of G6PD deficiency
3.2.2 Pregnant and/or breastfeeding, see above
3.2.3 Receiving any treatment drug for 2019-ncov within 14 days prior to screening evaluation (off label, compassionate use or trial related).
3.2.4 Co-enrollment onto another interventional COVID-19 study is not allowed.
3.2.5 Known history of retinal disease, including but not limited to, macular degeneration, retinal vein occlusion, visual field defect, diabetic retinopathy
3.2.6 Known history of interstitial lung disease, severe emphysema or asthma, or chronic pneumonitis unrelated COVID-19.
3.2.7 Taking any of the following medications that prolong Qtc: Chlorpromazine, Haloperidol, Droperidol, Quetiapine, Olanzapine, Amisulpride, Thioridazine
3.2.8 Due to risk of disease exacerbation patients with known porphyria or psoriasis are ineligible unless the disease is well controlled and they are under the care of a specialist for the disorder who agrees to monitor the patient for exacerbations.
3.2.6 Patients with serious intercurrent illness that requires active infusional therapy, intense monitoring, or frequent dose adjustments for medication including but not limited to infectious disease, cancer, autoimmune disease, cardiovascular disease.
3.2.7 Patients who have undergone major abdominal, thoracic, spine or CNS surgery in the last 2 months, or plan to undergo surgery during study participation.
3.2.8 Patients receiving cytochrome P450 enzyme-inducing anticonvulsant drugs (i.e. phenytoin, carbamazepine, Phenobarbital, primidone or oxcarbazepine) within 4 weeks of the start of the study treatment
3.2.9 History or evidence of increased cardiovascular risk including any of the following:

- Left ventricular ejection fraction (LVEF) < institutional lower limit of normal. Baseline echocardiogram is not required.
- Current clinically significant uncontrolled arrhythmias. Exception: Participants with controlled atrial fibrillation
- History of acute coronary syndromes (including myocardial infarction and unstable angina), coronary angioplasty, or stenting within 6 months prior to enrollment
- Current ≥ Class II congestive heart failure as defined by New York HeartAssociation

### 4.0 PARTICIPANT RECRUITMENT, CONSENT, AND REGISTRATION

#### 4.1 Sub-study –specific flow of recruitment, consent, registration, drug supply

4.1.1 Sub-Study 1 Outpatient Recruitment, Consent and Registration Procedure:

1. Patients with a positive SARS-CoV-2 test are identified by a clinical lab.
   For this sub-study, patients ≥40 with Fever or cough, or
   shortness of breath AND symptoms started 4 days or less before the patient came for testing
2. Delegated study team member will be notified of positive patients (as example: clinical research coordinators (CRC)).
3. Study team member screens the EPIC chart for specific eligibility criteria and calls the patient (if appropriate)
   a. Informs the patient of positive SARS-CoV-2 result
   b. Introduces the study and determines if the patient is interested in hearing more details from a clinician investigator
   c. If interested, study team member sends consent via email/DocuSign to interested patient
4. Clinical Investigator contacts the interested patient
   a. Conducts e-consent process with the patient and documents via telemedicine/DocuSign
   b. If the DocuSign process fails due to technical difficulties, and the participant can print out the pdf, sign the consent form, take a digital photo and send the study, the consenting process can proceed. The clinical investigator must sign, date and time the printed signature page with the patient signature, and document the consenting process in EPIC.
   c. Screens the participant for signs and symptoms that would warrant hospitalization (dizziness, shortness of breath at rest, orthostasis, low urine output).
   d. Completes the eligibility checklist and signs it.
5. CRC links EPIC chart to study and emails IDS for randomization
6. IDS randomizes the participant Clinical Investigator uses PennChart to order the study drug IDS prepares blinded medication in bottles
7. Blinded study drug (HCQ or placebo) are dispensed to delegated study member and delivered to the participant's house or shipped. Study team includes oral fluid collection kits, nares swabs, mobile cardiac telemetry monitoring device (ZioAT, iRhythm Technologies). The patient must call the CRC to inform receipt of study drug. CRC documents this receipt. If for some reason this is not possible, drug will be shipped to the participant using an approved pharmacy shipping vendor.
8. Appropriately delegated and credentialed study team member calls the participant and reviews use of the study drug, establishes best contact information for response monitoring (See Section 8.5), and asks the patient to connect/wear the cardiac telemetrymonitoring device. If the baseline, adjusted (Pre-treatment) Qtc is >500, the patient will be removed from the study and advised to go to the emergency room for admission to the hospital for evaluation for inpatient monitoring. Since the half life of HCQ is many days, the activation of the ZioAT and transmission of the baseline QTc does not need to occur before starting study medication.

4.1.2 Sub-Study 3 Outpatient Hospital Workers

1. Healthcare workers will enroll in stations in the ED.
2. Delegated and credentialed clinical investigator sends consent via email/DocuSign and reviews consent form with the participant
3. Conducts e-consent process with the patient and documents via telemedicine/DocuSign
4. If a participant is a member of the PATCH study team, then they are counselled to ensure they do not reveal their own participation during the consenting of participants to Sub-study 1.
5. Eligibility checklist is reviewed and signed by an appropriately delegated and credentialed clinical investigator

6. Participant is randomized by IDS
7. Sub-Investigator orders study drug and PCR test.
8. Nasopharyngeal swab for COVID19, and blood collection if available/possible is performed in the designated area
9. A baseline ECG (6 lead ECG is acceptable) will be obtained if the participant has not had one recently. If the participant has had an ECG in the last 6 months, and they have not started any QT prolonging medications in the interim, the previously obtained ECG can be used.
10. Blinded study drug and 5 oral fluid collection kits are provided to the participant by Study member who reviews dosing of study drug (three pills daily) with the participant.

##### 4.3 Safeguards against coercion and bias during the recruitment and consenting

4.3.1 Sub-study 1: To ensure there is ample time for the participant to consider participating, they will be provided with the informed consent form through DocuSign which includes a detailed summary of the study at the time the study team reaches out to the potential participant to report positive test findings, and gauge general interest in the study. When the participant comes to the EM tent for screening labs, an appropriately delegated and credentialed clinical investigator from the EM Department will review the consent form with the individual and address any questions.

4.3.2 Sub-study 3: Recruitment will occur through a survey, or in person information sessions. This initial introduction to the study will not be made by any employee's direct supervisor or a person in the supervisory hierarchy. This is intended to minimize any feelings of undue influence in recruiting amongst the UPHS healthcare workforce allowing for autonomy in decisions to participate.

*Safeguards against coercion:* UPHS employees who are under the direct supervision of the study PI and/or sub-Investigators are at an increased risk for undue coercion to participate and considered to be a vulnerable population. Assuming that Emergency Medicine and Infectious Disease study team physicians and study team nurses will enroll into Sub-study 3 as participants, a physician investigator from the alternate department will consent the UPHS worker onto this sub-study. For example, if a study physician in EM wants to enroll, the physician investigator obtaining consent would then be from ID. The delegated physician investigator that consents the participant in this sub-study will remind the participant that the decision about research participation will not affect (favorably or unfavorably) performance evaluations, career advancement, or other employment-related decisions made by peers or supervisors. Documentation of these conversations will be recorded as part of the informed consent process and provided in the informed consent document. EM and ID division leadership will be educated for awareness that they may not utilize enrollment in making any decisions about staff performance or advancement. These measures are put into place to minimize any undue coercion and allow for autonomy in decisions to participate. The PI, Primary ID Sub-Investigator Contact and/or Primary EM Sub-Investigator Contact will not enroll to participate personally as participants in the research, this will ensure that there is always at least one non-participating clinical investigator for study operations and ensure the integrity of the data analysis remains intact (unbiased).

*Safeguards against bias for physicians with dual roles:* Any PATCH trial sub-investigator physician (or study team member) who is enrolled as a participant into Sub-study #3 will be removed from certain elements of the study conduct to minimize bias. Such a person with a dual role as both a trial participant and trial study team member:

a. would not be able to perform data entry, review, evaluations associated with their own data; data should be entered, reviewed, and evaluated by a member from the alternate division

b. would not be able to perform data analysis, if the analysis is identifiable; aggregate analysis could be partially allowable in certain circumstances

c. would have some restrictions in performing informed consent for patients enrolling into Sub-study #1 —as above, the physician investigator would not be allowed to mention to the prospective patient being consented that the physician was also participating in the study--- minimizing bias of safety and therapeutic benefit ('if my doctor enrolled, then it's safe and so should I').

##### 4.4 Consenting process

4.4.1 Sub-study 1 and 3: Informed consent will be performed via DocuSign, an electronic mechanism that is both HIPAA and 21 CFR Part 11 compliant. It is important for infection control and research staff safety to use e-consent approaches; use of paper with wet signatures represents an excessive risk of contaminated fomites.
Appropriately delegated and credentialed physician investigators will conduct the consent process and answer any questions from participants via telephone. The e-consent will be obtained via the participant's smartphone/smart device.

4.4.2 Sub-study 3: For this sub-study 3, consent forms will include additional details relevant to healthcare workers as a vulnerable population, ensuring (a) that any medical data collected will not be shared with Penn managers or affect employment, (b) that agreement or refusal to participate will not impact employment or be included in any personnel files, and that (c) physician sub-investigators obtaining consent are not direct supervisors of any participants undergoing enrollment.

##### 4.3 Required Registration information

- Patient's initials (first and last)
- Patient's Hospital ID and/or last 4 digits of Social Security number
- Patient past medical history
- Body mass Index
- Smoking status Current former
- Gender
- Birth date (mm/yyyy)
- Race
- Ethnicity
- Nine-digit ZIP code
- State of residence
- County of residence
- Cell phone number and alternate numbers
- Email address
- Sub-study 1: A description (not containing PHI) for all full time inhabitants at the residence where the participant will stay during the 14-day quarantine.

#### 5.0 RANDOMIZATION

After all applicable screening assessments have been performed, participants who have met all inclusion criteria and none of the exclusion criteria will be centrally, randomly allocated to one of the two groups in each sub-study. Randomization will be done using computer generated randomization numbers. Treatment **should start within 3 working days after randomization.** Once the order for study drug arrives at the pharmacy and the participant is randomized only the pharmacist is aware of the assignment of HCQ versus placebo in the double blind sub-studies (1 and 3).

**Blinding:** Sub-Study 1 and 3 will have a double-blind placebo controlled design.

**Unblinding:** Sub-Study1: If a participant progresses with symptoms of COVID-19 based on the assessment scale outlined below an appropriately delegated and credentialed study team member will review the case with a clinical investigator who will sign off on the need to un-blind the participant. If the participant was on placebo and continues to have symptoms, then he/she will be allowed to crossover to the HCQ dosing. The EM van will deliver the HCQ to the participant. If the patient was on HCQ then HCQ will be stopped and the patient will be asked to seek the advice of their primary care doctor or go to the emergency room if symptoms are becoming more worrisome. Participants who have progressive symptoms and are subsequently hospitalized due to COVID-19 will be unblinded but not be crossed over to receive HCQ at the time of unblinding.

Sub-Study 3: If the participant at any time has worsening symptoms consistent with new onset of COVID-19 disease a COVID-19 PCR test is performed. Antibody tests alone will not be used to make decisions for this study. If the PCR test is positive for SARS-CoV-2 an appropriately delegated and credentialed study team member will contact IDS and the pharmacist will indicate which arm the participant was assigned. If the participant was assigned to placebo and wishes to cross over to HCQ, IDS will provide HCQ at the same dose as the Sub-Study 3 HCQ arm and IDS will ship the HCQ to the participant. If the participant was assigned to HCQ then the participant will be advised to seek the advice of their personal physician or go to the emergency room if symptoms are becoming more worrisome.

#### 6.0 TREATMENT PLAN

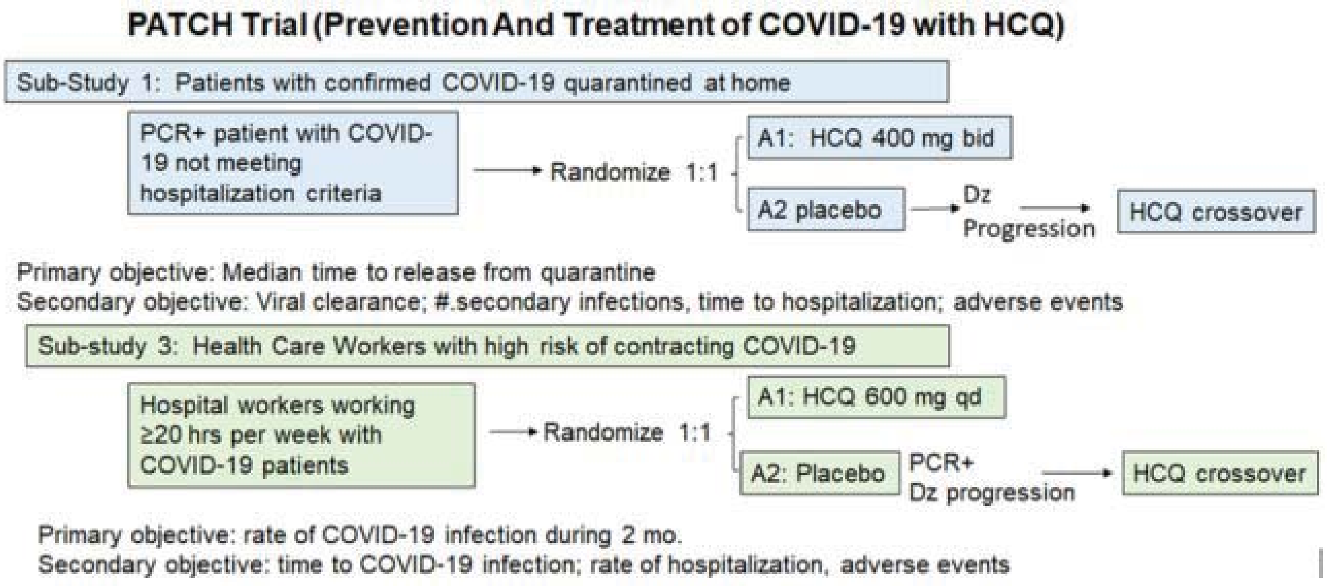

**Fig. 1 PATCH Trial Schema.** Dz: Disease A: Arm

**Sub-Study 1: 100 patients 2 interim analyses**

Arm 1: High dose HCQ: Hydroxychloroquine 400 mg twice a day for up to 14 days
Arm 2: Placebo: 2 pills twice a day for up to 14 days cross over for progressive disease

**Sub-Study 3: 200 participants 2 interim analyses**
Arm 1: High dose HCQ: Hydroxychloroquine 600 mg daily for 2 months
Arm 2: Placebo: 3 pills once a day for 2 months crossover to HCQ 600 mg daily if PCR positive

#### 7.0 DETAILS OF STUDY TREATMENT

##### 7.1 Hydroxychloroquine

Mechanism of Action: The mechanism of action is not fully understood. Previously it was thought that HCQ and other chloroquine derivatives are weak bases that deacidify lysosomes through purely chemical basis. Recently our group has identified the missing molecular target of HCQ as palmitoyl protein thioesterase 1 (PPT1).

Storage and formulation: HCQ tablets are manufactured by Sandoz. Each tablet contains 200 mg hydroxychloroquine sulfate (equivalent to 155 mg base). It is dispensed in a tight, light-resistant container as defined in the USP/NF.HCQ should be stored at room temperature up to 30° C (86° F).

Pharmacokinetics: The PK of HCQ is characterized by a large volume of distribution, binding to red blood cells, and long time to peak concentration and steady state. Population PK studies in cancer patients have demonstrated dose proportional change in exposure.

Administration: Hydroxychloroquine is an oral medication, requiring participants in sub-studies 1 and 3 on study to keep an electronic study diary. Diary must be submitted at 14 days (sub-study 1) and 2 months (sub-study 3). Hydroxychloroquine will be provided by IDS. Tablets of HCQ are available in 200 mg strength. When HCQ is administered in divided doses (every 12 hours) two daily doses of HCQ should be taken 12 hours apart, for example, 9 am and 9 pm, and documented clearly on the patient drug diary. The HCQ schedule may be adjusted if necessary to minimize gastrointestinal side effects.

For complete information please refer to the package inserts at http://dailymed.nlm.nih.gov/dailymed/

**7.2 Placebo:** Excipient only manufactured to match hydroxychloroquine. Manufactured at Temple University.

##### 7.3 Concomitant Medication, Drug-Drug interactions and Procedures

Participants in sub-study 3 must be instructed not to take any medications, except tylenol, including over the counter products such as NSAIDs, without first consulting with the investigator. Participants in sub-study 1 must be instructed not to take any medications, including Tylenol and over the counter products such as NSAIDs, without first consulting with the investigator.

###### Drug Interactions

The following medicines are allowed but should be carefully monitored.
Digoxin: Concomitant HCQ and digoxin therapy may result in increased serum digoxin levels.

Insulin or antidiabetic drugs: As HCQ may mildly enhance the effects of a hypoglycemic treatment, a decrease in doses of insulin or antidiabetic drugs may be required.

Drugs that prolong QT interval and other arrhythmogenic drugs: HCQ can prolong the QT interval and should not be administered with other drugs that have the potential to induce cardiac arrhythmias. Also, there may be an increased risk of inducing ventricular arrhythmias if HCQ is used concomitantly with other arrhythmogenic drugs. A full list of these drugs can be found at https://crediblemed.org

Antiepileptics: The activity of antiepileptic drugs might be impaired if co-administered with HCQ.

Cyclosporin: An increased plasma cyclosporin level was reported when cyclosporin and HCQ were co-administered.

Because HCQ has known effects on P450 enzymes, patients requiring anti-convulsants may be treated with any of the non-enzyme inducing anti-convulsants which include: felbamate, valproic acid, gabapentin, lamotrigine, tiagibine, topiramate, or levetiracetam. Due to the fact that both zonisamide and HCQ accumulate in red blood cells, zonisamide should be avoided if possible. All other concomitant medications are permitted.

The following medications are not allowed during the study. The sponsor must be notified if the subject receives any of these during the study:

1. Sub-study 1: Any investigational or off-label antiviral therapy
2. Immunosuppressive medications, including, corticosteroids at doses exceeding 10mg/day of prednisone or equivalent, methotrexate, azathioprine, and TNF-alpha blockers. Use of immunosuppressive medication for the management of study treatment-related AEs or in subjects with contrast allergies is acceptable. In addition, use of topical, inhaled and intranasal corticosteroids is permitted
3. Live attenuated vaccines during the study
4. Herbal and natural remedies must be avoided

##### 7.4 Duration of Protocol Treatment and Follow-up.

Sub-Study 1 duration of treatment is up to 2 weeks or at the time of release from quarantine. HCQ will be stopped in all patients at the time of release from quarantine
Sub-Study 3 duration of treatment is 2 months with assay to assess SARS CoV2 every 2 weeks.

**7.5 Crossover from Control to HCQ arm**. For Sub-Study 1 all participants are un-blinded upon symptom progression (see response assessment below) after at least 7 days of treatment or upon hospitalization if this occurs prior to 7 days of treatment. If the participant was assigned to placebo, he/she has the option to crossover to receive HCQ 400 mg po bid for up to 2 weeks or until criteria for release from quarantine are met (See 1.0 primary endpoint). During these 2 weeks of HCQ treatment, the patient will continue to complete the daily diary. An appropriately delegated study team member will call the patient daily to assess worsening symptoms and AEs. Patients who are not improving will be sent to the hospital for further care. Participants who have progressive symptoms and are subsequently hospitalized as related to COVID-19 will not be crossed over to receive HCQ at the time of unblinding.

In sub-study 3 participants who have worsening symptoms as defined as fever, significantly worsening cough, shortness of breath or hypoxia but do not require hospitalization and tests positive for SARS-CoV2 virus are allowed to cross over to receive HCQ, or treated with an alternative therapy per physician’s choice. Once the positive test returns, the CRC will email IDS to initiate unblinding IDS will inform the CRC and clinical investigator if the patient was on placebo. The clinical investigator will contact the subject and inform them that if they were on placebo they would have the option of being treated with HCQ 600 mg qd. If the patient agrees, the clinical investigator will place the order in EPIC for HCQ 600 mg qd through IDS. Drug will be delivered to the patient’s home. During these 2 weeks of HCQ 600 mg po qd treatment, clinical investigators will call the patient every 1-3 days depending on the severity of symptoms to assess worsening symptoms and AEs. Patients who are not improving will be sent to the hospital for further care.

#### 8.0 TOXICITY CRITERIA, MONITORING, DOSE DELAYS AND MODIFICATIONS

##### 8.1 Toxicity Criteria

This study will utilize the Common terminology criteria for adverse events version 5.0 (CTCAE v 5.0). https://ctep.cancer.gov/protocolDevelopment/electronic_applications/docs/CTCAE_v5_Quick_Reference_8.5x11.pdf All appropriate treatment areas should have access to a copy of the CTCAE v 5.0 grading tables.

##### 8.2 Dose Delays

Major Events are grade 3 and 4 hematologic and non-hematologic toxicities that are not treatment -related. Treatment should be delayed for major events if HCQ may further complicate the non-treatment related event. If a major event requires a delay of treatment, treatment must be delayed until toxicity is resolved (≤ Grade 2 or ≤ Baseline). For treatment-related toxicities and major events, if toxicity is not resolved in ≤ 14 days, patient will be taken off treatment unless there is an exception granted by the medical monitor.

**8.3 Ocular toxicity.** The only toxicity that requires discontinuation of HCQ is retinopathy. Published literature indicates HCQ retinopathy is idiosyncratic and is uncommon in patients receiving HCQ for less than a few years. Our ongoing study of dabrafenib, trametinib, and HCQ (BAMM; NCTNCT02257424) found no clinically meaningful ocular toxicity in 10 patients studied extensively with serial ocular exams (25). Although HCQ retinotoxicity can occur at shorter than expected intervals with the use of high dose (>5 mg/Kg/day of real body weight) treatment protocols, the cumulative doses required to elicit a retinopathy were only reached after about a year of treatment (26). Thus, we do not anticipate cumulative doses in this trial will reach retinotoxic levels (>200 grams) and have, therefore, not included mandatory ocular exams in this protocol. However, participant will be advised to report with any visual changes at which point a comprehensive ophthalmic exam will take place to decide whether HCQ should be permanently discontinued. As a precaution, patients with renal failure will also be excluded from the trial.

**Laboratory monitoring is not recommended for HCQ use in rheumatoid arthritis.** The American College of Rheumatology guidelines for the treatment of rheumatoid arthritis recommends no laboratory testing for outpatients that are started on hydroxychloroquine (27). HCQ doses for rheumatoid arthritis range from 400mg daily to 400mg twice a day encompassing the dosing paradigms in Sub-study 1 and sub-study 3.

**8.3 Safety of HCQ in pregnancy obviates the need for pregnancy monitoring.** Numerous case series and a recent large scale epidemiological study demonstrate the safety of HCQ in outpatient pregnant women with systemic lupus erythematous (28).Therefore we are not requiring pregnancy test prior to starting HCQ in sub-studies 1 and 3. Potential participants that are known to be pregnant are excluded.

##### 8.4 Participant safety monitoring and data collection

###### 8.4.1 Sub-Study 1:

<u>Clinical symptom monitoring:</u> Participants will be called by an appropriately delegated and credentialed member of the study team () daily at home to monitor clinical symptoms according to the following grading scales:

| Table 1. Example of Daily COVID-19 Symptom tracking score table | | | | |
|---|---|---|---|---|
| Symptom | Mild =1 | Moderate =2 | Severe =3 | Score |
| Abdominal pain | | | | 0 |
| Fatigue | | | | 0 |
| Cough | | 2 | | 2 |
| Shortness of breath | 1 | | | 1 |
| Diarrhea | | | | 0 |
| Myalgias | | 2 | | 2 |
| Headache | | | | 0 |
| Smell disturbance | | | | 0 |
| Other: | | | | 0 |
| Total Score | | | | 5 |

The participant will have a daily diary where this table can be filled in by the participant. The study team member will ask for each symptom and record the answers on an eCRF. At the end of the study the concordance of the participant recorded diary and the assessment made by the study team member will be determined. For discordant cases the study team member recorded AEs will be utilized for data analysis.

AE assessment: HCQ can cause nausea, constipation, diarrhea, rash. Participants will be asked if there have been changes in vision such as distortions of blurred vision. Diarrhea and nausea are known toxicities of HCQ that overlaps with the COVID-19 symptoms. In this case diarrhea or nausea can be considered COVID-19 symptoms by the study team. If an overlapping symptom is worsening while other symptoms are resolving it will be reclassified as a
treatment related AE. Cardiac AEs will be assessed directly by the cardiology Sub-Investigators (see below).

Triage: An appropriately delegated and credentialed member of the study team (as example: research nurse) will document treatment-related AEs, and triage the participant for continued home quarantine versus hospitalization. Follow up with an appropriately delegated and credentialed physician investigator will be undertaken for assessment and grading of events.

Temperature Measurements: Home quarantined participants will be asked to take twice daily temperature and record the measurements on their phone. For those patients that do not have a thermometer, one will be provided by the study. The temperature records in the participant diary will be reviewed for trends within participants and in HCQ v. placebo cohorts.

Mobile Cardiac Telemetry: Participants will be provided with a mobile cardiac telemetry device (ZioAT, iRhythm Technologies). This technology will wirelessly transmit continuous monitoring to Irhythm, Inc. Irhythm will notify the study team for prolonged QTc, or life threatening arrhythmiasin near real time.Dose adjustments will be performed as described below to achieve adjusted QTc <500 ms when necessary. Refusal to wear the mobile cardiac telemetry device will not prevent study participation and/or will not result in removal from the trial. Baseline QTc will be assessed by the cardiology SubI. Subsequent cardiac assessments will only be triggered by Irhythm alerts.

Oral fluid collection: Participants will be asked to collect oral fluid using the RNAPro-Sal collection kit manufactured by Oasis Diagnostics (https://4saliva.com/products/rnapro-sal/). These kits will be provided to the participant with the drug supply. The participant will have clear instructions on how to collect oral fluid on themselves. This process consists of inserting the gauze tip in the mouth for 4 minutes, removing and pushing the syringe to squeeze the fluid into the container. A video will be sent to the participant's email address and texted to them providing visual instructions on use. Participants should produce an early morning saliva sample from the posterior oropharynx (ie, coughed up by clearing the throat) before toothbrushing and breakfast, because nasopharyngeal secretions move posteriorly and bronchopulmonary secretions move by ciliary activity to the posterior oropharyngeal area while the patients are in a supine position during sleep. Participants will be instructed to collect oral fluid at the following time-points in the containers provided (before taking the first dose of study drug, day 3, day 7). The collected oral fluid should be placed in the freezer in a sealed container. This container will be collected after the participant is released from quarantine. The participant will then wipe the container surface with disinfectant and the ED van will come to pick up the sample. The van driver will open a bag and the participant will carefully place the container in the bag, so that the outside of the bag is not contaminated. If the participant becomes hospitalized with COVID-19, a primary contact for the participant will be contacted and if asymptomatic and/or COVID-19 negative the driver will pick up the saliva using the same procedure.

Nares swabs:
*Anterior nares specimen*: Participants will self-collect these samples on day 0 (start date for HCQ) before the HCQ is taken, Day 3, and day 7

*Foam swab provided*: Puritan 6” Sterile Standard Foam Swab w/ Polystyrene Handle (SKU # 25-1506 1PF)

*Collection instructions*: Use a single foam swab for collecting specimens from both nares. Insert foam swab into 1 nostril straight back (not upwards). Once the swab is in place, rotate it in a circular motion 2 times and keep it in place for 15 seconds. Repeat this step for the second nostril using the same swab. Remove foam swab and insert the swab into the supplied tube.

*Storage instructions*: The collected swab should be placed into the bag marked “Nares swabs collected” and the date and time of the swab collection should be filled out Redcap. The “Collected” bag is then placed in the freezer in the same sealed container as the oral fluid.
Specimen stability for all specimen types for all tests is now as follows:
Room temperature: 5 days
Refrigerated (2 °C–8 °C): 5 days
Frozen (-20 °C): 7 days
Frozen (-70 °C): Acceptable

Specimen pickup: At the end of quarantine, or end of study procedures, the nares swabs will be collected. And shipped to Quest Diagnostics to assess viral titers. Cold packs or dry ice should be used during collection and cold packs should be used during shipping to Quest.

Pill diary: When the participant is released from quarantine the electronic pill diary must be submitted electronically.

Monitoring co-inhabitants: Appropriately delegated and credentialed study team members (as example: Clinical Research Coordinator) will ask the participant about the status of co-inhabitants daily to 14 days after the quarantine is completed to determine if the participant’s co-inhabitants did or did not have symptoms or tested positive for COVID-19.

**8.4.2 Sub-Study 3:**

SARS-CoV-2 testing: Hospital workers will be tested for SARS-CoV-2 virus by RT-PCR every month. This will be a nasopharyngeal swab that will be processed by Quest diagnostics performed during enrollment at one month and two months. This screening NP swab will provide results in EPIC for clinical action within 72 hours. Eligibility criteria to start study medication does not require a negative PCR result to return.

Asymptomatic PCR+ at baseline: Given the 20% prevalence of asymptomatic coronavirus infection, we will need to screen 250 subjects to accrue the target enrollment of 200 to address the primary endpoint. if the PCR test returns positive while the health care worker is enrolled on the study, the participant will be notified and offered to be removed from the study or to continue with the study medication. Baseline PCR+ subjects who decide to withdraw from the study will be asked to go to occupational health and seek the care of their primary physician. Their study medication will remain blinded and the subject will be asked not to take any additional study medication. For baseline PCR+ subjects who decide to continue on study, they will be asked to keep the daily pill diary. If these subjects are asymptomatic after one week of study medication, they will be asked to return to undergo an on treatment nasopharyngeal swab. If the subject develops symptoms on study medication after 7 days of treatment, the subject will be unblended. If the subject was assigned to placebo, they will have the option to crossover and get treatment with HCQ 600 mg qd for up to 7 days. Weekly phone calls per the sub-study 3 schedule will continue during this time. AEs will be reported a planned for the study. Baseline PCR+ subjects will be replaced in the prevention study related to the primary objective of sub-study 3 and will not count towards the target accrual or interim analysis. Baseline PCR negative subjects who screen positive at the one month or two-month mark will be considered COVID-19 positive as it relates to the primary endpoint.

Asymptomatic PCR+ at one month: These subjects will count towards the primary endpoint of sub-study 3 as an infection event. Accordingly, they will be unblinded and if on placebo they will be allowed to crossover to HCQ 600 mg qd for 2 weeks. If the subject is assigned to HCQ and turns PCR positive while asymptomatic, the subject will be stopped from study drug and referred to occupational health for standard of care management of health care worker infection. Weekly phone calls will continue. If the subject remains asymptomatic after one month, he/she will return for the one-month swab.

Clinical Symptom monitoring: Appropriately delegated and credentialed study team members (as example: CRC) will screen for symptoms 1X per week and if present will document the likely treatment related AEs. Follow up with an appropriately delegated and credentialed physician investigator will be undertaken for assessment and grading of events. Participants will be asked if there have been changes in vision such as distortions of blurred vision. Patients who develop COVID-19 symptoms will be instructed to go to occupational health for nasopharyngeal swab. The rate of conversion to PCR positive in each arm at the 2-month mark will be used to assess the primary endpoint.

Electrocardiogram Recordings: An electrocardiogram (12 lead or 6 lead) will be recorded at baseline prior to the initiation of the study drug. Those participants with an ECG in the last 6 months and no QTc prolonging medications do not need a repeat baseline ECG. One month after starting the study drug a repeat ECG will be performed to confirm excessive QTc prolongation has not occurred. If the QTc has prolonged to >500ms dose reduction as described below will be attempted to achieve a QTc of <500ms. Due to the long half-life of HCQ, study medication can be started before the baseline ECG is read.

Pill diary: Electronic pill diary will be turned in by the participant at the end of the study.

Oral fluid collection: Participants will be asked to collect oral fluid using the RNAPro-Sal collection kit manufactured by Oasis Diagnostics (https://4saliva.com/products/rnapro-sal/). These kits will be provided to the participant with the drug supply. The participant will have clear instructions on how to collect oral fluid on themselves. This process consists of inserting the gauze tip in the mouth for 4 minutes, removing and pushing the syringe to squeeze the fluid into the container. A video will be sent to the participant's email address and texted to them providing visual instructions on use. Participants should produce an early morning saliva sample from the posterior oropharynx (ie, coughed up by clearing the throat) before tooth brushing and breakfast, because nasopharyngeal secretions move posteriorly and bronchopulmonary secretions move by ciliary activity to the posterior oropharyngeal area while the patients are in a supine position during sleep. Participants will be instructed to collect oral fluid at the following time-points in the containers provided (before taking the first dose of study and every 2 weeks after that). The collected oral fluid should be placed in the freezer in a sealed container. This container will be collected after the participant is at the end of their participation. The participant will then wipe the container surface with disinfectant and return to site. If the participant becomes quarantined or hospitalized with COVID-19, a primary contact for the participant will be contacted and if asymptomatic and/or COVID-19 negative the driver will pick up the saliva using the same procedure.

**8.5 Hydroxychloroquine Dose Reduction.**

In outpatient arms 1 and 3, any AE of ≥ Grade 2 or adjusted QTc prolongation to >500ms and attributed as possibly, probably or definitely related solely to HCQ will result in dose holiday and/or dose reduction of HCQ as described in Table 5. No more than 1 dose reductions are allowed.

**Table 3: Hydroxychloroquine Dose Reduction Schema**

| Sub-Study | Dose mg/day | First dose reduction |
|---|---|---|
| 1 | 400 mg twice daily | 400mg daily |
| 3 | 600 mg daily | 400 mg daily |

- If the second dose reduction is not tolerated, then the patient should discontinue thetreatment.

Toxicities that may be attributable to HCQ include: nausea, anorexia, vomiting, constipation, diarrhea, rash, and visual field deficit. If any of these AEs occur at grade < 2, HCQ may be continued and the AE managed with supportive care. For any AE with a grade ≥ 3, HCQ dose will be held until the toxicity resolves to ≤ grade 2, after which HCQ may be restarted at a reduced dose as described in table 5. With particular regard to visual field deficits participants should be cautioned to report any visual symptoms, particularly difficulty seeing entire words or faces, image distortions, intolerance to glare, decreased night vision, or loss of peripheral vision. **These symptoms of retinal toxicity or subclinical evidence of retinal toxicity on eye exam should prompt drug discontinuation and ophthalmologic evaluation at Scheie Eye Institute Urgent care.** The natural course of COVID_19 is not fully understood and additional toxicities attributable to HCQ may emerge when used to treated COVID-19.

**8.6** Procedure for handling of potentially contaminated material: For sub-study 1 institutional and CDC guidelines will be followed. Patients and subjects on sub-studies 1 and 3 will be instructed on safe practices for storage of oral fluid and disinfecation of outer layers of bags and receptacles. Proper hand washing techniques will be reviewed with subjects. Changes in recommendations from the CDC or the University of Pennsylvania will be immediately implemented.

#### 9.0 SCHEDULE OF EVENTS

| **Table 4. Schedule of events for sub-study 1** | | | |
|---|---|---|---|
| | **Before treatment** | **During treatment** | **Day of release from quarantine** |
| Eligibility checklist | X | | |
| Temperature (patient obtained) | | Twice daily | X |
| COVID-19 symptom score | | Daily | |
| Mobile Cardiac Telemetry | X | Daily | X |
| AE assessment, triage | | Daily as needed | X |
| SARS-COV-2 PCR nasophranyngeal swab | X | | |
| SARS-COV-2 PCR nares swab[1] | X | D3, D7 | X (if ≤7 or 10 days, per CDC guidelines) |
| Oral fluid collection[2] | X | D3, 7 | X |
| **TREATMENT** | | | |
| Hydroxychloroquine or placebo | | Twice ev ery day | Return electronic diary |

1- The nares swab before treatment sample can be performed at any time of day and should be collected before any study medication has been taken.

2-For oral fluid collections, these should be performed in the morning to maintain consistency.
For the before treatment collection, if HCQ is received during the day, there should not be a delay
In starting HCQ in order to collect morning oral fluid. Instead study medication should be started and
The next morning the first sample can be collected and considered the baseline sample. The timing of collection should be recorded in the daily diary.

| **Table 6. Schedule of events for Sub-study 3** | | | |
|---|---|---|---|
| | **Baseline / Pre-RX** | **During 2 months** | **End of 2 months** |
| Eligibility checklist | X | | |
| COVID-19 symptom score | X | Weekly | |
| AE check by telephone | | 1X/week | X |
| PCR SARS-COV-2[1] | X | One month | X[2] |
| Oral fluid collection | X | Every 2 weeks | X |
| Research Blood [2] | X | One month | X |
| 6 or 12 Lead Electrocardiogram | X | One month | |
| **TREATMENT** | | | |
| Hydroxychloroquine or placebo | | 3 pills daily | |

1- this testing is dependent on availability of the test kits and can be deferred if test kits or testing capacity are not available
2- End of 2 months or at the time of symptom progression 3- one 10-mL tiger top serum separator tubes (SST). This collection can be deferred to accommodate the conditions of the COVID-19 crisis situation and how it affects resources and health care worker schedules.
3- If subject has had an EKG in the last 6 months the baseline EKG can be deferred. Since this EKG is for research purposes only, the serial EKGs can be deferred to accommodate the conditions of the COVID-19 crisis and how it affects resources and health care worker schedules.

#### 10.0 MEASUREMENT OF EFFECT

##### 10.1 Definitions

<u>Evaluable for toxicity</u>. All participants will be evaluable for toxicity from the time of their first treatment with HCQ or placebo.

<u>Evaluable for primary outcome: Participants</u> who received at least one dose of study drug will be evaluable for the primary outcome as long as they meet the following status for SARS-COV2 PCR: sub-study 1: PCR+ Sub-study 3: PCR negative.

##### 10.2 Response Criteria

<u>Sub-Study 1:</u> Time to release from quarantine. The criterion is afebrile for 72 hours AND improvement of symptoms AND at least 7 or 10 days have passed since symptoms started (according to contemporary CDC guidelines). To quantitatively assess improvement of symptoms the grading system in Section 8.5 will be used.

May 2020. There was a change in CDC guidelines on quarantine time following onset of symptoms, changing from 7 to 10 days. We have adopted the new guidelines, but some of the earlier patients enrolled in the study were released in a shorter time than they would have been under the new guidelines. Because the response is time to release, we will record both release dates for each patient. While the release based on 10 days will be the primary outcome, we will run a parallel analysis of the 7-day measure as part of a sensitivity analysis.

Sub-Study 3: Measurement of SARS-CoV-2 positivity defined by PCR. Time to infection will be defined as the time in days from enrollment on the study to time to COVID-19 symptoms.

##### 10.3 Off treatment/Off Study

Each participant has the right to withdraw from the study at any time without prejudice. The investigator may discontinue any person's participation for any reason, including adverse event or failure to comply with the protocol. Should a participant withdraw from the study, the reason(s) must be stated on the case report form, and a final evaluation of the participant should be performed. Reasons for withdrawal include the following:

Progression of Disease: Remove participant from protocol therapy at the time progressive disease is documented. Progression of disease is left to the discretion of the treating physician/ sub-Investigator; however, the following guidelines are provided to help decision making for discontinuing study drug:

Sub-study 1: Progression of disease can be confirmed if there is increase in COVID-19 symptom score for 2 days in a row and is three points higher than baseline. Once criteria for release from quarantine is met in SS1 the patient is off study. Once the person is hospitalized for COVID-19 in sub-study 1 the subject is off study. Participants who have progressive symptoms and are subsequently hospitalized as related to COVID-19 will not be crossed over to receive HCQ at the time of unblinding.

Sub-study 3: Development of new symptomology consistent with COVID-19: e.g. fever, cough, shortness of breath and SARS-COV2 test positivity would lead to unblinding. If on placebo the subject can crossover to HCQ for treatment and stay on study for 14 days or release from quarantine. If the subject was on HCQ the subject is off study and will be instructed to seek medical attention through their primary care physician

Extraordinary Medical Circumstance: If at any time the treating physician feels constraints of this protocol are detrimental to the participant's health remove the participant from protocol therapy.

Participant's refusal to continue treatment: In this event, document the reason(s) for withdrawal.

Failure to comply with protocol (as judged by the investigator such as compliance below 80%, failure to maintain appointments, etc.).

Withdrawal from treatment per patient choice: Patients may withdraw from study treatment at any point in the study for any reason. These subjects are allowed to withdraw from treatment but continue to participate with any of the other study procedures for the intended 2 month period.

Delay in treatment > 7 days due to toxicity

Once the subject has completed 2 months the person is off study.

#### 11.0 ADVERSE EVENTS AND REPORTING

The timely reporting of adverse events (including toxic deaths) is required by the Food and Drug Administration. The reporting of toxicities is part of the data reporting for this study. The sponsor-investigator is responsible for ensuring that all adverse events (AEs) and serious adverse events (SAEs) that are observed or reported during the study are collected. Misuse and abuse of the study medication, medication errors and uses outside of what is foreseen in the protocol (irrespective if a clinical event has occurred) must be reported within 24 hours to the Study Sponsor/ Principal Investigator.

Adverse Events

An ***adverse event*** (AE) is any symptom, sign, illness or experience that develops or worsens in severity during the course of the study that does not necessarily have a causal relationship with this treatment. Intercurrent illnesses or injuries should be regarded as adverse events. Abnormal results of diagnostic procedures are considered to be adverse events only if the abnormality:

- results in study withdrawal
- is associated with a serious adverse event
- is associated with clinical signs or symptoms
- leads to additional treatment or to further diagnostic tests is considered by the investigator to be of clinical significance

**Adverse Event Reporting Period**

The study period during which adverse events must be reported is defined as the period from the initiation of the first study treatment to the last administration of study treatment.

**Post-study Adverse Event.**

All unresolved adverse events should be followed by the clinical investigator until the events are resolved, the participant is lost to follow-up, or the adverse event is otherwise explained. At the last scheduled visit, the investigator should instruct each participant to report any subsequent event(s) that the participant, or the participant's personal physician, believes might reasonably be related to participation in this study.

**Abnormal Laboratory Values.**

A clinical laboratory abnormality should be documented as an adverse event if the abnormality is grade 1 or more and a change from baseline.

##### 11.1 Recording of Adverse Events.

At each contact with the participant, the appropriately delegated and credentialed study team member must seek information on adverse events by specific questioning and, as appropriate, by examination. Information on all adverse events should be recorded immediately in the source document, and also in the appropriate adverse event module of the case report form (CRF). All clearly related signs, symptoms, and abnormal diagnostic procedures results should be recorded in the source document, though should be grouped under one diagnosis.

All adverse events occurring during the study period must be recorded. Adverse events will be measured and graded in accordance with the CTCAE. The clinical course of each event should be followed until resolution, stabilization, or until it has been determined that the study treatment or participation is not the cause. Serious adverse events that are still ongoing at the end of the study period must be followed up to determine the final outcome. Any serious adverse event that occurs after the study period and is considered to be at least possibly related to the study treatment or study participation should be recorded and reported immediately.

###### 11.1.1 Serious Adverse Events

Adverse events are classified as serious or non-serious.

A serious adverse event is any AE that is:

- fatal
- life-threatening
- requires or prolongs hospital stay
- results in persistent or significant disability or incapacity
- a congenital anomaly or birth defect
- Suspected transmission of an infectious agent (eg, pathogenic or nonpathogenic) via the study drug is an

SAE.

- an important medical event

Important medical events are those that may not be immediately life threatening, but are clearly of major clinical significance. They may jeopardize the participant, and may require intervention to prevent one of the other serious outcomes noted above. For example, drug overdose or abuse, a seizure that did not result in in-patient hospitalization, or intensive treatment of bronchospasm in an emergency department would typically be considered serious. Theft, sale, or use of the study product by any person other than the participant will be reported as a medically important event.

**ADVERSE EVENT COLLECTION AND REPORTING INFORMATION:**

- All Serious Adverse Events (SAEs) that occur following the participant's written consent to participate in the study must be reported, whether related or not related to study drug. If applicable, SAEs must be collected that relate to any later protocol-specified procedure.
- Following the participant's written consent to participate in the study, all SAEs, whether related or not related to study drug, are collected, including those thought to be associated with protocol-specified procedures. The investigator-sponsor should report any SAE occurring after these aforementioned time periods, which is believed to be related to study drug or protocol-specified procedure.
- An SAE report should be completed for any event where doubt exists regarding its seriousness;
- If the investigator believes that an SAE is not related to study drug, but is potentially related to the conditions of the study (such as withdrawal of previous therapy or a complication of a study procedure), the relationship should be specified in the narrative section of the SAE Report Form.

All SAEs should be followed to resolution or stabilization.

Adverse events can be spontaneously reported or elicited during open-ended questioning, examination, or evaluation of a participant. (In order to prevent reporting bias, participants should not be questioned regarding the specific occurrence of one or more AEs.)

**All adverse events that do not meet any of the criteria for serious should be regarded as non-serious adverse events. Non-serious adverse events are documented and assessed as adverse events noted above.**

**Hospitalization, Prolonged Hospitalization or Surgery.**

Any adverse event that results in hospitalization or prolonged hospitalization should be documented and reported as a serious adverse event unless specifically instructed otherwise in this protocol. Any condition responsible for surgery should be documented as an adverse event if the condition meets the criteria for an adverse event.

Neither the condition, hospitalization, prolonged hospitalization, nor surgery are reported as an adverse event in the following circumstances:

- Hospitalization or prolonged hospitalization for diagnostic or elective surgical procedures for a preexisting condition. Surgery should not be reported as an outcome of an adverse event if the purpose of the surgery was elective or diagnostic and the outcome was uneventful.
- Hospitalization or prolonged hospitalization required to allow efficacy measurement for the study.
- Hospitalization or prolonged hospitalization for therapy of the target disease of the study, unless it is a worsening or increase in frequency of hospital admissions as judged by the clinical investigator.

##### 11.2 Assessment of Adverse Events

All AEs and SAEs whether volunteered by the participant, discovered by study personnel during questioning, or detected through physical examination, laboratory test, or other means will be reported appropriately. Each reported AE or SAE will be described by its duration (i.e., start and end dates), regulatory seriousness criteria if applicable, suspected relationship to the study drug (see following guidance), and actions taken. To ensure consistency of AE and SAE causality assessments, only the PI or physician sub-investigators may grade AEs, and investigators should apply the following general guideline:

###### 11.2.1 Relationship to study drug: Yes

There is a plausible temporal relationship between the onset of the AE and administration of the study drug, and the AE cannot be readily explained by the participant's clinical state, intercurrent illness, or concomitant therapies; and/or the AE follows a known pattern of response to the study drug; and/or the AE abates or resolves upon discontinuation of the study drug or dose reduction and, if applicable, reappears upon re-challenge.

###### 11.2.2 Relationship to study drug: No

Evidence exists that the AE has an etiology other than the study drug (e.g., preexisting medical condition, underlying disease, intercurrent illness, or concomitant medication); and/or the AE has no plausible temporal relationship to study drug administration (e.g., cancer diagnosed 2 days after first dose of study drug).

Expected adverse events are those adverse events that are listed or characterized in the Package Insert.

Unexpected adverse events are those not listed in the Package Insert. This includes adverse events for which the specificity or severity is not consistent with the description in the Package Insert. For example, under this definition, hepatic necrosis would be unexpected if the Package Insert only referred to elevated hepatic enzymes or hepatitis.

###### 11.2.3 Diagnosis vs. Signs and Symptoms

If known at the time of reporting, a diagnosis should be reported rather than individual signs and symptoms (e.g., record only liver failure or hepatitis rather than jaundice, asterixis, and elevated transaminases). However, if a constellation of signs and/or symptoms cannot be medically characterized as a single diagnosis or syndrome at the time of reporting, it is ok to report the information that is currently available. If a diagnosis is subsequently established, it should be reported as follow-up information.

###### 11.2.4 Deaths

All deaths that occur during the protocol-specified AE reporting period (see Section 12.1), regardless of attribution, will be reported to the appropriate parties. When recording a death, the event or condition that caused or contributed to the fatal outcome should be reported as the single medical concept. If the cause of death is unknown and cannot be ascertained at the time of reporting, report "Unexplained Death".

###### 11.2.5 Preexisting Medical Conditions

A preexisting medical condition is one that is present at the start of the study. Such conditions should be reported as medical and surgical history. A preexisting medical condition should be re-assessed throughout the trial and reported as an AE or SAE only if the frequency, severity, or character of the condition worsens during the study. When reporting such events, it is important to convey the concept that the preexisting condition has changed by including applicable descriptors (e.g., "more frequent headaches").

###### 11.2.6 Pregnancy

Female Participants

*The PATCH Trial*
Participants should not become pregnant while on this study and for 90 days after last dose of study drug. In addition, participants should not breastfeed while on this study as these drugs may also affect a breast-feeding child. Pregnant women and women who are breast-feeding are not allowed to participate in this study. Participants must agree to use two medically accepted forms of birth control including condoms, diaphragms, cervical cap, an intra-uterine device (IUD), surgical sterility (tubal ligation or a partner that has undergone a vasectomy), or oral contraceptives, OR must agree to completely abstain from intercourse for at least two weeks before receiving first dose of study drug, during participation in this study and for 90 days after last dose of study drug. Abstinence at certain times of the cycle only, such as during the days of ovulation, after ovulation and withdrawal are not acceptable methods of birth control. Even when an approved contraceptive method is used, there is always a small risk that the participant could still become pregnant. Subjects must agree to self-report pregnancy to the study team during the 90 days after the last dose of study drug.

Male Participants
Participants should not father a child while on this study. If the participants spouse or partner has the potential to become pregnant, the participant and partner must use two medically accepted forms of birth control including condoms, diaphragms, cervical cap, an intra-uterine device (IUD), surgical sterility (vasectomy or a partner that has undergone a tubal ligation), or oral contraceptives, OR must agree to completely abstain from intercourse during participation in this study and for 90 days after last dose of study drug. If the partner is taking oral contraceptives, she must begin taking them at least two weeks before the participants first dose of study drug. Abstinence at certain times of the cycle only, such as during the days of ovulation, after ovulation and withdrawal are not acceptable methods of birth control.

If a female participant becomes pregnant while receiving investigational therapy or within 90 days after the last dose of study drug, she must agree to self-report the pregnancy to the clinical investigator. The clinical investigator must immediately notify sponsor-investigator (who will then notify Novartis/Sandoz) in accordance with SAE reporting guidelines. Follow-up to obtain the outcome of the pregnancy should also occur. Abortion, whether accidental, therapeutic, or spontaneous, should always be classified as serious, and expeditiously reported as an SAE. Similarly, details of the birth, and the presence or absence of any congenital anomaly/birth defect or maternal and/or newborn complications in a child born to a female participant exposed to the study drug should be reported as an SAE. Follow-up information regarding the course of the pregnancy, including perinatal and neonatal outcome and, where applicable, offspring information must be reported to the sponsor-investigator (and Novartis/ Sandoz). In order for sponsor-investigator or designee to collect any pregnancy surveillance information from the female participant a signed informed consent addendum for disclosure of this information must be obtained (and will be supplied to the IRB for review at the time the incidental pregnancy is encountered, not anticipated and therefore not included with the original protocol documents). Any pregnancy that occurs in a female partner of a male study participant should be reported. Information on this pregnancy will be collected on the Pregnancy Surveillance Form (supplied for IRB review in the event pregnancy in a female partner of a male participant is encountered). In order for sponsor-investigator or designee to collect any pregnancy surveillance information from the female partner, the female partner must sign an informed consent form for disclosure of this information (form to be supplied in the event such a pregnancy was to be encountered).

##### 11.3 UPENN IRB Notification by Investigator-Sponsor

The University of Pennsylvania IRB (Penn IRB) requires expedited reporting of those events related to study participation that are unforeseen and indicate that participants or others are at increased risk of harm. The Penn IRB will not acknowledge safety reports or bulk adverse event submissions that do not meet the criteria outlined below. The Penn IRB requires researchers to submit reports of the following problems within 10 working days from the time the investigator becomes aware of the event:

- Any adverse event (regardless of whether the event is serious or non-serious, on-site or off-site) that occurs any time during or after the research study, which in the opinion of the principal investigator is:

**Unexpected** (An event is “unexpected” when its specificity and severity are not accurately reflected in the protocol-related documents, such as the IRB-approved research protocol, any applicable investigator brochure, and the current IRB-approved informed consent document and other relevant sources of information, such as product labeling and package inserts.)

**AND**

**Related to the research procedures** (An event is “related to the research procedures” if in the opinion of the principal investigator or sponsor, the event was more likely than not to be caused by the research procedures.)

Deaths occurring for patients on-study and within 30 days of study drug administration that are considered unforeseen and indicates participants or others are at increased risk of harm (i.e. unexpected and probably/definitely related), must be reported to the IRB within 24 hours of notification.

See: https://irb.upenn.edu/ for additional reporting details.

##### 11.3.1 Reporting Process to IRB at Penn.

Principal Investigators are encouraged to submit reports of unanticipated problems posing risks to participants or others using the form: “**Unanticipated Problems Posing Risks to Participants or Others Including Reportable Adverse Events**” via HS-ERA or a written report of the event within 7 working days. FDA Notification by Investigator-Sponsor.

This study is IND exempt and reporting to the FDA is voluntary using a MedWatch 3500 or via the FDA’s website for voluntary reporting.

##### 11.4 Medical Monitoring

It is the responsibility of the Sponsor-Investigator to oversee the safety of the study at this site. This safety monitoring will include careful assessment and appropriate reporting of adverse events, as noted above. Medical monitoring by an independent clinician, Dr. Sunita Nasta, Department of Medicine, will include a regular assessment of the number and type of serious adverse events on a periodic basis.

##### **11.5** Data Safety and Monitoring Board (DSMB)

#### DSMB Description

The Data and Safety Monitoring Board (DSMB) for the Prevention And Treatment of COVID-19 with Hydroxychloroquine (PATCH) trial has been selected by the Sponsor-Principal Investigator Dr. Ravi Amaravadi and acts in an advisory capacity to Dr. Amaravadi for the PATCH trial to monitor participant safety, data quality and evaluate the progress of the study. Operational details of the DSMB are provided in the PATCH trial DSMB Charter.

- This DSMB will consist of 5 members with expertise in statistics, clinical trials, infectious disease, cardiology. The DSMB chair may change the membership of the DSMB in consultation with the PI.
- Observers from other research committees at PENN can be invited to join the open session of the DSMB deliberation.
- This DSMB will be independent of the Sponsor, regulatory agencies, IRBs/ECs, and investigators.

#### Roles and Responsibilities

The primary charge of the DSMB is to monitor the study for subject safety. Formal DSMB safety reviews occur as outlined in the charter. Additional review may be required. The DSMB may monitor effectiveness outcomes to determine relative risk/benefit, futility, or for early termination due to overwhelming effectiveness.

The primary safety endpoint as well as guidance for the conduct of analyses and guidelines/stopping rules are established in the protocol and are reviewed by the DSMB.

The DSMB may also review data related to study conduct. Data to be reviewed and listed in the DSMB reports may include: enrollment rates over time, time from last subject enrolled to date of report (indication of delay between treatment or follow-up and reporting), summary of protocol violations as specified in monitoring reports generated by the Office of Clinical Research (OCR), completeness of treatment and follow-up visit data, and follow-up duration for the population included in the report. Protocol deviations, SAE reports, and assistance in day to day medical decision making on study participants will be handled by the Medical Monitor Dr. Sunita Nasta who is also a member of the DSMB. The DSM will in turn be able to review formal reports generated by the OCR monitor and the interim analysis reports generated by the Sponsor-Investigator and his team.

The DSMB:

- Meets as outlined in this charter
- Provides recommendations about continuing, modifying, and stopping the study
- Operates according to the procedures described in this charter

##### 11.7 Study Monitoring Plan

This study will be monitored by the Principal Investigator and sub-investigators, as appropriate. Such monitoring will include at least weekly meetings of the study team to review accrual, toxicity, SAEs. Dose escalations and study finding. In addition, the PI will ensure that data are completed in a timely manner and he or his designee will review the data for accuracy, completeness and integrity. Further, the PI will view real-time toxicity and all laboratory results on an ongoing basis by accessing the Monitoring Report database and the Protocol Labs database.

##### 11.8 Auditing and Inspecting

The Investigator is required to permit direct access to the facilities where the study takes place, source documents, CRFs and applicable supporting records of study subject participation for audits and inspections by IRB, regulatory authorities (FDA) and academic authorized representatives (OCR). The Investigator will make every effort to be available for the audits and/or inspections.

#### 12.0 Sample collection and processing for Correlative Science

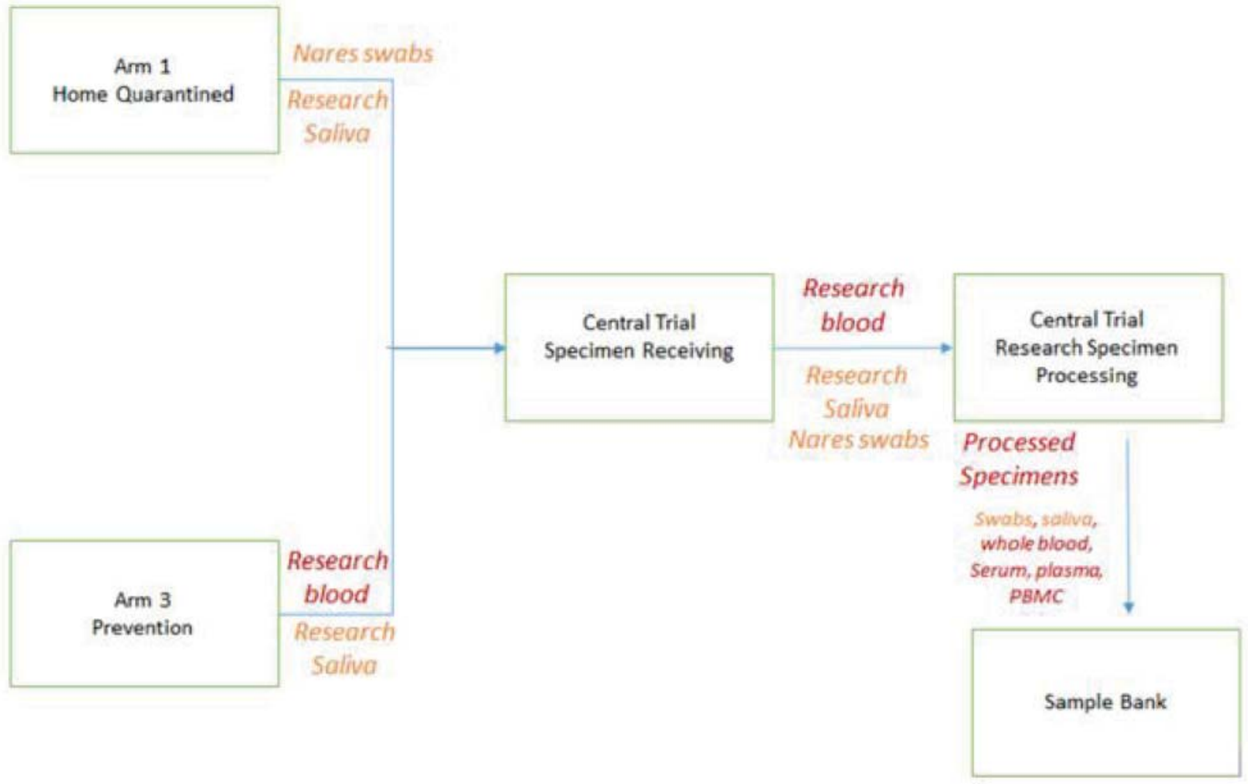

Fig. 2. PATCH Sample Flow

##### 12.1 Research Specimen Process, Banking and Analysis

Blood and oral fluid specimens collected as delineated in 9.0 Schedule of Events. Samples will be assigned a unique identified and then processed and frozen as per SOP within a BSL2+ space that is approved by the University of Pennsylvania's Environmental Health and Radiation Safety (EHRS) program. The processed samples will be stored in the University of Pennsylvania within locked and temperature monitored freezers, and sample tracking. Documentation for sample receipt and processing will be collected and stored in either locked cabinet for paper records or within an encrypted and password protected database. The planned research analyses outlined in the exploratory objectives will be performed using qualified and if possible validated methods; however, due to nature of these research procedures, they may not conform to principles of good clinical laboratory practice.

#### 13.0 STATISTICAL CONSIDERATIONS

##### 13.1 Sample size calculation

Because we are uncertain about the outcome values for both control and treatment arms of the study, we will use group-sequential methods, initially proposing a real difference that is small, and allowing us to stop early for both efficacy and futility. We conduct two interim analyses and a final analysis, after approximately 1/3, 2/3, and 100% of subjects have completed. We will use the Hwang-Shih-DeCani alpha spending rules. Boundaries for efficacy and futility decisions are show below. Futility boundaries arenon-binding.

| z-value | —— Boundaries —— | | Information |
|---|---|---|---|
| **Stage** | **Efficacy** | **Futility** | **Proportion** |
| 1 | 2.7819 | -0.8235 | 0.3400 |

| | | | |
|---|---|---|---|
| 2 | 2.2653 | 0.4262 | 0.6800 |
| 3 | 1.6813 | 1.6813 | 1.0000 |

| p-value Stage | Boundaries Efficacy | Futility | Information Proportion |
|---|---|---|---|
| 1 | 0.00270 | 0.79490 | 0.3400 |
| 2 | 0.01175 | 0.33497 | 0.6800 |
| 3 | 0.04636 | 0.04636 | 1.0000 |

Because we are uncertain about the outcome values for both control and treatment arms of the study, we will use group-sequential methods, initially proposing a real difference that is small, and allowing us to stop early for both efficacy and futility. We conduct two interim analyses and a final analysis, after approximately 1/3, 2/3, and 100% of subjects have completed. We will use the Hwang-Shih-DeCani alpha spending rules. Boundaries for efficacy and futility decisions are show below. Futility boundaries arenon-binding.

**Sub-Study 1, Home quarantined patients with COVID-19:** The first date of quarantine will be considered the date of SARS-CoV-2 nasopharyngeal swab test that confirmed positivity.

The CDC guidelines for release from quarantine change during the course of this study. The original criteria were: 1) 72 hours without a fever 2) improvement in symptoms 3) 7 days since the beginning of symptoms have elapsed. The new criteria are: 1) 72 hours without a fever 2) improvement in symptoms 3) 10 days since the beginning of symptoms have elapsed. Our original sample size calculation was as follows:

Given the median duration of viremia is 14-20 days in mild cases, we will assume the null hypothesis that in the placebo cohort the median time to release from quarantine is 10 days from receiving a positive test. In order for HCQ to be considered more effective in this treatment setting we aim to see a median time to release from quarantine of 5 days. These values correspond to daily hazard rates of 0.069 and 0.139 respectively. We will randomize 1:1 HCQ to control and with 100 patients with 50 in the placebo arm and 50 in the HCQ arm. The one-sided z- test (alpha=.05) will have an overall 95% power to detect a significant difference between the two groups with medians of 10 and 5 days.

Based on the new CDC guidelines for quarantine release we will modify the analysis as follows:
Given the median duration of viremia is 14-28 days in mild cases, we will assume the null hypothesis that in the placebo cohort the median time to release from quarantine is 14 days from receiving a positive test. In order for HCQ to be considered more effective in this treatment setting we aim to see a median time to release from quarantine of 7 days. These values correspond to daily hazard rates of 0.048 and 0.094 respectively. We will randomize 1:1 HCQ to control and with 100 patients with 50 in the placebo arm and 50 in the HCQ arm. The one-sided z- test (alpha=.05) will have an overall 93% power to detect a significant difference between the two groups with medians of 14 and 7 days.

The *hypothesis* will be tested using a one-sided test, with a z-score corresponds to the log of the hazard-ratio for recovery between the two groups. We will obtain estimates of the hazard ratio and group median survival, and 95% CI, from Cox regression. As a secondary analysis, we will stratify the analysis, and obtain separate estimates of the hazard-ratio by group. However, we are not powered to test the interaction for significance, and we do not expect significance within group.

Given that 20 subjects were recruited and treated under the old CDC guidance, we will conduct a primary analysis based on the updated 10-days from onset criterion. For the subjects who were released under the old criterion, this number will be constructed by assessing when a patient would have been released under the new rules. We will follow that with additional secondary analyses to assess the sensitivity of the results to the change in recommendations. These secondary analyses will include a) an analysis using mixed patients as released by the recommendation that was in place at the time, b) a stratified analysis, and c) an analysis of constructed release time based on the original 7-days from onset recommendation. We anticipate that these will yield similar results and that the conclusions will be insensitive to the changes made by the CDC.

Interim analysis: We will perform two interim analyses targeting 34% and 68% completion of participant enrollment, testing for early efficacy or futility, using z-score boundaries that follow Hwang-Shih-DeCani alpha spending rules. The determination of information for survival analysis is somewhat imprecise during the planning stages; for that reason, we will conduct analyses when 34% and 68% of the final sample have had 10 days of follow-up, and determine boundaries based on actual information available at analysis time. Following those rules, we have 93% power using 100% of the sample, if our under our 7 days versus 14 days scenario is true. Under those circumstances, we expect to have a 30% chance of stopping for early efficacy at the first interim analysis. We also have a 40% chance of stopping early for efficacy at the first interim analysis if the placebo arm really had 16 days median time instead of 14 days. Under normal circumstances (14 days v 7 days), we have a 48% chance of stopping for early efficacy at the second interim analysis. We also have a 63% chance of stopping early for efficacy at the second interim analysis if the placebo arm really has 16 days median time instead of 14.

**Z-Value Boundaries**

Maximum Information: 2000.1128
Alternative Hypothesis: h1 - h2 < 0 (one-sided)
Futility Boundaries: Non-Binding

| | Boundaries | | Time | | Information |
|---|---|---|---|---|---|
| **Stage** | **Efficacy** | **Futility** | **Proportion** | **Time** | **Proportion** |
| 1 | -2.7370 | 0.7159 | 0.4000 | 24.00 | 0.3660 |
| 2 | -2.2814 | -0.3990 | 0.6300 | 37.80 | 0.6740 |
| 3 | -1.6808 | -1.6808 | 1.0000 | 60.00 | 1.0000 |

**P-Value Boundaries**

Maximum Information: 2000.1128
Alternative Hypothesis: h1 - h2 < 0 (one-sided)
Futility Boundaries: Non-Binding
P-value boundaries are one-sided values.

| | Boundaries | | Time | | Information |
|---|---|---|---|---|---|
| **Stage** | **Efficacy** | **Futility** | **Proportion** | **Time** | **Proportion** |
| 1 | 0.00310 | 0.76297 | 0.4000 | 24.00 | 0.3660 |
| 2 | 0.01126 | 0.34494 | 0.6300 | 37.80 | 0.6740 |
| 3 | 0.04640 | 0.04640 | 1.0000 | 60.00 | 1.0000 |

**Sub-Study 3, prophylaxis in hospital workers:** The transmission of SARS-COV2 from patient to hospital worker depends on many factors including specifics of standard care to prevent transmission, but especially on the number of patients seen at a given hospital or outpatient practice. Across China the reported hospital worker infection rate is 3.8%, but in Wuhan it is reported as 58% at the height if the epidemic. We will use a

10% transmission rate as the null hypothesis (low dose group). In order for HCQ to be considered effective our alternative hypothesis will be a 1% transmission rate. With a 1:1 randomization for the HCQ to control arms we would require a total of 200 hospital worker subjects total for Arm 4. With the placebo group of 100 subjects and the h HCQ arm of 100 subjects, a one-sided z-test (alpha=0.05) comparing the rates in the two groups would have an 80% power to detect a significant difference when the difference in the population rates is at least 9%.

The *hypothesis* will be tested using a one-sided test, with a z-score corresponds to the log of the odds-ratio for recovery between the two groups. We will obtain the estimate of the odds ratio, and 95% CI, from logistic regression.

Interim analysis: We will perform two interim analyses at 25% and 50% completion of participant enrollment, testing for early efficacy or futility, using z-score boundaries that follow Hwang-Shih-DeCani alpha spending rules. Following those rules, we have 84% power using 100% of the sample, if our under our 10% versus 1% scenario is true. Under those circumstances, we have a 6% chance of stopping for early efficacy at the first interim analysis. We also have an 81% chance of stopping early for efficacy at the first interim analysis if the low dose arm really has 45% infection instead of 10%. Under normal circumstances (10% v 1%), we have a 27% chance of stopping for early efficacy at the second interim analysis. We also have an 80% chance of stopping early for efficacy at the second interim analysis if the low dose arm really has 23% infection instead of 10%.

In addition, at our second interim analysis for SS3, we will evaluate the conditional power based on our original assumptions. The actual effect size for this study is completely unknown. If the effect size is smaller than we anticipated but still promising, and our conditional power falls between 0.5 and 0.8, we will consider formally adjusting the sample size in SS3 to achieve a conditional power of 0.8 based on an updated understanding of the effect size (29).

**Z-Value Boundaries**

| | Boundaries | | Information |
|---|---|---|---|
| **Stage** | **Efficacy** | **Futility** | **Proportion** |
| 1 | 2.9473 | -1.2318 | 0.2500 |
| 2 | 2.5825 | -0.2676 | 0.5000 |
| 3 | 1.6664 | 1.6664 | 1.0000 |

**P-Value Boundaries**

| | Boundaries | | Information |
|---|---|---|---|
| **Stage** | **Efficacy** | **Futility** | **Proportion** |
| 1 | 0.00160 | 0.89099 | 0.2500 |
| 2 | 0.00491 | 0.60550 | 0.5000 |
| 3 | 0.04782 | 0.04782 | 1.0000 |

#### 12.2 Analysis of Secondary Endpoints.

Secondary outcomes will at least be analyzed as summary statistics, with group means, odds-ratios, and 95% CIs. We may conduct exploratory analyses using regression methods appropriate for each type of measure.

**Sub-Study 1:** Rate of secondary infection of housemates is a binomial count by household. We will summarize the proportion or rate by group, with 95% CI. With sufficient data, we will explore subgroups, patient, and secondary patient characteristics using logistic regression. Rate of hospitalization is a binary measure. We will summarize rates by group, and estimate the odds-ratio for treatment. Adverse event rates will be summarized as a count by subject, and mean count by group, and we will estimate a rate-ratio for treatment.

**Sub-Study 3:** For hospital employees, Adverse event rates will be summarized as a count by subject, and mean count by group, and we will estimate a rate-ratio for treatment.

### Data and Safety Monitoring Board Charter

**Trial Title(s) per IND/IDE: The PATCH Trial Prevention and Treatment of COVID-19 with Hydroxychloroquine**

**Trial Sponsor: Ravi Amaravadi, MD**

**IND/IDE# : IND exempt**

**Protocol(s) ID#: 842838**

#### DSMB Description

The Data and Safety Monitoring Board (DSMB) for the Prevention And Treatment of COVID-19 with Hydroxychloroquine (PATCH) trial has been selected by the Sponsor-Principal Investigator Dr. Ravi Amaravadi and acts in an advisory capacity to Dr. Amaravadi for the PATCH trial to monitor participant safety, data quality and evaluate the progress of the study.

- This DSMB will consist of 5 members:
  - Dr. John Younger, expertise in Emergency medicine, Chair of DSMB
  - Dr. Keith Hamilton, expertise in infectious disease
  - Dr. Jeffrey Morris: expertise in biostatistics,
  - Dr. Sunita Nasta: expertise in clinical trials and therapeutics
  - Dr. Rajat Deo: expertise in clinical trials and cardiology
- This DSMB will be independent of the Sponsor, regulatory agencies, IRBs/ECs, and investigators.

#### DSMB Membership

##### Requirements for Membership

- Each member declares any conflicts of interest related to the study. Members will notify Sponsor of any change in conflict.
- Each DSMB member maintains confidentiality of DSMB communications.

##### Remuneration

No remuneration will be provided

##### Roles and Responsibilities

The primary charge of the DSMB is to monitor the study for subject safety. Formal DSMB safety reviews occur as outlined in the charter. Additional review may be required. The DSMB may monitor effectiveness outcomes to determine relative risk/benefit, futility, or for early termination due to overwhelming effectiveness.

The primary safety endpoint as well as guidance for the conduct of analyses and guidelines/stopping rules are established in the protocol and are reviewed by the DSMB.

The DSMB may also review data related to study conduct. Data to be reviewed and listed in the DSMB reports may include: enrollment rates over time, time from last subject enrolled to date of report (indication of delay between treatment or follow-up and reporting), summary of protocol violations as specified in monitoring reports generated by the Office of Clinical Research (OCR), completeness of treatment and follow-up visit data, and follow-up duration for the population included in the report. Protocol deviations, SAE reports, and assistance in day to day medical decision making on study participants will be handled by the Medical Monitor Dr. Sunita Nasta who is also a member of the DSMB. The DSMB will in turn be able to review formal reports generated by the OCR monitor and the interim analysis reports generated by the Sponsor-Investigator and his team.

The DSMB:

- Meets as outlined in this charter
- Provides recommendations about continuing, modifying, and stopping the study
- Operates according to the procedures described in this charter

##### Conflicts of Interest

Members comply with the conflict of interest policies specified by University of Pennsylvania to ensure members do not have serious scientific, financial, personal, or other conflicts of interest related to the conduct, outcome, or impact of the study according to the guidelines specified below (e.g., engaged in any simultaneously occurring competitive trials in any role that could pose a conflict of interest for this study).

The DSMB follows 42 C.F.R. Part 50, Subpart F and Responsible Prospective Contractors 45 C.F.R. Part 94.

As determined by the Sponsor, conflicts of interest and/or potential conflicts of interest are mitigated to the greatest extent, consistent with assembling a highly competent DSMB.

#### DSMB Meetings

##### First Meeting

The purpose of the first meeting is to review, discuss, and make recommendations about the Charter and protocol. In addition, a chairperson is elected.

##### Subsequent Meetings

The DSMB meets and reviews unblinded deidentified data provided in the form of a report generated by the sponsor at the timepoints detailed below. The unblinded assignments of subjects may only be shared with the minimum number of study team members needed to generate the report for the DSMB review.
Additional meetings or conference calls are scheduled as needed.
Substudy 1:

1. Substudy 1 Interim analysis1: after 34 out of a target 100  evaluable patients have completed the study and the report is available for review
2. Substudy 1 Interim analysis 2: after 68 out of a target 100 evaluable patients have completed the study and the report is available for review

Substudy 3

1. Substudy 3 Interim analysis1: after 50 out of a target 200  evaluable patients have completed the study and the report is available for review
2. Substudy 3 Interim analysis 2: after 100 out of a target 200 evaluable patients have completed the study and the report is available for review

This DSMB is coordinated by the Sponsor. DSMB meetings are held by teleconference. All three members of the DSMB must be present to hold a DSMB meeting. Critical decisions of the DSMB are made by unanimous vote whenever possible, or majority vote if this is not possible.

While the Medical Monitor will review deviations, unanticipated serious adverse events and safety data, she may consult the DSMB as needed.

##### Meeting Format

DSMB meetings are organized into open and closed sessions. Data reviewed by the DSMB is provided by the Sponsor.

**Open Sessions**
Data presented in the open session may include enrollment data, individual adverse event data, baseline characteristics, overall data accuracy and compliance data or issues, and other administrative data. The DSMB also considers data from other studies or external sources as made available during its deliberations.

During the open sessions, prepared summary reports and tables are reviewed and discussed by the DSMB.

Facilitator(s) (e.g., member of Sponsor and/or investigator team responsible for the report preparation) and observers from University research oversight comittees may attend the DSMB meeting Open Sessions as necessary in order to facilitate data presentation, follow-up reporting, and answer questions posed by the DSMB.

**Closed Sessions**
The closed session is restricted to the DSMB members. Data which may compromise the integrity of the study (e.g., comparative data) is analyzed and discussed only in the closed session. Details of closed session deliberations (e.g., minutes) are considered privileged and not subject to disclosure except as required by law.

Meetings conclude with a recommendation to continue, modify, pause, or terminate the study. In addition, recommendations for modification, pause, or termination may be endorsed for perceived safety concerns based on clinical judgment.

#### DSMB Reports

##### Formal Minutes

Minutes of the open session are recorded and finalized. Minutes are approved by the chairperson and maintained by the Sponsor.

The minutes of the closed session are recorded. They are stored in a secure location by the DSMB chair, separately from the minutes of the open session. Once finalized and approved by the DSMB chair, they will be retained until the trial is completed.

##### DSMB Communication of Recommendations

A formal report containing the recommendations of the study is sent to the Sponsor.
If the DSMB recommends continuation of the study, the minutes and report for continuation are shared with the Sponsor no later than 3 business days after the meeting.

If the DSMB recommends modifications, pause, or termination of the study, the minutes and report are shared with the Sponsor no later than 1 business days after the meeting.

**In addition, any findings that are considered to be serious and potentially consequential that require immediate action are promptly shared with the Sponsor.**

##### Sponsor’s Response to DSMB Recommendations

The Sponsor reviews and responds to the DSMB recommendations.

#### DSMB Closeout

This study may be terminated under a variety of circumstances including, but not limited to, termination for overwhelming effectiveness, futility, or safety issues per protocol, poor enrollment, or DSMB monitoring guidelines.

#### Confidentiality

All data provided to the DSMB and all deliberations of the DSMB are privileged and confidential. The DSMB agrees to use this information to accomplish the responsibilities of the DSMB and will not use it for other purposes without written consent from the Sponsor.

**The PATCH (Prevention And Treatment of COVID-19 with HCQ) Trial:**
**A 3 Arm Randomized Trial of Hydroxychloroquine in the Prevention and Treatment of COVID-19**

| | |
|---|---|
| Sponsor-Investigator/<br>Principal Investigator: | Ravi Amaravadi, MD (Medicine)<br>University of Pennsylvania Perelman School of Medicine<br>Department of Medicine, Division of Hematology Oncology<br>852 BRB 2/3 421 Curie Blvd,<br>Philadelphia, PA 19104<br> |
| Primary Emegrency Medicine Sub-Investigator | Benjamin Abella, MD (Emergency Medicine)<br>University of Pennsylvania Perelman School of Medicine<br>Department of Emergency Medicine<br>Hospital of the University of Pennsylvania<br>Emergency Department, Ground Ravdin<br>3400 Spruce Street<br>Philadelphia, PA 19104<br> |
| Primary Infectious Diseases Sub-Investigator | Ian Frank, MD<br>502 Johnson Pavilion<br>3610 Hamilton Walk 6073<br>Philadelphia, PA 19104-6073<br> |
| Other Significant<br>Sub-Investigators | Anish Agarwal, MD (EM)<br>Julie Uspal, MD (EM)<br>Ari Friedman, MD (EM)<br>John Greenwood, MD (EM)<br>Felipe Teran, MD (EM)<br>Michael Milone, MD PhD (Pathology)<br>William Short, MD (ID)<br>Keith Hamilton, MD (ID)<br>Kathleen Degnan, MD (ID)<br>Pablo Tebas, MD (ID)<br>E. Paul Wiletyo (Statistics)<br>Phyllis Gimotty, PhD (Statistics)<br>Joseph Carver, MD (Cardiology)<br>Dasha Babushok, MD (Oncology)<br>Saar Gill, MD (Oncology)<br>IvanMaillard Md PhD (Oncology)<br>Alexander Huang, MD (Oncology)<br>Dan Vogl, MD (Oncology)<br>Kara Maxwell, MD (Genetics) |
| Medical Monitor: | Sunita Nasta, MD<br>University of Pennsylvania Perelman School of Medicine<br>Department of Medicine, Division of Hematology Oncology<br><br>215-605-2493 |
| Funding: | Internal Funding |

| | |
|---|---|
| Protocol Number: | IRB: 842838 |
| Clinical Trial Management System | Penn-CTMS (Velos) |
| Investigational Agents: | Hydroxychloroquine/Placebo |
| IND #: | IND exempt (Granting entity University of Pennsylvania, Office of Clinical Research (OCR)) |
| Version: | 4.2.20 |

**PRINCIPAL INVESTIGATOR SIGNATURE**

| | |
|---|---|
| SPONSOR-INVESTIGATOR: | Ravi Amaravadi, MD |
| STUDY TITLE: | The PATCH (Prevention And Treatment of COVID-19 with HCQ) Trial: A 3 Arm Randomized Trial of Hydroxychloroquine in the Prevention and Treatment of COVID-19 |
| STUDY ID | IRB #842838 |
| PROTOCOL VERSION | INITIAL VERSION (04.02.2020) |

I have read the referenced protocol. I agree to conduct the study in accordance to this protocol, in compliance with the Declaration of Helsinki, Good Clinical Practices (GCP), and all applicable regulatory requirements and guidelines.

| ______________________________ | ______________________________ |
|---|---|
| Principal Investigator Name | Signature |
| ______________________________ | ______________________________ |
| Affiliation | Date |

**Abbreviations:**

Ab: antibody
AE: adverse event
ALT: alanine aminotransferase
ANC: absolute neutrophil count
AST: aspartate aminotransferases
BID: twice daily
BP: blood pressure
BSA: body surface area
CNS: central nervous system
CT: computed tomography
CTCAE: Common Terminology Criteria for Adverse Events
DSMC: Data Safety and Monitoring Committee
eCRF: electronic case report form
EKG: electrocardiogram
FDA: Food and Drug Administration
FFPE: formalin fixed-paraffin embedded
HBV: hepatitis B virus
HCQ: hydroxychloroquine
HCV: hepatitis C virus
HUP: Hospital of the University of Pennsylvania
IHC: immunohistochemistry
IND: investigational new drug
INR: international normalization ratio
IRB: institutional review board
IV: intravenous
LLN: lower limit of normal
LVEF: left ventricular ejection fraction
PPMC: Penn-Presbyterian Medical Center
PPT1: palmitoyl-protein thioesterase 1
PR: partial response
PT: prothrombin time
PTT: partial thromboplastin time
SAE: serious adverse event
SUSAR: Suspected, Unexpected Serious Adverse Reaction
ULN: upper limit of normal
WBC: white blood cell

PRINCIPAL INVESTIGATOR SIGNATURE ........ 3
1.0 OBJECTIVES ........ 11
1.1 PRIMARY OBJECTIVE ........ ERROR! BOOKMARK NOT DEFINED.
1.2 SECONDARY OBJECTIVES ........ ERROR! BOOKMARK NOT DEFINED.
1.3 CORRELATIVE OBJECTIVES ........ ERROR! BOOKMARK NOT DEFINED.

Study Summary

| | |
|---|---|
| **Title** | The PATCH (Prevention And Treatment of COVID-19 with HCQ) Trial: A 3 Arm Randomized Trial of Hydroxychloroquine in the Prevention and Treatment of COVID-19 |
| **Phase** | Phase II |
| **Methodology** | A prevention and treatment trial with multiple sub-studies including randomized double-blind placebo controlled sub-studies and open label randomized sub-study. |
| **Study Duration** | 1 year |
| **Study Center(s)** | Single Institution: University of Pennsylvania (HUP and PPMC) |
| **Objectives** | **1.1 <u>PRIMARY OBJECTIVES</u>**<br>Sub-Study 1 (home quarantined COVID-19 patients): Median time to release from quarantine by meeting the following criteria: 1) No fever for 72 hours without the use of fever-reducing medications 2) improvement in other symptoms and 3) 7 days have elapsed since the beginning of symptom onset.<br>Sub-Study 2 (hospitalized COVID-19 patients): Rate of hospital discharge at 14 days<br>Sub-Study 3 Physicians and nurse prophylaxis: Rate of COVID-19 infection at 2 months<br><br>**<u>1.2 SECONDARY OBJECTIVES</u>**<br>Sub-Study 1: Rate of participant-reported secondary infection of housemates, rate of hospitalization, rate of treatment related adverse events<br>Sub-Study 2: Time to condition appropriate for discharge, rate of ICU admission, time to PCR negativity, rate of treatment related adverse events<br>Sub-Study 3: Number of scheduled shifts missed, rate of treatment related adverse events<br><br>**<u>1.3 CORRELATIVE OBJECTIVE</u>**<br>To bank serially collected samples to enable correlative science related this trial |
| **Number of Subjects** | Sub-Study 1: 100; interim analysis after 34 and 68<br>Sub-Study 2: 100; interim analysis after 34 and 68<br>Sub-Study 3: 200; interim analysis after 50 and 100<br>**Total: 400** |
| **Diagnosis and Main Inclusion/Exclusion Criteria** | • Age ≥ 18 years old (Sub-studies 2 and 3)<br>• Competent and capable to provide informed consent |

| | |
|---|---|
| | • Have access to a smart device such as a cell phone, tablet, laptop computer with necessary data/internet accessibility<br>• Subjects meeting the following criteria by Sub-Study<br>Sub-Study 1:<br>• Age ≥40 years since the risk of prolonged disease that progresses to severe COVID-19 disease increases with age.<br>• PCR-positive for the SARS-CoV2 virus<br>• (Fever, and cough, or Fever and shortness of breath,<br>• ≤4 days since the first symptoms of COVID-19 and date of testing<br>• Not taking azithromycin<br>• Not requiring hospitalization and is sent home for quarantine.<br>• Must live within 30 miles of HUP or Penn Presbytarian Medical Center to facilitate drop-off of medication<br>• Must own a working computer, or smartphone and have internet access<br>• Must be willing to fill out a daily symptom diary<br>• Must be available for a daily phone call,<br>• Must take their own temperature twice a day<br>• Must be willing to report the observed symptoms and development of COVID-19 in the co-inhabitants of the residence at which the quarantine will be served.<br><br>Sub-Study 2 Hospitalized non-ICU service patients.<br>• PCR-positive for SARS-CoV-2<br>• Patients admitted to a floor bed at Hospital of the University of Pennsylvania or Penn Presbyterian.<br>• One or more of the following risk factors for progression to severe disease including: immunocompromising conditions, structural lung disease, hypertension, coronary artery disease, diabetes, age > 60, ferritin > 850, CRP > 6, D-dimer > 1000<br>Sub-Study 3 Health Care Worker Prevention<br>• Emergency Medicine or Infectious Disease Team physician or nurse at HUP or PPMC<br>• ≥20 hours per week of clinical work scheduled in the coming 2 months during the COVID-19 pandemic<br>• No fever, cough, or shortness of breath in the past 2 weeks<br><br>• Willing to report compliance with HCQ in the form of a diary<br>• Patients must be able to swallow and retain oral medication and must not have any clinically significant gastrointestinal abnormalities that may alter absorption such as malabsorption syndrome or major resection of the stomach or bowels.<br>**Exclusion Criteria**<br>• <18 years of age<br>• Prisoners or other detained persons<br>• Allergy to hydroxychloroquine |

<table>
<tr><td></td><td>
<ul>
<li>Pregnant or lactating or positive pregnancy test during pre-medication examination</li>
<li>Receiving any treatment drug for 2019-ncov within 14 days prior to screening evaluation (off label, compassionate use or trial related).</li>
<li>Co-enrollment onto another COVID-19 study is not allowed.</li>
<li>Known history of retinal disease including but not limited to age related macular degeneration.</li>
<li>Taking any of the following medications that prolong Qtc: Chlorpromazine.Haloperidol, Droperidol, Quetiapine, Olanzapine. Amisulpride. Thioridazine</li>
<li>History of interstitial lung disease or chronic pneumonitis unrelated COVID-19.</li>
<li>Due to risk of disease exacerbation patients with porphyria or psoriasis are ineligible unless the disease is well controlled and they are under the care of a specialist for the disorder who agrees to monitor the patient for exacerbations.</li>
<li>Patients with serious intercurrent illness that requires active infusional therapy, intense monitoring, or frequent dose adjustments for medication including but not limited to infectious disease, cancer, autoimmune disease, cardiovascular disease.</li>
<li>Patients who have undergone major abdominal, thoracic, spine or CNS surgery in the last 2 months, or plan to undergo surgery during study participation.</li>
<li>Patients receiving cytochrome P450 enzyme-inducing anticonvulsant drugs (i.e. phenytoin, carbamazepine, Phenobarbital, primidone or oxcarbazepine) within 4 weeks of the start of the study treatment</li>
<li>History or evidence of increased cardiovascular risk including any of the following:
<ul>
<li>Left ventricular ejection fraction (LVEF) < institutional lower limit of normal. Baseline echocardiogram is not required.</li>
<li>A QT interval corrected for heart rate using the Frederica formula > 500 msec (Sub-study 2)</li>
<li>Current clinically significant uncontrolled arrhythmias. Exception: Subjects with controlled atrial fibrillation</li>
<li>History of acute coronary syndromes (including myocardial infarction and unstable angina), coronary angioplasty, or stenting within 6 months prior to enrollment</li>
<li>Current ≥ Class II congestive heart failure as defined by New York Heart Association</li>
</ul>
</li>
</ul>
</td></tr>
<tr><td>Study Product, Dose, Route, Regimen</td><td>Hydroxychloroquine, various doses, oral<br><br>Placebo, oral</td></tr>
</table>

| | |
|---|---|
| **Duration of administration** | Sub-Study 1 at home quarantine<br>Arm 1: HCQ 2 weeks 400 mg bid; Arm 2: Placebo 2 pills bid for 2 weeks; crossover allowed upon disease progression or lack of clearance of virus<br>Sub-Study 2 Hospitalized patient: randomized to Arm 1 HCQ 600 mg qd for one week (low dose) versus Arm 2: HCQ 600 mg bid for 2 weeks (high dose).<br>Sub-Study 3 EM or ID team physician or nurse prophylaxis. Arm 1: HCQ 600 mg qd for 2 months versus Arm 2: Placebo 3 pills qd for 2 months; crossover from placebo to HCQ is allowed upon disease progression/ virus confirmation |
| **Study design** | 1:1 randomization in each cohort between the arms 1 and 2 |
| **Duration of trial** | Approximately 1 year |

#### 2.0 OBJECTIVES

##### 2.1 PRIMARY OBJECTIVES

Sub-Study 1 (home quarantined COVID-19 patients): Median time to release from quarantine by meeting the following criteria: 1) No fever for 72 hours without the use of fever-reducing medication 2) improvement in other symptoms and 3) 7 days have elapsed since the beginning of symptom onset.
Sub-Study 2 (hospitalized COVID-19 patients): Rate of hospital discharge at 14 days
Sub-Study 3 Physicians and nurse prophylaxis: Rate of COVID-19 infection at 2 months

##### 1.2 SECONDARY OBJECTIVES

Sub-Study 1: Rate of participant-reported secondary infection of housemates, rate of hospitalization, rate of treatment related adverse events
Sub-Study 2: Time to condition appropriate for discharge, rate of ICU admission, time to PCR negativity, rate of treatment related adverse events
Sub-Study 3: Number of scheduled shifts missed, rate of treatment related adverse events

##### 1.4 CORRELATIVE OBJECTIVE

To bank serially collected samples to enable correlative science related this trial

#### 2 BACKGROUND AND RATIONALE

**2.1 Chloroquine derivatives show preclinical efficacy against coronavirus but very little clinical data is available.** Emerging viral diseases (EVDs) encompass a growing list of zoonotic viruses that have a major impact on global health and economics. These include Ebola, SARS, MERs, Marburg, and the recently identified virus that causes COVID-19 disease (Wuhan Coronavirus: SARS-CoV-2) (1). In each of these cases there is currently no effective drug treatments or prophylaxis agents that can be quickly applied to large populations at risk for the virus. As of March 2020, The SARS-CoV2 virus has infected more than 180,000 people causing more than 5000 deaths. This has led to a lockdown of entire megacities in China, international travel bans, and disruptions in global supply chains. More alarming is the recent evidence that health care workers are getting infected and dying of the virus, wreaking havoc on the chain of care. In addition, while prior epidemics such as Ebola seemed to be contained in Africa, the current COVID-19 global crisis demonstrates how with the extent of globalization and international travel, no country is safe from these EVDs.

Coronaviruses are a large family of viruses that commonly infect many animals, including camels, cattle, cats, and are often found in bats as their zoonotic reservoir. While it is rare, animal coronaviruses can infect people and then spread between people, as had occurred with Middle East respiratory coronavirus (MERS-CoV) and severe acute respiratory coronavirus (SARS-CoV), and now the newly emerged coronavirus SARS-CoV-2 (also known as COVID-19). The SARS-CoV-2 virus is a betacoronavirus, like MERS-CoV and SARS-CoV. All three of these viruses have their origins in bats. While SARS-CoV and MERS-CoV exhibit significantly higher mortality than SARS-CoV-2, the ability to spread between humans is less than SARS-CoV-2. There is emerging evidence from Asia that patients can be highly contagious with one patient spreading viral particles to 87% of 15 sites within the patient's room. Virus can be shed in urine and feces as well as respiratory secretions increasing the likelihood that sick contacts and hospital staff will become infected (2). An epidemiological survey of 72,000 cases in China found and overall 3.8% infection rate in hospital workers, but a 68% infection rate of hospital workers in Wuhan at the epicenter of disease. This suggests that if low rates of infection are in a community, the likelihood of hospital workers contracting disease is low, but if there are a large number of cases then the likelihood of hospital workers contracting the disease is high (3). Patients above the age of 50 are more likely to die of this disease and the time from diagnosis to death can occur within one month despite access to high-level intensive care units (4). A high rate of intra-family transmission and rapid community transmission has been documented in Shenzen China (5). Meanwhile a screen of compounds found that chloroquine prevented and irradicated established infection of SARS-CoV-2 virus in vitro. The Chinese and Korean consensus guidelines for COVID-19 treatment include chloroquine. However, based on our experience in cancer clinical trials we believe hydroxychloroquine is a safer drug than chloroquine and affords the ability to dose escalate to concentrations that we know are effective at blocking the lysosome in patients. A recent study found that hydroxychloroquine produced rapid clearance of virus in hospitalized patients with COVID-19 (Gautret et al Int. J Antimicrobial Agents 2020 in press).

Our ***hypothesis*** is that high doses of hydroxychloroquine for at least 2 weeks can be effective antiviral medication both as a treatment in hospitalized and ambulatory patients and prophylaxis in health care workers because it impairs lysosomal function and reorganizes lipid raft (cholesterol and sphingolipid rich microdomains in the plasma membrane) content in cells, which are both critical determinants of EVD infection. This hypothesis is based on a growing literature linking chloroquine to antiviral activity and our own work in the field. We believe there is enough information to launch a clinical trial at the Hospital of the University of Pennsylvania of hydroxychloroquine for COVID-19

##### *2.2 Mechanistic rationale for antiviral properties of chloroquine derivatives.*

**2.2a Coronaviruses.** Coronaviruses (CoV) infect mammals and birds causing respiratory, gastrointestinal, and central nervous system diseases. Coronavirus virions contain an envelope, a helical capsid, and a single-stranded RNA genome, the largest among all RNA viruses. The name "coronaviruses" derives from the spike proteins on their envelope that give the virions a crown-like shape. CoVs include the following: 1) α-genus: human coronavirus NL63 (HCoV-NL63), porcine transmissible gastroenteritis coronavirus (TGEV), and porcine respiratory coronavirus (PRCV) 2) β-genus: SARS-CoV-2/COVID-19, severe acute respiratory syndrome coronavirus (SARS-CoV), Middle East respiratory syndrome coronavirus (MERS-CoV), mouse hepatitis coronavirus (MHV), and bovine coronavirus (BCoV). 3) CoVs of the γ-genus include avian infectious bronchitis virus (IBV) and do not infect humans so are valuable laboratory agents (6). Coronaviruses impose health threats to humans and animals. SARS-CoV caused the SARS epidemic in 2002 to 2003, with over 8,000 infections and a fatality rate of ~10%. MERS-CoV emerged from the Middle East in 2012 causing 877 infections with a fatality rate of ~36%. HCoV-NL63 from the α-genus is a widespread pathogen that produces the common cold in healthy adults and acute respiratory illness in young children. CoVs are also major animal pathogens. TGEV and MHV cause close to 100% fatality in young pigs an young mice, respectively; BCoV and IBV also cause significant healthcare burden for domesticated cattle and chickens, respectively. Therefore, research on coronaviruses has strong health and economic implications (6).

**2.2b CoV infection requires functional lysosomes.** Receptor recognition by viruses is the first and essential step of viral infections of host cells. An envelope-anchored spike protein mediates coronavirus entry into host cells by first binding to a receptor on the host cell surface and then fusing viral and host membranes. CoVs recognize a number of different host receptors. Once the receptor is bound, there is viral and cell membrane fusion, followed by endocytosis and lysosomal processing. CoVs like SARS enter the cell through lipid raft enriched endocytosis (7). The details of the interaction between the virus spike protein, virus membrane and the lipid rafts are not worked out. In the lipid raft literature, it is often the case that lipid rafts (semisolid phase membrane filled with cholesterol and sphingolipids) are only physiologically functional if there is a non-lipid raft membrane region next to it. After membrane fusion, endosomal pH acidification is a fusion trigger for CoVs and other viruses including influenza virus and vesicular stomatitis virus (VSV). For instance the use of lysosomal inhibitor blocked the entry of the model coronavirus IBV (8). The authors directly assessed the pH dependence of IBV fusion and found that fusion only occurs at acidic pH. For some CoVs that harbor a non-cleaved spike protein on their surface, such as MHV-2 and SARS-CoV, it has been shown that they rely on lysosomal proteases for productive entry. In fact one study found that bat cells have more efficient lysosomal proteolysis than human cells providing an explanation for why bats are preferred zoonotic hosts for CoVs (9).

**2.2c Evidence that chloroquine prevents coronavirus infection.** It has been reported that chloroquine has strong antiviral effects on SARS-CoV infection and spread in vitro (10-12). In to increasing endosomal/lysosomal pH, these studies demonstrated that CQ abrogates glycosylation of ACE2, the cellular receptor of SARS-CoV, which may contribute further to infection suppression. The IC50 of chloroquine for inhibition of SARS-CoV in vitro is 8.8 micromolar which would require higher concentrations than typically delivered in malaria. Importantly, the suppressive effects are observed when the cells are treated with chloroquine either before or after exposure to the virus, suggesting both prophylactic and therapeutic treatment paradigms could be employed (10, 11). The work by Kayaerts looked at human coronavirus OC43 which causes neonatal death in mice. Treatment with chloroquine of pregnant mothers provided a 98.6% protection against death in newborn mice. This is in the only in vivo demonstration of chloroquine efficacy against a CoV. DeWilde et al. screened a library of 348 FDA-approved drugs for anti- MERS-CoV activity in cell culture and only four compounds (chloroquine, chlorpromazine, loperamide, and lopinavir) were found to inhibit the viral replication (50% effective concentrations, EC50 3–8 lmol/L) (12). The protective activity of chloroquine against CoVs such as SARS, MERS and COVID-19 has not yet been established in animal models.

**2.2d Chloroquine is active against COVID-19/ SARS-CoV-2:** A recent paper from Wuhan China isolated SARS-CoV2 (the virus that causes COVID-19 disease) and tested 6 modern antiviral drugs and chloroquine and found that the best suppressors of viral infection were remdesivir (IC50 0.77 μmol) and chloroquine (IC50 1 μmol) (13). Only chloroquine given at 10 μM was able to suppress infection either with pretreatment, at the time of infection, or after infection occurred. This has led to clinical trials and off label use of chloroquine in China (Dr.

Amaravadi personal communication with scientists in China). There are Chinese guidelines for the use of chloroquine 500 mg bid for the treatment of COVID-19.

**2.2e Evidence for immunopathology in severe CoVs infection**. Although the mechanisms of pathology in severe SARS and MERS CoV infections are not well understood, the severe lung damage observed in SARS patients is associated with high viral loads during the early phase of infection and abundant macrophage-monocyte and neutrophil accumulation in the lungs (14). Dysregulated innate and adaptive immunity have been postulated to contribute to severe CoV disease. The acute lung injury of severe CoV infection is associated with elevated serum pro-inflammatory cytokines. In particular, fatal SARS is associated with a high and persistent expression of interferon (IFN) and IFN-responsive genes with evidence for impaired adaptive immunity, most notably reduced anti-Spike neutralizing antibody responses (15). In a murine model of SARS, disruption of type I IFN signaling or monocyte-macrophage depletion protected mice from lethal infection supporting a role for dysregulated type I IFN expression in CoV pathology (16).

**2.2f Chloroquine modulates type I interferon expression and T cell immune responses**. The endosomal compartment plays a central role in immunobiology, especially in innate pathogen sensing as well as antigen presentation. Toll-like receptors that recognize virally-associated nucleic acid are localized to the endosome of monocytes and specialized DC populations including the plasmacytoid DC (pDC), which are a major producer of type I interferons (17). HCQ is able to disrupt this endosomal sensing pathway to reduce type I interferons production by these cells, and may attenuate excessive type I interferon expression that has been postulated to contribute to severe CoV infection. HCQ has also been reported to modulate endosomal membrane permeability to enhance cross-presentation of endosomal antigens via MHC class I, which enhances adaptive CD8+ T cell immunity.

2.3 **Clinical experience with high dose hydroxychloroquine in cancer patients.** In contrast to CQ, which can produce blindness at high cumulative doses, our recent body of work has demonstrated that high dose HCQ can be administered safely to humans for months. Our work on autophagy as a resistance mechanism to cancer therapy had identified CQ derivatives as potential anti-cancer agents (18). Since HCQ has had a longer track record of dose escalation and chronic dosing in rheumatoid arthritis and lupus, we chose HCQ as our lysosomal autophagy inhibitor to test in combination regimens in cancer patients. We reported the first 6 phase I dose escalation clinical trials involving HCQ in combination with FDA approved anti-cancer drugs in refractory stage IV cancer patients (19-24). In most patients on these trials we were able to escalate the dose of HCQ to 1200 mg per day and dose patients in some cases safely for more than one year. In over 220 patients treated across multiple studies the rate of grade 3-4 non-hematological toxicities was < 10%. Most of these had more to do with the severity of the cancer and would likely not be seen in a healthy population. Specifically, no retinal, cardiac, neurological, hepatic or renal toxicity was observed. Hematological toxicities could be attributed to the other cancer drug (chemotherapy) that HCQ was paired with, and the most common side effects included manageable gastrointestinal symptoms symptoms such as bloating, diarrhea, constipation and mild non-bloody diarrhea.

2.4 **Low dose versus high dose HCQ.** In HCQ studies in cancer patients we conducted population pharmacokinetic studies and pharmacodynamic studies and determined that 800-1200 mg daily was required to effectively and consistently impair the lysosome in peripheral blood mononuclear cells. Our PK studies determined that it took roughly 2 weeks to achieve steady state in cancer patients. For these reasons there is rationale to propose a high dose (600 mg po bid) prolonged schedule treatment (2 weeks). In contrast an in vitro study recently published which included PK-PD modeling extrapolated from published PK studies indicated that HCQ was very active against COVID-19 but only 400 mg po bid X1 followed by 200 mg po bid for 5-10 days was enough for treatment. A recent non-randomized open label French trial (Gautret et al Int. J Antimicrobial Agents 2020) showed that 600 mg qd of HCQ cleared virus in 70 % of mildly ill hospitalized patients compared to 12.5% at 6 days of control patients. This generates a major question in how to use this agent in different populations that can best be answered in this three-part placebo controlled randomized trial.

#### 3.0 ELIGIBILITY

##### 3.1 Inclusion Criteria

3.1.1 Age ≥ 18 years old, except sub-study 1
3.1.2 Competent and capable to sign informed consent
3.1.3 Subjects meeting the following criteria by arm:
3.1.3 Sub-Study 1:

- Age ≥40 years since the risk of prolonged disease that progresses to severe COVID-19 disease increases with age.
- PCR-positive for the SARS-CoV2 virus
- Fever and cough OR Fever and shortness of breath
- ≤4 days since the first symptoms of COVID-19 and date of testing
- Not requiring hospitalization and is sent home for quarantine.
- Not taking azithromycin
- Must live within 30 miles of HUP or Penn Presbytarian Medical Center to facilitate drop-off of medication
- Must own a working computer, or smartphone and have internet access
- Must be willing to fill out a daily symptom diary
- Must be available for a daily phone call,
- Must take their own temperature twice a day
- Must be willing to report the observed symptoms and development of COVID-19 in the co-inhabitants of the residence at which the quarantine will be served.

Sub-Study 2 Hospitalized non-ICU service patients.

- PCR-positive for SARS-CoV-2
- Patients admitted to a floor bed at Hospital of the University of Pennsylvania or Penn Presbytarian. Ward service patients that are housed in non-ward beds will not be disqualified.
- One or more of the following risk factors for progression to severe disease including: immunocompromising conditions, structural lung disease, hypertension, coronary artery disease, diabetes, age > 60, ferritin > 850, CRP > 6, D-dimer > 1000

Sub-Study 3 Health Care Worker Prevention

- Emergency Medicine or Infectious Disease Team physician or nurse at HUP or PPMC
- ≥20 hours per week of clinical work scheduled in the coming 2 months during the COVID-19 pandemic
- No fever, cough, or shortness of breath in the past 2 weeks

3.1.4 Sub-studies 1 and 3: Willing to report compliance with HCQ in the form of a diary
3.1.5 Sub-Studies 1 and 3: Participants are willing to collect oral fluid using a simple applicator that is provided, and freeze these samples.
3.1.6 Not pregnant and/or breastfeeding, as determined below:

Sub-Study 2: Negative serum or urine pregnancy test within 14 days prior to commencement of dosing in premenopausal women. Women of non-childbearing potential may be included without serum or urine pregnancy test if they are either surgically sterile or have been postmenopausal for ≥ 1 year.

Sub-Study 1 and 3: Not known to be pregnant or breast feeding as determined through self-report

[All participants] 3.1.7 Patients must be able to swallow and retain oral medication and must not have any clinically significant gastrointestinal abnormalities that may alter absorption such as, but not limited to malabsorption syndrome, major resection of the stomach or bowels, gastric bypasss, lap banding.
3.1.8 Sub-study 2: Patients must have adequate baseline organ function as determined by **Table 1**.

**Table 1. Definitions for adequate baseline organ function**

| System | Laboratory Values |
|---|---|
| **Hematologic** | |
| Absolute neutrophil count (ANC) | ≥ 1.0 × $10^9$/L |
| Hemoglobin | ≥ 9 g/dL |
| Platelet count | ≥ 100 x $10^9$/L |
| PT/INR[a] and PTT | ≤ 1.3 x ULN |
| **Hepatic** | |
| Total bilirubin[b] | ≤ 1.5 x ULN |
| AST and ALT | ≤ 2.5 x ULN |
| **Renal** | |
| Serum creatinine[c] | ≤ 1.5 mg/dL |

Abbreviations: ALT = alanine transaminase; ANC = absolute neutrophil count; AST = aspartate aminotransferase; INR = international normalized ratio; LLN = lower limit of normal; PT = prothrombin time; PTT = partial thromboplastin time; ULN = upper limit of normal.

[a] Subjects receiving anticoagulation treatment may be allowed to participate with INR established within the therapeutic range prior to enrollment.

[b] Except subjects with known Gilbert's syndrome.

[c] If serum creatinine is > 1.5 mg/dL, calculate creatinine clearance using standard Cockcroft-Gault formula. Creatinine clearance must be ≥ 50 mL/min to be eligible.

**3.2 Exclusion Criteria**

3.2.1 <18 years of age [Sub-Studies 2 and 3]; <40 years old [Sub-Study 1]
3.2.2 Prisoner or other detained person
3.2.3 Allergy to hydroxychloroquine, 4 aminoquinolines, or quinine
3.2.2 Patients with known history of G6PD deficiency
3.2.2 Pregnant and/or breastfeeding, see above
3.2.3 Receiving any treatment drug for 2019-ncov within 14 days prior to screening evaluation (off label, compassionate use or trial related).
3.2.4 Co-enrollment onto another COVID-19 study is not allowed.
3.2.5 Known history of retinal disease, including but not limited to, macular degeneration, retinal vein occlusion, visual field defect, diabetic retinopathy
3.2.6 Known history of interstitial lung disease, severe emphysema or asthma, or chronic pneumonitis unrelated COVID-19.
3.2.7 Taking any of the following medications that prolong Qtc: Chlorpromazine.Haloperidol, Droperidol, Quetiapine, Olanzapine. Amisulpride. Thioridazine

3.2.8 Due to risk of disease exacerbation patients with known porphyria or psoriasis are ineligible unless the disease is well controlled and they are under the care of a specialist for the disorder who agrees to monitor the patient for exacerbations.

3.2.6 Patients with serious intercurrent illness that requires active infusional therapy, intense monitoring, or frequent dose adjustments for medication including but not limited to infectious disease, cancer, autoimmune disease, cardiovascular disease.

3.2.7 Patients who have undergone major abdominal, thoracic, spine or CNS surgery in the last 2 months, or plan to undergo surgery during study participation.

3.2.8 Patients receiving cytochrome P450 enzyme-inducing anticonvulsant drugs (i.e. phenytoin, carbamazepine, Phenobarbital, primidone or oxcarbazepine) within 4 weeks of the start of the study treatment

3.2.9 History or evidence of increased cardiovascular risk including any of the following:

- Left ventricular ejection fraction (LVEF) < institutional lower limit of normal. Baseline echocardiogram is not required.
- A QT interval corrected for heart rate using the Frederica's formula > 500 msec (for Sub-study 2 only; hospitalized patients; for Sub-study 1 and Sub-study 3, as outpatients routine EKGs are not indicated for starting HCQ)
- Current clinically significant uncontrolled arrhythmias. Exception: Participants with controlled atrial fibrillation
- History of acute coronary syndromes (including myocardial infarction and unstable angina), coronary angioplasty, or stenting within 6 months prior to enrollment
- Current ≥ Class II congestive heart failure as defined by New York Heart Association

#### 4.0 PARTICIPANT RECRUITMENT, CONSENT, AND REGISTRATION

##### 4.1 Sub-study –specific flow of recruitment, consent, registration, drug supply

4.1.1 <u>Sub-Study 1 Outpatient Recruitment, Consent and Registration Procedure:</u>

1. Patients with a positive SARS-CoV-2 test are identified by a clinical lab. The sites for testing for Sub-Study 1 are the EM influenza-like illness (ILI) tents outside of HUP and PPMC.
   . In general patients that meet minimal screening criteria for wellness get swabbed and discharged from the tent to home. Patients that have more severe symptoms and need further care are directed to the ED and do not undergo testing in the tent. Patients seen in the ED and discharged home that end up positive by PCR can be approached for participation in the study.
   For this substudy, EM doctors will screen for patients ≥40 with Fever and cough, or Fever and shortness of breath AND symptoms started 4 days or less before the patient came for testing.

a. For these selected patients ER docs in Presby and HUP ED tents will order a study-specific SARS-CoV-2 test in EPIC that maps to LDT testing at Quest in Cerner. This orderable will be added to Quick pick lists in the HUP and Presby ED facilities only.
b. The samples collected on potential study subjects will be stored in a dedicated cooler on site in the tent separate from the regular COVID-19 testing samples. The use of the unique test oderable appearing on the label will also distinguish these from the normal COVID-19 test specimens.
c. HUP-Presby courier will pick up these study specimens from ER tents and bring them directly to HUP CR&P.

d. CR&P will receive and package these study-related specimens for FEDEX to be sent directly to Quest's Chantilly lab with information in/on the packaging o direct them to a supervisor at Chantilly for priority processing. Order information for these specimens will flow through Cerner interface to Quest.
e. Quest will priority process the specimens on their LDT platform and result.
f. Result from Quest will return to Cerner via interface
g. Result in Cerner will flow to EPIC PennChart under study-specific test name, which will be created to make it apparent that it is a SARS-CoV-2 test.

2. Delegated study team member will be notified of  positive patients t (as example: clinical research coordinators (CRC)).
3. Study team member screens the EPIC chart for specific eligibility criteria and calls the patient (if appropriate)
    a. Informs the patient of positive SARS-CoV-2 result
    b. Introduces the study and determines if the patient is interested in hearing more details from a clinician investigator
    c. If interested, study team member sends consent via email/DocuSign to interested patient
4. Clinical Investigator contacts the interested patient
    a. Conducts e-consent process with the patient and documents via telemedicine/DocuSign
    b. Screens the participant for signs and symptoms that would warrant hospitalization (dizziness, shortness of breath at rest, orthostasis, low urine ouput).
    c. Completes the eligibility checklist and signs it.

6. CRC links EPIC chart to study and assigns a subject ID number
5. Clinical Investigator uses PennChart to order the study drug and includes subject study number
6. IDS randomizes the participant
7. IDS prepares blinded medication in bottles and includes 3 oral fluid collector applicators
8. Blinded study drug (HCQ or placebo) is dispensed to CRC who couriers it to the driver of the EM van for delivery to the participant's house. The patient must call the CRC to inform receipt o study drug. CRC documents this receipt. If for some reason this is not possible, drug will be shipped to the participant using an approved pharmacy shipping vendor.
9. Appropriately delegated and credentialed study team member (as example: study nurse) calls the participant and reviews use of the study drug, and establishes best contact information for response monitoring (See Section 8.5)

4.1.2 Sub-Study 2 Hospitalized patients

1. Inpatient registration can occur in the emergency room or the hospital inpatient floor
2. ID team notifies study team member (as example: CRC) that a patient is admitted to hospital
3. Appropriately delegated and credentialed clinical investigator econsents patient using docusign.
4. Baseline research blood draw is coordinated with nursing/phlebotomy for routine inpatients labs
5. Research blood is then obtained by an appropriately delegated study team member and brought to TTAB for correlative sample processing and storage
6. Baseline lab data and EKG recorded in EMR
7. Baseline CXR findings recorded in EMR
8. Appropriately delegated and credentialed clinical investigator orders study drug through PennChart
9. Participant is randomized to open label low versus high dose HCQ by IDS
10. Inpatient IDS courier delivers the medication bedside
11. Appropriately delegated study team member (as example: CRC) collects clinical and laboratory data each day

4.1.3 Sub-Study 3 Outpatient Hospital Workers

1. Healthcare workers that are part of EM or ID divisions will be invited to fill out a survey to express interest in this study that is sent by email by the University.
2. Based on this survey, Healthcare workers are instructed to contact an appropriately delegated and credentialed clinical investigator of the alternate division (see section 4.2).
3. Delegated and credentialed clinical investigator sends consent via email/DocuSign and reviews consent form with the participant
4. Conducts e-consent process with the patient and documents via telemedicine/DocuSign
5. If a participant is a member of the PATCH study team, then they are counselled to ensure they do not reveal their own participation during the consenting of participants to Sub-study 1 and 2.
6. Eligibility checklist is reviewed and signed by an appropriately delegated and credentialed clinical investigator
7. Nasopharyngeal swab for COVID19, and blood collection if available/possible is performed in the designated area
8. Participant is randomized by IDS
9. Blinded study drug and 5 oral fluid collection kits are provided directly to the participant by IDS in Maloney building basement. Research pharmacist reviews dosing of study drug (three pills daily) with the participant.

##### 4.2 Safeguards against coercion and bias during the recruitment and consenting

4.2.1 Sub-study 1: To ensure there is ample time for the participant to consider participating, they will be provided with the informed consent form through Docusign which includes a detailed summary of the study at the time the study team reaches out to the potential participant to report positive test findings, and gauge general interest in the study. When the participant comes to the EM tent for screening labs, an appropriately delegated and credentialed clinical investigator from the EM Department will review the consent form with the individual and address any questions.
4.2.2 Sub-Study 2: Potential participants will be approached by an appropriately delegated and credentialed clinical investigator from the ID team to consent to participation. The physician investigator will review the e-consent form with the potential participant using telemedicine. The study team will give the participant time to independently review the information in the consent form and discuss with family/friends. The research team will be available to address any of the participant's questions/concerns. Since these participants will be in isolation, the initial version of the consent document could be provided via e-mail/text for a patient to review on their phone or smart device, with documentation obtained through Docusign. For participants who are not technologically astute, paper copy of the consent form can be reviewed with appropriate personal protective equipment. When using a paper consent process, the study team will take a digital image of the signed paper consent signature page, and insert the primary document in a sealed plastic bag and spray the bag with 70% ethanol to sterilize the outside of the bag. This will allow the paper document to be transported to a central office for safekeeping while minimizing the spread of virus. The digital picture of the signature lines will be used by the study team as signal of executed consent process while the paper version decontaminates.

4.2.3 Sub-study 3: Recruitment will occur through a survey which is sent by the University asking all of the healthcare workers that are employed by ID or EM to respond if they are interested in participation. This initial introduction to the study will not be made by any employee's direct supervisor or a person in the supervisory hierarchy. This is intended to minimize any feelings of undue influence in recruiting amongst the UPHS healthcare workforce allowing for autonomy in decisions to participate.

*Safeguards against coercion:* UPHS employees who are under the direct supervision of the study PI and/or sub-Investigators are at an increased risk for undue coercion to participate and considered to be a vulnerable population. Assuming that Emergency Medicine and Infectious Disease study team physicians and study team nurses will enroll into Sub-study 3 as participants, a physician investigator from the alternate department will consent the UPHS worker onto this sub-study. For example, if a study physician in EM wants to enroll, the physician investigator obtaining consent would then be from ID. The delegated physician investigator that consents the participant in this sub-study will remind the participant that the decision about research participation will not affect (favorably or unfavorably) performance evaluations, career advancement, or other employment-related decisions made by peers or supervisors. Documentation of these conversations will be recorded as part of the informed consent process and provided in the informed consent document. EM and ID division leadership will be educated for awareness that they may not utilize enrollment in making any decisions about staff performance or advancement. These measures are put into place to minimize any undue coercion and allow for autonomy in decisions to participate. The PI, Primary ID Sub-Investigator Contact and/or Primary EM Sub-Investigator Contact will not enroll to participate personally as participants in the research, this will ensure that there is always at least one non-participating clinical investigator for study operations and ensure the integrity of the data analysis remains intact (unbiased).

*Safeguards against bias for physicians with dual roles:* Any PATCH trial sub-investigator physician (or study team member) who is enrolled as a participant into Sub-study #3 will be removed from certain elements of the study conduct to minimize bias. Such a person with a dual role as both a trial participant and trial study team member:
a. would not be able to perform data entry, review, evaluations associated with their own data; data should be entered, reviewed, and evaluated by a member from the alternate division
b. would not be able to perform data analysis, if the analysis is identifiable; aggregate analysis could be partially allowable in certain circumstances
c. would have some restrictions in performing informed consent for patients enrolling into Sub-study #1 and Sub-study #2—as above, the physician investigator would not be allowed to mention to the prospective patient being consented that the physician was also participating in the study--- minimizing bias of safety and therapeutic benefit (‘if my doctor enrolled, then it’s safe and so should I’).

##### 4.3 Consenting process

4.3.1 <u>Substudy 1, 2, and 3</u>: Informed consent will be performed via Docusign, an electronic mechanism that is both HIPAA and 21 CFR Part 11 compliant. It is important for infection control and research staff safety to use e-consent approaches; use of paper with wet signatures represents an excessive risk of contaminated fomites. Appropriately delegated and credentialed physician investigators will conduct the consent process and answer any questions from participants via telephone. The e-consent will be obtained via the participant’s smartphone/smart device.

4.3.2 <u>Substudy 3:</u> For this sub-study 3, consent forms will include additional details relevant to healthcare workers as a vulnerable population, ensuring (a) that any medical data collected will not be shared with Penn managers or affect employment, (b) that agreement or refusal to participate will not impact employment or be included in any personnel files, and that (c) physician sub-investigators obtaining consent are not direct supervisors of any participants undergoing enrollment.

##### 4.3 Required Registration information

- Patient’s initials (first and last)
- Patient’s Hospital ID and/or last 4 digits of Social Security number
- Patient past medical history
- Smoking status Current former
- Gender
- Birth date (mm/yyyy)
- Race

- Ethnicity
- Nine-digit ZIP code
- State of residence
- County of residence
- Cell phone number and alternate numbers
- Email address
- Sub-study 1: A description (not containing PHI) for all full time inhabitants at the residence where the participant will stay during the 14-day quarantine.

#### 5.0 RANDOMIZATION

After all applicable screening assessments have been performed, participants who have met all inclusion criteria and none of the exclusion criteria will be centrally, randomly allocated to one of the two groups in each sub-study. Randomization will be done using computer generated randomization numbers. Treatment **should start within 3 working days after randomization.** Once the order for study drug arrives at the pharmacy and the participant is randomized only the pharmacist is aware of the assignment of HCQ versus placebo in the double blind sub-studies (1 and 3).

**Blinding:** Sub Study 1 and 3 will have a double-blind placebo controlled design. Sub-Study 2 is open label randomized study.

**Unblinding:** Sub-Study1: If a participant progresses with symptoms of COVID-19 based on the assessment scale outlined below an appropriately delegated and credentialed study team member will review the case with a clinical investigator who will sign off on the need to un-blind the participant. If the participant was on placebo and continues to have symptoms, then he/she will be allowed to crossover to the HCQ dosing. The EM van will deliver the HCQ to the participant. If the patient was on HCQ then HCQ will be stopped and the patient will be asked to seek the advice of their primary care doctor or go to the emergency room if symptoms are becoming more worrisome.

Sub-Study 3: If the participant at any time has worsening symptoms consistent with new onset of COVID-19 disease a COVID-19 test is performed. If the test is positive for SARS-CoV-2 an appropriately delegated and credentialed study team member will contact IDS and the pharmacist will indicate which arm the participant was assigned. If the participant was assigned to placebo and wishes to cross over to HCQ, IDS will provide HCQ at the same dose as the Sub-Study 3 HCQ arm and IDS will ship the HCQ to the participant. If the participant was assigned to HCQ then the participant will be advised to seek the advice of their personal physician or go to the emergency room if symptoms are becoming more worrisome.

#### 6.0 TREATMENT PLAN

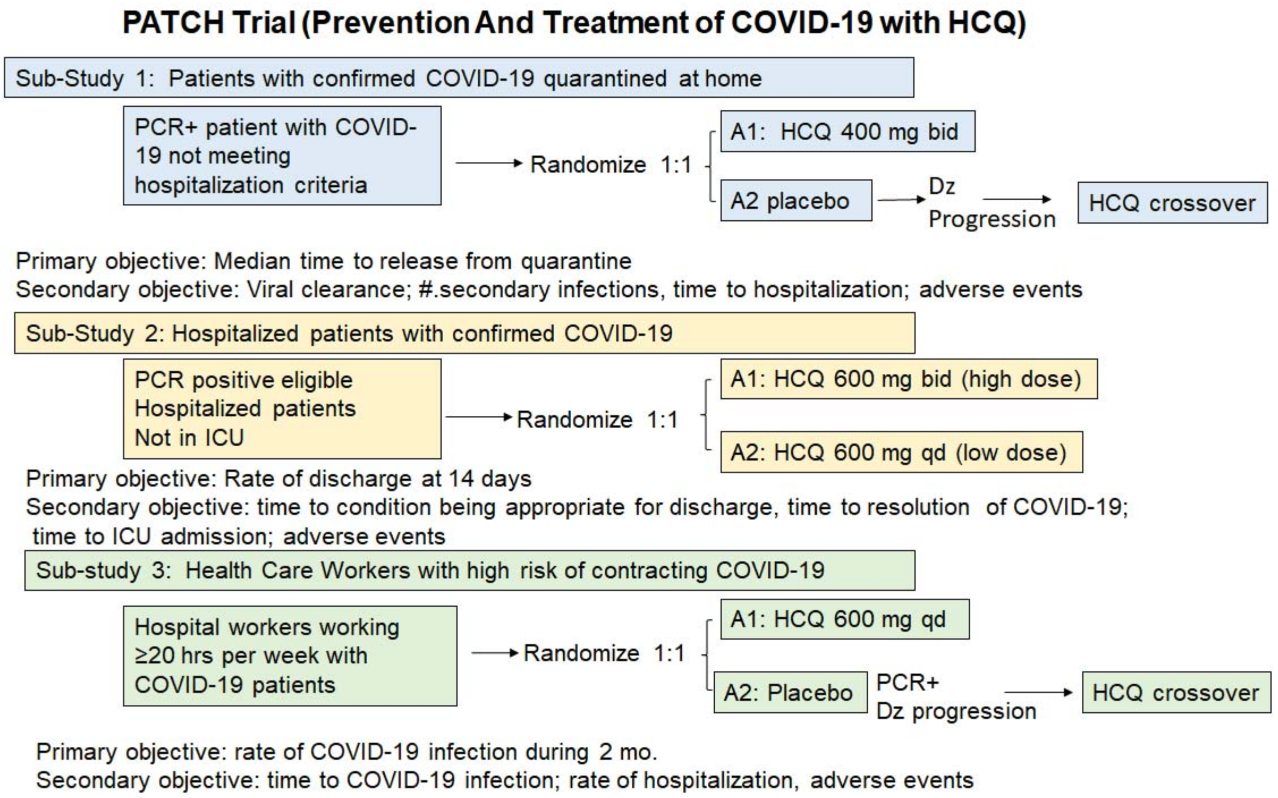

**Fig. 1 PATCH Trial Schema.** Dz: Disease A: Arm

<u>**Sub-Study 1: 100 patients 2 interim analyses**</u>
Arm 1: High dose HCQ: Hydroxychloroquine 400 mg twice a day for up to 14 days
Arm 2: Placebo: 2 pills twice a day for up to 14 days cross over for progressive disease

<u>**Sub-Study 2: 100 patients 2 interim analyses**</u>
Arm 1: High dose HCQ: Hydroxychloroquine 600 mg twice a day for 14 days
Arm 2: Low dose HCQ arm: Hydroxcyhloroquine 600 mg once a day for 7 days

<u>**Sub-Study 3: 200 participants 2 interim analyses**</u>
Arm 1: High dose HCQ: Hydroxychloroquine 600 mg daily for 2 months
Arm 2: Placebo: 3 pills once a day for 2 months crossover to HCQ 600 mg daily if PCR positive

#### 7.0 DETAILS OF STUDY TREATMENT

##### 7.1 Hydroxychloroquine

<u>Mechanism of Action:</u> The mechanism of action is not fully understood. Previously it was thought that HCQ and other chloroquine derivatives are weak bases that deacidify lysosomes through purely chemical basis. Recently our group has identified the missing molecular target of HCQ as palmitoyl protein thioesterase 1 (PPT1).

Storage and formulation: HCQ tablets are manufactured by Sandoz. Each tablet contains 200 mg hydroxychloroquine sulfate (equivalent to 155 mg base). It is dispensed in a tight, light-resistant container as defined in the USP/NF.HCQ should be stored at room temperature up to 30° C (86° F).

Pharmacokinetics: The PK of HCQ is characterized by a large volume of distribution, binding to red blood cells, and long time to peak concentration and steady state. Population PK studies in cancer patients have demonstrated dose proportional change in exposure.

Administration: Hydroxychloroquine is an oral medication, requiring participants in sub-studies 1 and 3 on study to keep an electronic study diary. Diary must be submitted at 14 days (sub-study 1) and 2 months (sub-study 3). Hydroxychloroquine will be provided by IDS. Tablets of HCQ are available in 200 mg strength. When HCQ is administered in divided doses (every 12 hours) two daily doses of HCQ should be taken 12 hours apart, for example, 9 am and 9 pm, and documented clearly on the patient drug diary. The HCQ schedule may be adjusted if necessary to minimize gastrointestinal side effects.

For complete information please refer to the package inserts at http://dailymed.nlm.nih.gov/dailymed/

**7.2 Placebo:** Excipient only manufactured to match hydroxychloroquine. Manufactured at Temple University.

##### 7.3 Concomitant Medication, Drug-Drug interactions and Procedures

Participants in sub-study 3 must be instructed not to take any medications, except tylenol, including over the counter products such as NSAIDs, without first consulting with the investigator. Participants in sub-study 1 must be instructed not to take any medications, including Tylenol and over the counter products such as NSAIDs, without first consulting with the investigator.

**Drug Interactions**

The following medicines are allowed but should be carefully monitored.
Digoxin: Concomitant HCQ and digoxin therapy may result in increased serum digoxin levels.

Insulin or antidiabetic drugs: As HCQ may mildly enhance the effects of a hypoglycemic treatment, a decrease in doses of insulin or antidiabetic drugs may be required.

Drugs that prolong QT interval and other arrhythmogenic drugs: HCQ can prolong the QT interval and should not be administered with other drugs that have the potential to induce cardiac arrhythmias. Also, there may be an increased risk of inducing ventricular arrhythmias if HCQ is used concomitantly with other arrhythmogenic drugs.

Antiepileptics: The activity of antiepileptic drugs might be impaired if co-administered with HCQ.

Cyclosporin: An increased plasma cyclosporin level was reported when cyclosporin and HCQ were co-administered.

Because HCQ has known effects on P450 enzymes, patients requiring anti-convulsants may be treated with any of the non-enzyme inducing anti-convulsants which include: felbamate, valproic acid, gabapentin, lamotrigine, tiagibine, topiramate, or levetiracetam. Due to the fact that both zonisamide and HCQ accumulate in red blood cells, zonisamide should be avoided if possible. All other concomitant medications are permitted.

The following medications are not allowed during the study. The sponsor must be notified if the subject receives any of these during the study:

1. Sub-studies 1 and 2: Any investigational or off-label antiviral therapy

2. Immunosuppressive medications, including, corticosteroids at doses exceeding 10mg/day of prednisone or equivalent, methotrexate, azathioprine, and TNF-alpha blockers. Use of immunosuppressive medication for the management of study treatment-related AEs or in subjects with contrast allergies is acceptable. In addition, use of topical, inhaled and intranasal corticosteroids is permitted

3. Live attenuated vaccines during the study

4. Herbal and natural remedies must be avoided

**7.4 Duration of Protocol Treatment and Follow-up.**

Sub-Study 1 duration of treatment is up to 2 weeks or at the time of release from quarantine
Sub-Study 2 duration of treatment if one week (low dose) versus 2 weeks (high dose) discharge criteria according the hospital practice. If the patient is discharged before the prescribed treatment is completed then the treatment course is discontinued at the time of discharge. If patient has progressive disease that requires intubation the HCQ will be stopped.
Sub-Study 3 duration of treatment is 2 months with assay to assess SARS CoV2 every 2 weeks.

**7.5 Crossover from Control to HCQ arm**. For Sub-Study 1 all participants are un-blinded upon symptom progression (see response assessment below) after at least 7 days of treatment or upon hospitalization if this occurs prior to 7 days of treatment. If the participant was assigned to placebo, he/she has the option to crossover to receive HCQ for up to 2 weeks or until criteria for release from quarantine are met (See 1.0 primary endpoint). In sub-study 3 participants who have worsening symptoms as defined as fever, significantly worsening cough, shortness of breath or hypoxia but do not require hospitalization and tests positive for SARS-CoV2 virus are allowed to cross over to receive HCQ, or treated with an alternative therapy per physician's choice. If these participants need hospitalization and meet eligibility for Sub-Study 2, they can be enrolled into Sub-Study 2 as a new participant. In this case they would be registered as a new participant according to Section 4.

**8.0 TOXICITY CRITERIA, MONITORING, DOSE DELAYS AND MODIFICATIONS**

**8.1 Toxicity Criteria**

This study will utilize the Common terminology criteria for advers events version 5.0 (CTCAE v 5.0). https://ctep.cancer.gov/protocolDevelopment/electronic_applications/docs/CTCAE_v5_Quick_Reference_8.5x11.pdf All appropriate treatment areas should have access to a copy of the CTCAE v 5.0 grading tables.

**8.2 Dose Delays**

Major Events are grade 3 and 4 hematologic and non-hematologic toxicities that are not treatment -related. Treatment should be delayed for major events if HCQ may further complicate the non-treatment related event. If a major event requires a delay of treatment, treatment must be delayed until toxicity is resolved (≤ Grade 2 or ≤ Baseline). For treatment-related toxicities and major events, if toxicity is not resolved in ≤ 14 days, patient will be taken off treatment unless there is an exception granted by the medical monitor.

**8.3 Ocular toxicity.** The only toxicity that requires discontinuation of HCQ is retinopathy. Published literature indicates HCQ retinopathy is idiosyncratic and is uncommon in patients receiving HCQ for less than a few years. Our ongoing study of dabrafenib, trametinib, and HCQ (BAMM; NCTNCT02257424) found no clinically meaningful ocular toxicity in 10 patients studied extensively with serial ocular exams (25). Although HCQ retinotoxicity can occur at shorter than expected intervals with the use of high dose (>5 mg/Kg/day of real body weight) treatment protocols, the cumulative doses required to elicit a retinopathy were only reached after about a year of treatment (26). Thus, we do not anticipate cumulative doses in this trial will reach retinotoxic levels (>200 grams) and have, therefore, not included mandatory ocular exams in this protocol. However, participants will be advised to report with any visual changes at which point a comprehensive ophthalmic exam will take place to decide whether HCQ should be permanently discontinued. As a precaution, patients with renal failure will also be excluded from the trial.

**8.3 Laboratory monitoring is not recommended for HCQ use in rheumatoid arthritis.** The American College of Rheumatology guidelines for the treatment of rheumatoid arthritis recommends no laboratory testing for outpatients that are started on hydroxychloroquine (27). HCQ doses for rheumatoid arthritis range from 400mg daily to 400mg twice a day encompassing the dosing paradigms in Sub-study 1 and sub-study 3. Sub-study 2 will include patients with greater severity of COVID-19 illness and therefore baseline laboratory assessment is warranted.

**8.4 Safety of HCQ in pregnancy obviates the need for pregnancy monitoring.** Numerous case series and a recent large scale epidemiological study demonstrate the safety of HCQ in outpatient pregnant women with systemic lupus erythematous (28).Therefore we are not requiring pregnancy test prior to starting HCQ in sub-studies 1 and 3. Potential participants that are known to be pregnant are excluded. In sub-study 2 in the hospital setting with patients with more severe COVID-19, who are at risk for multi-organ failure we will be requiring a pregnancy testing because the safety of HCQ in pregnancy is not established in acute illnesses requiring hospitalization and with potential for multiorgan failure.

**8.5 Participant safety monitoring and data collection**

**8.5.1 Sub-Study 1:**

Clinical symptom monitoring: Participants will be called by an appropriately delegated and credentialed member of the study team (as example: research nurse) daily at home to monitor clinical symptoms according to the following grading scales:

| Table 2. Example of Daily COVID-19 Symptom tracking score table | | | | |
|---|---|---|---|---|
| Symptom | Mild =1 | Moderate =2 | Severe =3 | Score |
| Fever | | | | 0 |
| Fatigue | | | | 0 |
| Cough | | 2 | | 2 |
| Shortness of breath | 1 | | | 1 |
| Diarrhea | | | | 0 |
| Myalgias | | 2 | | 2 |
| Headache | | | | 0 |
| Smell disturbance | | | | 0 |
| Other: | | | | 0 |
| Total Score | | | | 5 |

The participant will have a daily diary where this table can be filled in by the participant. The study team member will ask for each symptom and record the answers on an eCRF. At the end of the study the concordance of the participant recorded diary and the assessment made by the study team member will be determined. For discordant cases the study team member recorded AEs will be utilized for data analysis.

AE assessment: HCQ can cause nausea, constipation, diarrhea, rash. Participants will be asked if there have been changes in vision such as distortions of blurred vision. Diarrhea is a known toxicity of HCQ that overlaps with the COVID-19 diarrhea. In this case diarrhea will be scored as a COVID-19 symptom to assess the primary endpoint and if diarrhea is worsening while other symptoms are resolving it will be reclassified as a treatment related AE.

Triage: An appropriately delegated and credentialed member of the study team (as example: research nurse) will document treatment-related AEs, and triage the participant for continued home quarantine versus hospitalization. Follow up with an appropriately delegated and credentialed physician investigator will be undertaken for assessment and grading of events.
Temperature Measurements: Home quarantined participants will be asked to take twice daily temperature and record the measurements on their phone. For those patients that do not have a thermometer, one will be provided by the study. The temperature records in the participant diary will be reviewed for trends within participants and in HCQ v. placebo cohorts.

Oral fluid collection: Participants will be asked to collect oral fluid using the RNAPro-Sal collection kit manufactured by Oasis Diagnostics (https://4saliva.com/products/rnapro-sal/). These kits will be provided to the participant with the drug supply. The participant will have clear instructions on how to collect oral fluid on themselves. This process consists of inserting the gauze tip in the mouth for 4 minutes, removing and pushing the syringe to squeeze the fluid into the container. A video will be sent to the participant's email address and texted to them providing visual instructions on use. Participants should produce an early morning saliva sample from the posterior oropharynx (ie, coughed up by clearing the throat) before toothbrushing and breakfast, because nasopharyngeal secretions move posteriorly and bronchopulmonary secretions move by ciliary activity to the posterior oropharyngeal area while the patients are in a supine position during sleep. . Participants will be instructed to collect oral fluid at the following time-points in the containers provided (before taking the first dose of study drug, day 3, day 7). The collected oral fluid should be placed in the freezer in a sealed container. This container will be collected after the participant is released from quarantine. The participant will then wipe the container surface with disinfectant and the ED van will come to pick up the sample. The van driver will open a bag and the participant will carefully place the container in the bag, so that the outside of the bag is not contaminated. If the participant becomes hospitalized with COVID-19, a primary contact for the participant will be contacted and if asymptomatic and/or COVID-19 negative the driver will pick up the saliva using the same procedure.

Pill diary: When the participant is released from quarantine the electronic pill diary must be submitted electronically.

Monitoring co-inhabitants: Appropriately delegated and credentialed study team members (as example: Clinical Research Coordinator) will ask the participant about the status of co-inhabitants daily to 14 days after the quarantine is completed to determine if the participant's co-inhabitants did or did not have symptoms or tested positive for COVID-19.

**8.5.2 Sub-Study 2:** Hospitalized participants will be cared for the assigned inpatient team and monitored daily according clinical practice standards.

Research Blood draws: Appropriately delegated and credentialed study team members (as example: Clinical Research Coordinator) will arrange/coordinate research blood on Day 0, day 3, day 7, Day 14 in coordination with UPHS Staff nurse assigned to participant. These study team members will leave appropriate blood tubes on the outside of the participant's door discretely marked research and coordinate with UPHS staff nurse assigned to participant when routine labs are obtained on the participant. At the time of routine lab collection, research blood will be collected and bagged. The bag will be cleaned with 70% ethanol and placed in a clean secondary bag and handed to the study team member who will transport this bag in a sealed container to TTAB for processing and storage.

Oral fluid collection: Participants will be asked to collect oral fluid using the RNAPro-Sal collection kit manufactured by Oasis Diagnostics (https://4saliva.com/products/rnapro-sal/). These kits will be provided to the participant with the drug supply. The participant will have clear instructions on how to collect oral fluid on themselves. This process consists of inserting the gauze tip in the mouth for 4 minutes, removing and pushing the syringe to squeeze the fluid into the container. A video will be sent to the participant's email address and texted to them providing visual instructions on use. Participants should produce an early morning saliva sample from the posterior oropharynx (ie, coughed up by clearing the throat) before tooth brushing and breakfast, because nasopharyngeal secretions move posteriorly and bronchopulmonary secretions move by ciliary activity to the posterior oropharyngeal area while the patients are in a supine position during sleep. Participants will be instructed to collect oral fluid at the following time-points in the containers provided (before taking the first dose of study drug, day 3, day 7). The collected oral fluid should be placed in the freezer in a sealed container. This container will be collected after the participant is released from quarantine. The participant will then wipe the container surface with disinfectant and the ED van will come to pick up the sample. The van driver will open a bag and the participant will carefully place the container in the bag, so that the outside of the bag is not contaminated. If the participant becomes hospitalized with COVID-19, a primary contact for the participant will be contacted and if asymptomatic and/or COVID-19 negative the driver will pick up the saliva using the same procedure.

AE assessment: Appropriately delegated and credentialed study team members (as example: Research Nurse) will review and abstract from EMR hospital records and a daily patient-reported AE chart remotely to document AEs. The study team member may also contact the participant remotely via telephone communication for documentation of AEs. Daily Vital Sign assessments will also be abstracted from the EMR hospital records and recorded on case report forms. Follow up with an appropriately delegated and credentialed physician investigator will be undertaken for assessment and grading of events. Laboratory test results will be extracted from the EMR. Date of admission and discharge will be used to assess the primary endpoint of rate of discharge at 14 days.

**8.5.3 Sub-Study 3:**

SARS-CoV-2 testing: If available, hospital workers will be tested for SARS-CoV-2 virus by RT-PCR every month in the hospital employee testing site. This assay is not required for participation in the study.
Clinical Symptom monitoring: Appropriately delegated and credentialed study team members (as example: CRC) will screen for symptoms 1X per week and if present will document the AEs. Follow up with an appropriately delegated and credentialed physician investigator will be undertaken for assessment and grading of events. Participants will be asked if there have been changes in vision such as distortions of blurred vision. The rate of conversion to PCR positive in each arm at the 2 month mark will be used to assess the primary endpoint.

Pill diary: Electronic pill diary will be turned in by the participant at the end of the study.

Oral fluid collection: Participants will be asked to collect oral fluid using the RNAPro-Sal collection kit manufactured by Oasis Diagnostics (https://4saliva.com/products/rnapro-sal/). These kits will be provided to the participant with the drug supply. The participant will have clear instructions on how to collect oral fluid on themselves. This process consists of inserting the gauze tip in the mouth for 4 minutes, removing and pushing the syringe to squeeze the fluid into the container. A video will be sent to the participant's email address and texted to them providing visual instructions on use. Participants should produce an early morning saliva sample from the posterior oropharynx (ie, coughed up by clearing the throat) before tooth brushing and breakfast, because nasopharyngeal secretions move posteriorly and bronchopulmonary secretions move by ciliary activity to the posterior oropharyngeal area while the patients are in a supine position during sleep. Participants will be instructed to collect oral fluid at the following time-points in the containers provided (before taking the first dose of study and every 2 weeks after that). The collected oral fluid should be placed in the freezer in a sealed container. This container will be collected after the participant is at the end of their participation. The participant will then wipe the container surface with disinfectant and return to site. If the participant becomes quarantined or hospitalized with COVID-19, a primary contact for the participant will be contacted and if asymptomatic and/or COVID-19 negative the driver will pick up the saliva using the same procedure.

**8.6 Hydroxychloroquine Dose Reduction.**
In outpatient arms 1 and 3, any AE of ≥ Grade 2 and attributed as possibly, probably or definitely related solely to HCQ will result in dose reduction of HCQ as described in Table 5. No more than 2 dose reductions are allowed.

**Table 3: Hydroxychloroquine Dose Reduction Schema**

| Sub-Study | Dose mg/day | First dose reduction | Second Dose reduction |
|---|---|---|---|
| 1 | 400 mg twice daily | 400mg daily | 200 mg daily |
| 2 High dose | 600 mg twice daily | 400 mg twice daily | 400mg daily |
| 2 Low dose | 600 mg daily | 400 mg daily | 200 mg daily |
| 3 | 600 mg daily | 400 mg daily | 200 mg daily |

- If the second dose reduction is not tolerated, then the patient should discontinue the treatment.

Toxicities that may be attributable to HCQ include: nausea, anorexia, vomiting, constipation, diarrhea, rash, and visual field deficit. If any of these AEs occur at grade < 2, HCQ may be continued and the AE managed with supportive care. For any AE with a grade ≥ 3, HCQ dose will be held until the toxicity resolves to ≤ grade 2, after which HCQ may be restarted at a reduced dose as described in table 5. With particular regard to visual field deficits participants should be cautioned to report any visual symptoms, particularly difficulty seeing entire words or faces, image distortions, intolerance to glare, decreased night vision, or loss of peripheral vision. **These symptoms of retinal toxicity or subclinical evidence of retinal toxicity on eye exam should prompt drug discontinuation and ophthalmologic evaluation at Scheie Eye Institute Urgent care.** The natural course of COVID_19 is not fully understood and additional toxicities attributable to HCQ may emerge when used to treated COVID-19.

8.7 Procedure for handling of potentially contaminated material: These materials should be disposed of in hazardous waste containers in sub-study 2. For sub-study 1 institutional and CDC guidelines will be followed. Patients and subjects on sub-studies 1 and 3 will be instructed on safe practices for storage of oral fluid and disinfecation of outer layers of bags and receptacles. Proper hand washing techniques will be reviewed with subjects. Changes in recommendations from the CDC or the University of Pennsylvania will be immediately implemented.

**9.0 SCHEDULE OF EVENTS**

| Table 4. Schedule of events for sub-study 1 | | | |
|---|---|---|---|
| | **Before treatment** | **During treatment** | **Day of release from quarantine** |
| Eligibility checklist | X | | |
| Temperature (patient obtained) | | Twice daily | X |
| COVID-19 symptom score | | Daily | |

| Table 4. Schedule of events for sub-study 1 | | | |
|---|---|---|---|
| | **Before treatment** | **During treatment** | **Day of release from quarantine** |
| AE assessment, triage | | Daily as needed | X |
| SARS-COV-2 PCR assay[1] | X | | |
| Oral fluid collection | X | D3, 7 | |
| **TREATMENT** | | | |
| Hydroxychloroquine or placebo | | Twice every day | Return electronic diary |

| Table 5. Schedule of events for Sub-study 2 | | | |
|---|---|---|---|
| | **Baseline / Pre-RX[a]** | **As clinically indicated during hospital admission** | **On discharge day** |
| History and physical examination | X | X | X |
| Temp, BP, HR, pulse Ox | X | X | X |
| EKG | X | Every 1-2 days | |
| Chest xray | X | X | |
| CBC with differential | X | X | |
| Sodium, potassium, BUN, serum creatinine, glucose, SGOT (AST), SGPT (ALT), total bilirubin, alkaline phosphatase, albumin; PT, PTT, Ferriting, CRP, D-dimer | X | X | |
| RT-PCR SARS-COV-2 | X | X | X |
| Serum BHCG[3] | X | | |
| Oral fluid collection[4] | X | D3, 7 | |
| **Research Whole blood [1,2]** | X | D0, d3, d7, d14 | X |
| **TREATMENT** | | | |
| Hydroxychloroquine | | Daily low dose for 7 days or twice daily high dose for 14 days | |

Table 4 footnotes:

1-Four tubes of blood to be collected as follows: one 5-mL purple top ($K_2EDTA$), two 10-mL green top (sodium heparin) tubes and one 10-mL tiger top serum separator tubes (SST). These tubes will be processed into components (i.e. EDTA plasma and buffy coat for RNA extraction, heparinized plasma and mononuclear cells, and serum) that will be cryopreserved for the planned assays.

2- Research Blood draws are optional and should be coordinated with scheduled phlebotomy draws. No more than 60 mL total can be removed for clinical and research abs at any one blood draw.

3-The safety of the patient and research staff and hospital personnel is critical and if there are any concerns research blood draws should not be performed.

4-A limited number of these collections will be made to further validate the assay and enable its use in sub-studies 1 and 3.

**Table 6. Schedule of events for Sub-study 3**

| | **Baseline / Pre-RX** | **During 2 months** | **End of 2 months** |
|---|---|---|---|
| Eligibility checklist | X | | |
| COVID-19 symptom score | X | Weekly | |
| AE check by telephone | | 1X/week | X |
| PCR SARS-COV-2[1] | X | One month | X[2] |
| Oral fluid collection | X | Every 2 weeks | X |
| Research Blood [3] | X | One month | X |
| **TREATMENT** | | | |
| Hydroxychloroquine or placebo | | 3 pills daily | |

1- this testing is dependent on availability of the test kits and can be deferred if test kits or testing capacity are not available

2- End of 2 months or at the time of symptom progression

3- one 10-mL tiger top serum separator tubes (SST).

#### 10.0 MEASUREMENT OF EFFECT

##### 10.1 Definitions

Evaluable for toxicity. All participants will be evaluable for toxicity from the time of their first treatment with HCQ or placebo.

Evaluable for primary outcome: Participants who received at least one dose of study drug will be evaluable for the primary outcome.

##### 10.2 Response Criteria

Sub-Study 1: Time to release from quarantine. The criteria is afebrile for 72 hours AND improvement of symptoms AND at least 7 days have past since symptoms started. To quantitatively assess improvement of symptoms the grading system in Section 8.5 will be used.

Sub-study 2: Time to discharge is the primary outcome, length of hospital stay will be defined as length of time in days since admission to the hospital to discharge from the hospital. Rate of intensive care admission (ICU) admission is defined as the percentage of participants that require ICU level care. Survival rate is the percentage of participants surviving at the end of study.

Sub-Study 3: Measurement of SARS-CoV-2 positivity defined by PCR. Time to infection will be defined as the time in days from enrollment on the study to time to COVID-19 symptoms.

##### 10.3 Off treatment/Off Study

Each participant has the right to withdraw from the study at any time without prejudice. The investigator may discontinue any persons participation for any reason, including adverse event or failure to comply with the protocol. Should a participant withdraw from the study, the reason(s) must be stated on the case report form, and a final evaluation of the participant should be performed. Reasons for withdrawal include the following:

Progression of Disease: Remove participant from protocol therapy at the time progressive disease is documented. Progression of disease is left t the discretion of the treating physician/ sub-I, however the following guidelines are provided to help decision making for discontinuing study drug:

Substudy 1: Progression of disease can be confirmed if there is increase in COVID-19 symptom score for 2 days in a row and is three points higher than baseline

Substudy2: Progression of disease is defined as subject is anticipated to need mechanical ventilation in the next 24 hours

Substudy 3: Development of new symptomotology consistent with COVID-19 : e.g. fever, cough, shortness of breath.

Extraordinary Medical Circumstance: If at any time the treating physician feels constraints of this protocol are detrimental to the participant’s health remove the participant from protocol therapy.

Participant’s refusal to continue treatment: In this event, document the reason(s) for withdrawal.

Failure to comply with protocol (as judged by the investigator such as compliance below 80%, failure to maintain appointments, etc.).

Delay in treatment > 7 days due to toxicity

#### 11.0 ADVERSE EVENTS AND REPORTING

The timely reporting of adverse events (including toxic deaths) is required by the Food and Drug Administration. The reporting of toxicities is part of the data reporting for this study. The sponsor-investigator is responsible for ensuring that all adverse events (AEs) and significant adverse events (SAEs) that are observed or reported during the study are collected.

Adverse Events

An ***adverse event*** (AE) is any symptom, sign, illness or experience that develops or worsens in severity during the course of the study that does not necessarily have a causal relationship with this treatment. Intercurrent illnesses or injuries should be regarded as adverse events. Abnormal results of diagnostic procedures are considered to be adverse events only if the abnormality:

- results in study withdrawal
- is associated with a serious adverse event
- is associated with clinical signs or symptoms
- leads to additional treatment or to further diagnostic tests is considered by the investigator to be of clinical significance

**Adverse Event Reporting Period**

The study period during which adverse events must be reported is defined as the period from the initiation of the first study treatment to the last administration of study treatment.

**Post-study Adverse Event.**

All unresolved adverse events should be followed by the clinical investigator until the events are resolved, the participant is lost to follow-up, or the adverse event is otherwise explained. At the last scheduled visit, the investigator should instruct each participant to report any subsequent event(s) that the participant, or the participant's personal physician, believes might reasonably be related to participation in this study.

**Abnormal Laboratory Values.**

A clinical laboratory abnormality should be documented as an adverse event if the abnormality is grade 1 or more and a change from baseline.

##### 11.1 Recording of Adverse Events.

At each contact with the participant, the appropriately delegated and credentialed study team member must seek information on adverse events by specific questioning and, as appropriate, by examination. Information on all adverse events should be recorded immediately in the source document, and also in the appropriate adverse event module of the case report form (CRF). All clearly related signs, symptoms, and abnormal diagnostic procedures results should be recorded in the source document, though should be grouped under one diagnosis.

All adverse events occurring during the study period must be recorded. Adverse events will be measured and graded in accordance with the CTCAE. The clinical course of each event should be followed until resolution, stabilization, or until it has been determined that the study treatment or participation is not the cause. Serious adverse events that are still ongoing at the end of the study period must be followed up to determine the final outcome. Any serious adverse event that occurs after the study period and is considered to be at least possibly related to the study treatment or study participation should be recorded and reported immediately.

###### 11.1.1 Serious Adverse Events

Adverse events are classified as serious or non-serious.

A serious adverse event is any AE that is:

- fatal
- life-threatening
- requires or prolongs hospital stay
- results in persistent or significant disability or incapacity
- a congenital anomaly or birth defect
- Suspected transmission of an infectious agent (eg, pathogenic or nonpathogenic) via the study drug is an SAE.
- an important medical event

Important medical events are those that may not be immediately life threatening, but are clearly of major clinical significance. They may jeopardize the participant, and may require intervention to prevent one of the other serious outcomes noted above. For example, drug overdose or abuse, a seizure that did not result in in-patient hospitalization, or intensive treatment of bronchospasm in an emergency department would typically be considered serious. Theft, sale, or use of the study product by any person other than the participant will be reported as a medically important event.

**ADVERSE EVENT COLLECTION AND REPORTING INFORMATION:**

- All Serious Adverse Events (SAEs) that occur following the participant's written consent to participate in the study must be reported, whether related or not related to study drug. If applicable, SAEs must be collected that relate to any later protocol-specified procedure.
- Following the participant's written consent to participate in the study, all SAEs, whether related or not related to study drug, are collected, including those thought to be associated with protocol-specified procedures. The investigator-sponsor should report any SAE occurring after these aforementioned time periods, which is believed to be related to study drug or protocol-specified procedure.
- An SAE report should be completed for any event where doubt exists regarding its seriousness;
- If the investigator believes that an SAE is not related to study drug, but is potentially related to the conditions of the study (such as withdrawal of previous therapy or a complication of a study procedure), the relationship should be specified in the narrative section of the SAE Report Form.

All SAEs should be followed to resolution or stabilization.

Adverse events can be spontaneously reported or elicited during open-ended questioning, examination, or evaluation of a participant. (In order to prevent reporting bias, participants should not be questioned regarding the specific occurrence of one or more AEs.)

**All adverse events that do not meet any of the criteria for serious should be regarded as non-serious adverse events. Non-serious adverse events are documented and assessed as adverse events noted above.**

**Hospitalization, Prolonged Hospitalization or Surgery.**

Any adverse event that results in hospitalization or prolonged hospitalization should be documented and reported as a serious adverse event unless specifically instructed otherwise in this protocol. Any condition responsible for surgery should be documented as an adverse event if the condition meets the criteria for an adverse event.

Neither the condition, hospitalization, prolonged hospitalization, nor surgery are reported as an adverse event in the following circumstances:

- Hospitalization or prolonged hospitalization for diagnostic or elective surgical procedures for a preexisting condition. Surgery should not be reported as an outcome of an adverse event if the purpose of the surgery was elective or diagnostic and the outcome was uneventful.
- Hospitalization or prolonged hospitalization required to allow efficacy measurement for the study.
- Hospitalization or prolonged hospitalization for therapy of the target disease of the study, unless it is a worsening or increase in frequency of hospital admissions as judged by the clinical investigator.

##### 11.2 Assessment of Adverse Events

All AEs and SAEs whether volunteered by the participant, discovered by study personnel during questioning, or detected through physical examination, laboratory test, or other means will be reported appropriately. Each reported AE or SAE will be described by its duration (i.e., start and end dates), regulatory seriousness criteria if applicable, suspected relationship to the study drug (see following guidance), and actions taken. To ensure consistency of AE and SAE causality assessments, only the PI or physician sub-investigators may grade AEs, and investigators should apply the following general guideline:

###### 11.2.1 Relationship to study drug: Yes

There is a plausible temporal relationship between the onset of the AE and administration of the study drug, and the AE cannot be readily explained by the participant's clinical state, intercurrent illness, or concomitant therapies; and/or the AE follows a known pattern of response to the study drug; and/or the AE abates or resolves upon discontinuation of the study drug or dose reduction and, if applicable, reappears upon re-challenge.

###### 11.2.2 Relationship to study drug: No

Evidence exists that the AE has an etiology other than the study drug (e.g., preexisting medical condition, underlying disease, intercurrent illness, or concomitant medication); and/or the AE has no plausible temporal relationship to study drug administration (e.g., cancer diagnosed 2 days after first dose of study drug).

Expected adverse events are those adverse events that are listed or characterized in the Package Insert.

Unexpected adverse events are those not listed in the Package Insert. This includes adverse events for which the specificity or severity is not consistent with the description in the Package Insert. For example, under this definition, hepatic necrosis would be unexpected if the Package Insert only referred to elevated hepatic enzymes or hepatitis.

###### 11.2.3 Diagnosis vs. Signs and Symptoms

If known at the time of reporting, a diagnosis should be reported rather than individual signs and symptoms (e.g., record only liver failure or hepatitis rather than jaundice, asterixis, and elevated transaminases). However, if a constellation of signs and/or symptoms cannot be medically characterized as a single diagnosis or syndrome at the time of reporting, it is ok to report the information that is currently available. If a diagnosis is subsequently established, it should be reported as follow-up information.

###### 11.2.4 Deaths

All deaths that occur during the protocol-specified AE reporting period (see Section 12.1), regardless of attribution, will be reported to the appropriate parties. When recording a death, the event or condition that caused or contributed to the fatal outcome should be reported as the single medical concept. If the cause of death is unknown and cannot be ascertained at the time of reporting, report "Unexplained Death".

###### 11.2.5 Preexisting Medical Conditions

A preexisting medical condition is one that is present at the start of the study. Such conditions should be reported as medical and surgical history. A preexisting medical condition should be re-assessed throughout the trial and reported as an AE or SAE only if the frequency, severity, or character of the condition worsens during the study. When reporting such events, it is important to convey the concept that the preexisting condition has changed by including applicable descriptors (e.g., "more frequent headaches").

##### 11.2.6 Pregnancy

Female Participants
Participants should not become pregnant while on this study and for 90 days after last dose of study drug. In addition, participants should not breastfeed while on this study as these drugs may also affect a breast-feeding child. Pregnant women and women who are breast-feeding are not allowed to participate in this study. Participants must agree to use two medically accepted forms of birth control including condoms, diaphragms, cervical cap, an intra-uterine device (IUD), surgical sterility (tubal ligation or a partner that has undergone a vasectomy), or oral contraceptives, OR must agree to completely abstain from intercourse for at least two weeks before receiving first dose of study drug, during participation in this study and for 90 days after last dose of study drug. Abstinence at certain times of the cycle only, such as during the days of ovulation, after ovulation and withdrawal are not acceptable methods of birth control. Even when an approved contraceptive method is used, there is always a small risk that the participant could still become pregnant.

Male Participants
Participants should not father a child while on this study. If the participants spouse or partner has the potential to become pregnant, the participant and partner must use two medically accepted forms of birth control including condoms, diaphragms, cervical cap, an intra-uterine device (IUD), surgical sterility (vasectomy or a partner that has undergone a tubal ligation), or oral contraceptives, OR must agree to completely abstain from intercourse during participation in this study and for 90 days after last dose of study drug. If the partner is taking oral contraceptives, she must begin taking them at least two weeks before the participants first dose of study drug. Abstinence at certain times of the cycle only, such as during the days of ovulation, after ovulation and withdrawal are not acceptable methods of birth control.

If a female participant becomes pregnant while receiving investigational therapy or within 90 days after the last dose of study drug, the clinical investigator must immediately notify sponsor-investigator (who will then notify Novartis/Sandoz) in accordance with SAE reporting guidelines. Follow-up to obtain the outcome of the pregnancy should also occur. Abortion, whether accidental, therapeutic, or spontaneous, should always be classified as serious, and expeditiously reported as an SAE. Similarly, details of the birth, and the presence or absence of any congenital anomaly/birth defect or maternal and/or newborn complications in a child born to a female participant exposed to the study drug should be reported as an SAE. Follow-up information regarding the course of the pregnancy, including perinatal and neonatal outcome and, where applicable, offspring information must be reported to the sponsor-investigator (and Novartis/ Sandoz). In order for sponsor-investigator or designee to collect any pregnancy surveillance information from the female participant a signed informed consent addendum for disclosure of this information must be obtained (and will be supplied to the IRB for review at the time the incidental pregnancy is encountered, not anticipated and therefore not included with the original protocol documents). Any pregnancy that occurs in a female partner of a male study participant should be reported. Information on this pregnancy will be collected on the Pregnancy Surveillance Form (supplied for IRB review in the event pregnancy in a female partner of a male participant is encountered). In order for sponsor-investigator or designee to collect any pregnancy surveillance information from the female partner, the female partner must sign an informed consent form for disclosure of this information (form to be supplied in the event such a pregnancy were to be encountered).

#### 11.3 UPENN IRB Notification by Investigator-Sponsor

The University of Pennsylvania IRB (Penn IRB) requires expedited reporting of those events related to study participation that are unforeseen and indicate that participants or others are at increased risk of harm. The Penn IRB will not acknowledge safety reports or bulk adverse event submissions that do not meet the criteria outlined below. The Penn IRB requires researchers to submit reports of the following problems within 10 working days from the time the investigator becomes aware of the event:

- Any adverse event (regardless of whether the event is serious or non-serious, on-site or off-site) that occurs any time during or after the research study, which in the opinion of the principal investigator is:

**Unexpected** (An event is "unexpected" when its specificity and severity are not accurately reflected in the protocol-related documents, such as the IRB-approved research protocol, any applicable investigator brochure, and the current IRB-approved informed consent document and other relevant sources of information, such as product labeling and package inserts.)

**AND**

**Related to the research procedures** (An event is "related to the research procedures" if in the opinion of the principal investigator or sponsor, the event was more likely than not to be caused by the research procedures.)

Deaths occurring for patients on-study and within 30 days of study drug administration that are considered unforeseen and indicates participants or others are at increased risk of harm (i.e. unexpected and probably/definitely related), must be reported to the IRB within 24 hours of notification.

See: https://irb.upenn.edu/ for additional reporting details.

##### 11.3.1 Reporting Process to IRB at Penn.

Principal Investigators are encouraged to submit reports of unanticipated problems posing risks to participants or others using the form: "**Unanticipated Problems Posing Risks to Participants or Others Including Reportable Adverse Events**" via HS-ERA or a written report of the event within 7 working days. FDA Notification by Investigator-Sponsor.

This study is IND exempt and reporting to the FDA is voluntary using a MedWatch 3500 or via the FDA's website for voluntary reporting.

##### 11.4 Medical Monitoring

It is the responsibility of the Sponsor-Investigator to oversee the safety of the study at this site. This safety monitoring will include careful assessment and appropriate reporting of adverse events, as noted above. Medical monitoring by an independent clinician, Dr. Sunita Nasta, Department of Medicine, will include a regular assessment of the number and type of serious adverse events on a periodic basis.

##### 11.7 Study Monitoring Plan

This study will be monitored by the Principal Investigator and sub-investigators, as appropriate. Such monitoring will include at least weekly meetings of the study team to review accrual, toxicity, SAEs. Dose escalations and study finding. In addition, the PI will ensure that data are completed in a timely manner and he or his designee will review the data for accuracy, completeness and integrity. Further, the PI will view real-time toxicity and all laboratory results on an ongoing basis by accessing the Monitoring Report database and the Protocol Labs database.

##### 11.8 Auditing and Inspecting

The Investigator is required to permit direct access to the facilities where the study takes place, source documents, CRFs and applicable supporting records of study subject participation for audits and inspections by IRB, regulatory authorities (FDA) and academic authorized representatives (OCR). The Investigator will make every effort to be available for the audits and/or inspections.

#### 12.0 Sample collection and processing for Correlative Science

#### 12.1 Research Specimen Process, Banking and Analysis

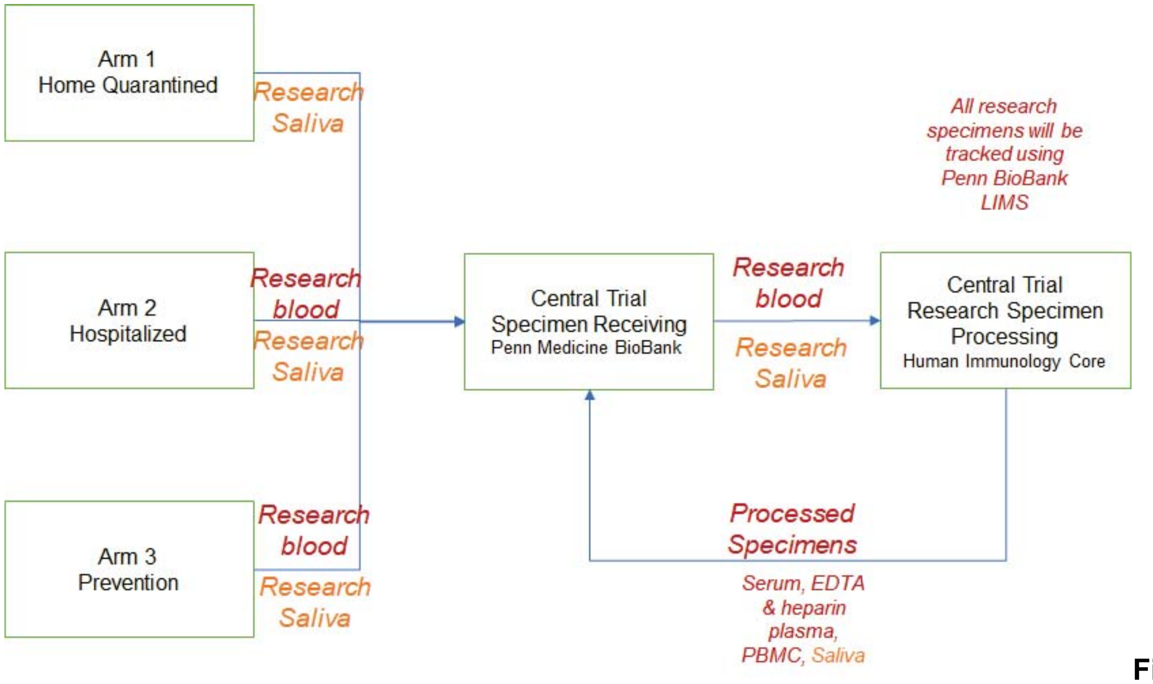

**Figure 2.**

**Sample flow for correlative samples that will be banked.**

Blood and oral fluid specimens collected as delineated in 9.0 Schedule of Events. Samples will be assigned a unique identified and then processed and frozen as per SOP within a BSL2+ space within the Human Immunology Core that is approved by the University of Pennsylvania's Environmental Health and Radiation Safety (EHRS) program. The processed samples will be stored in the Penn BioBank at the University of Pennsylvania within locked and temperature monitored freezers, and sample tracking will be conducted using the Penn BioBank Laboratory Information Management System (LIMS). Documentation for sample receipt and processing will be collected and stored in either locked cabinet for paper records or within an encrypted and password protected database maintained within the HIC. The planned research analyses outlined in the exploratory objectives will be performed using qualified and if possible validated methods; however, due to nature of these research procedures, they may not conform to principles of good clinical laboratory practice.

#### 13.0 STATISTICAL CONSIDERATIONS

##### 13.1 Sample size calculation

Because we are uncertain about the outcome values for both control and treatment arms of the study, we will use group-sequential methods, initially proposing a real difference that is small, and allowing us to stop early for both efficacy and futility. We conduct two interim analyses and a final analysis, after approximately 1/3, 2/3, and 100% of subjects have completed. We will use the Hwang-Shih-DeCani alpha spending rules. Boundaries for efficacy and futility decisions are show below. Futility boundaries are non-binding.

| z-value | —— Boundaries —— | | Information |
|---|---|---|---|
| **Stage** | **Efficacy** | **Futility** | **Proportion** |
| 1 | 2.7819 | -0.8235 | 0.3400 |
| 2 | 2.2653 | 0.4262 | 0.6800 |
| 3 | 1.6813 | 1.6813 | 1.0000 |

| p-value——— Boundaries ——— | | | Information |
|---|---|---|---|
| **Stage** | **Efficacy** | **Futility** | **Proportion** |
| 1 | 0.00270 | 0.79490 | 0.3400 |
| 2 | 0.01175 | 0.33497 | 0.6800 |
| 3 | 0.04636 | 0.04636 | 1.0000 |

Because we are uncertain about the outcome values for both control and treatment arms of the study, we will use group-sequential methods, initially proposing a real difference that is small, and allowing us to stop early for both efficacy and futility. We conduct two interim analyses and a final analysis, after approximately 1/3, 2/3, and 100% of subjects have completed. We will use the Hwang-Shih-DeCani alpha spending rules. Boundaries for efficacy and futility decisions are show below. Futility boundaries are non-binding.

**Sub-Study 1, Home quarantined patients with COVID-19:** Given the median duration of viremia is 14-20 days in mild cases, we will assume the null hypothesis that in the placebo cohort the median time to release from quarantine is 10 days from receiving a positive test. In order for HCQ to be considered more effective in this treatment setting we aim to see a median time to release from quarantine of 5 days. These values correspond to daily hazard rates of 0.069 and 0.139 respectively. We will randomize 1:1 HCQ to control and with 100 patients with 50 in the placebo arm and 50 in the HCQ arm. The one-sided z- test (alpha=.05) will have an overall 95% power to detect a significant difference between the two groups with medians of 10 and 5 days.

The *hypothesis* will be tested using a one-sided test, with a z-score corresponds to the log of the hazard-ratio for recovery between the two groups. We will obtain estimates of the hazard ratio and group median survival, and 95% CI, from Cox regression. As a secondary analysis, we will stratify the analysis, and obtain separate estimates of the hazard-ratio by group. However, we are not powered to test the interaction for significance, and we do not expect significance within group.

<u>Interim analysis:</u> We will perform two interim analyses targeting 34% and 68% completion, testing for early efficacy or futility, using z-score boundaries that follow Hwang-Shih-DeCani alpha spending rules. The determination of information for survival analysis is somewhat imprecise during the planning stages; for that reason, we will conduct analyses when 34% and 68% of the final sample have had 10 days of followup, and determine boundaries based on actual information available at analysis time. Following those rules, we have 95% power using 100% of the sample, if our under our 5 days versus 10 days scenario is true. Under those circumstances, we expect to have a 18% chance of stopping for early efficacy at the first interim analysis. We also have a 30% chance of stopping early for efficacy at the first interim analysis if the placebo arm really had 12 days median time instead of 10 days . Under normal circumstances (10 days v 5 days), we have a 63% chance of stopping for early efficacy at the second interim analysis. We also have an 82% chance of stopping early for efficacy at the second interim analysis if the placebo arm really has 12 days median time instead of 10

**Z-Value Boundaries**

Maximum Information: 2000.1128
Alternative Hypothesis: h1 - h2 < 0 (one-sided)
Futility Boundaries: Non-Binding

| | ——— Boundaries ——— | | Time | | Information |
|---|---|---|---|---|---|
| **Stage** | **Efficacy** | **Futility** | **Proportion** | **Time** | **Proportion** |
| 1 | -2.7370 | 0.7159 | 0.4000 | 24.00 | 0.3660 |
| 2 | -2.2814 | -0.3990 | 0.6300 | 37.80 | 0.6740 |
| 3 | -1.6808 | -1.6808 | 1.0000 | 60.00 | 1.0000 |

**P-Value Boundaries**
Maximum Information: 2000.1128
Alternative Hypothesis: h1 - h2 < 0 (one-sided)
Futility Boundaries: Non-Binding
P-value boundaries are one-sided values.

| | Boundaries | | Time | | Information |
|---|---|---|---|---|---|
| Stage | Efficacy | Futility | Proportion | Time | Proportion |
| 1 | 0.00310 | 0.76297 | 0.4000 | 24.00 | 0.3660 |
| 2 | 0.01126 | 0.34494 | 0.6300 | 37.80 | 0.6740 |
| 3 | 0.04640 | 0.04640 | 1.0000 | 60.00 | 1.0000 |

**Sub-Study 2, Hospitalized patients:** We will assume the null hypothesis that in the low dose HCQ cohort 70% of hospitalized patients recover by 21 days. In order for high dose HCQ to be considered more effective in the treatment setting we aim to see a 95% recovery (i.e. an effect of at least 25%). We will randomize 1:1 HCQ to control and with 100 patients with 50 in the low dose HCQ arm and 50 in the high dose treatment arm. The one-sided z-test (alpha=.05) based on the log of the odds-ratio will have 90% power to detect a significant difference between the two groups of greater than 25% between the population rates. The analysis will be stratified by radiographic signs of pneumonia at admission and age.

The *hypothesis* will be tested using a one-sided test, with a z-score corresponds to the log of the odds-ratio for recovery between the two groups. We will obtain the estimate of the odds ratio, and 95% CI, from logistic regression. As a secondary analysis, we will stratify the analysis, and obtain separate estimates of the odds-ratio by group. However, we are not powered to test the interaction for significance, and we do not expect significance within group.

Interim analysis: We will perform two interim analyses at 34% and 68% completion, testing for early efficacy or futility, using z-score boundaries that follow Hwang-Shih-DeCani alpha spending rules. Following those rules, we have 95% power using 100% of the sample, if our under our 70% versus 95% scenario is true. Under those circumstances, we have a 17% chance of stopping for early efficacy at the first interim analysis. We also have a 92% chance of stopping early for efficacy at the first interim analysis if the low dose arm really has 30% recovery instead of 70%. Under normal circumstances (70% v 95%), we have a 67% chance of stopping for early efficacy at the second interim analysis. We also have a 90% chance of stopping early for efficacy at the second interim analysis if the low dose arm really has 60% recovery instead of 70%.

| z-value | Boundaries | | Information |
|---|---|---|---|
| Stage | Efficacy | Futility | Proportion |
| 1 | 2.7819 | -0.8235 | 0.3400 |
| 2 | 2.2653 | 0.4262 | 0.6800 |
| 3 | 1.6813 | 1.6813 | 1.0000 |

| p-value | Boundaries | | Information |
|---|---|---|---|
| Stage | Efficacy | Futility | Proportion |
| 1 | 0.00270 | 0.79490 | 0.3400 |
| 2 | 0.01175 | 0.33497 | 0.6800 |
| 3 | 0.04636 | 0.04636 | 1.0000 |

**Sub-Study 3, prophylaxis in hospital workers:** The transmission of SARS-COV2 from patient to hospital worker depends on many factors including specifics of standard care to prevent transmission, but especially on the number of patients seen at a given hospital or outpatient practice. Across China the reported hospital worker infection rate is 3.8%, but in Wuhan it is reported as 58% at the height if the epidemic. We will use a

10% transmission rate as the null hypothesis (low dose group). In order for HCQ to be considered effective our alternative hypothesis will be a 1% transmission rate. With a 1:1 randomization for the HCQ to control arms we would require a total of 200 hospital worker subjects total for Arm 4. With the placebo group of 100 subjects and the h HCQ arm of 100 subjects, a one-sided z-test (alpha=0.05) comparing the rates in the two groups would have an 80% power to detect a significant difference when the difference in the population rates is at least 9%.

The *hypothesis* will be tested using a one-sided test, with a z-score corresponds to the log of the odds-ratio for recovery between the two groups. We will obtain the estimate of the odds ratio, and 95% CI, from logistic regression.

Interim analysis: We will perform two interim analyses at 25% and 50% completion, testing for early efficacy or futility, using z-score boundaries that follow Hwang-Shih-DeCani alpha spending rules. Following those rules, we have 84% power using 100% of the sample, if our under our 10% versus 1% scenario is true. Under those circumstances, we have a 6% chance of stopping for early efficacy at the first interim analysis. We also have an 81% chance of stopping early for efficacy at the first interim analysis if the low dose arm really has 45% infection instead of 10%. Under normal circumstances (10% v 1%), we have a 27% chance of stopping for early efficacy at the second interim analysis. We also have an 80% chance of stopping early for efficacy at the second interim analysis if the low dose arm really has 23% infection instead of 10%.

**Z-Value Boundaries**

| | **Boundaries** | | **Information** |
|---|---|---|---|
| **Stage** | **Efficacy** | **Futility** | **Proportion** |
| 1 | 2.9473 | -1.2318 | 0.2500 |
| 2 | 2.5825 | -0.2676 | 0.5000 |
| 3 | 1.6664 | 1.6664 | 1.0000 |

**P-Value Boundaries**

| | **Boundaries** | | **Information** |
|---|---|---|---|
| **Stage** | **Efficacy** | **Futility** | **Proportion** |
| 1 | 0.00160 | 0.89099 | 0.2500 |
| 2 | 0.00491 | 0.60550 | 0.5000 |
| 3 | 0.04782 | 0.04782 | 1.0000 |

**12.2 Analysis of Secondary Endpoints.**

Secondary outcomes will at least be analyzed as summary statistics, with group means, odds-ratios, and 95% CIs. We may conduct exploratory analyses using regression methods appropriate for each type of measure.

**Sub-Study 1:** Rate of secondary infection of housemates is a binomial count by household. We will summarize the proportion or rate by group, with 95% CI. With sufficient data, we will explore subgroups, patient, and secondary patient characteristics using logistic regression. Rate of hospitalization is a binary measure. We will summarize rates by group, and estimate the odds-ratio for treatment. Adverse event rates will be summarized as a count by subject, and mean count by group, and we will estimate a rate-ratio for treatment.

**Sub-Study 2:** The rate of ICU admission is a binary variable by subject. We will summarize the rates by group, and calculate an odds-ratio for treatment. Time to PCR negativity is a time to event measure, beginning at diagnosis time and ending at negativity or censorship. Time to negativity will be summarized using Kaplan-Meier methods to give time-to-event curves by group, and estimates of median time. We will also estimate a hazard ratio by group. Adverse event rates will be summarized as a count by subject, and mean count by group, and we will estimate a rate-ratio for treatment.

**Sub-Study 3:** For hospital employees, we will summarize the count of shifts missed by group, and estimate the rate ratio for treatment. Adverse event rates will be summarized as a count by subject, and mean count by group, and we will estimate a rate-ratio for treatment.

Property of University of Pennsylvania
Confidential
